# Non-pharmaceutical interventions, vaccination and the Delta variant: epidemiological insights from modelling England’s COVID-19 roadmap out of lockdown

**DOI:** 10.1101/2021.08.17.21262164

**Authors:** Raphael Sonabend, Lilith K. Whittles, Natsuko Imai, Pablo N Perez-Guzman, Edward S Knock, Thomas Rawson, Katy AM Gaythorpe, Bimandra A Djaafara, Wes Hinsley, Richard G FitzJohn, John A Lees, Divya Thekke Kanapram, Erik M Volz, Azra C Ghani, Neil M Ferguson, Marc Baguelin, Anne Cori

## Abstract

**Background:** England’s COVID-19 “roadmap out of lockdown” set out the timeline and conditions for the stepwise lifting of non-pharmaceutical interventions (NPIs) as vaccination roll-out continued. Here we assess the roadmap, the impact of the Delta variant, and potential future epidemic trajectories.

**Methods:** We extended a model of SARS-CoV-2 transmission to incorporate vaccination and multi-strain dynamics to explicitly capture the emergence of the Delta variant. We calibrated the model to English surveillance data using a Bayesian evidence synthesis framework, then modelled the potential trajectory of the epidemic for a range of different schedules for relaxing NPIs.

**Findings:** The roadmap was successful in offsetting the increased transmission resulting from lifting NPIs with increasing population immunity through vaccination. However due to the emergence of Delta, with an estimated transmission advantage of 73% (95%CrI: 68-79) over Alpha, fully lifting NPIs on 21 June 2021 as originally planned may have led to 3,400 (95%CrI: 1,300-4,400) peak daily hospital admissions under our central parameter scenario. Delaying until 19 July reduced peak hospitalisations by three-fold to 1,400 (95%CrI: 700-1,500) per day. There was substantial uncertainty in the epidemic trajectory, with particular sensitivity to estimates of vaccine effectiveness and the intrinsic transmissibility of Delta.

**Interpretation:** Our findings show that the risk of a large wave of COVID hospitalisations resulting from lifting NPIs can be substantially mitigated if the timing of NPI relaxation is carefully balanced against vaccination coverage. However, with Delta, it may not be possible to fully lift NPIs without a third wave of hospitalisations and deaths, even if vaccination coverage is high. Variants of concern, their transmissibility, vaccine uptake, and vaccine effectiveness must be carefully monitored as countries relax pandemic control measures.

**Funding:** National Institute for Health Research, UK Medical Research Council, Wellcome Trust, UK Foreign, Commonwealth & Development Office.

**Research in context:** *Evidence before this study:* We searched PubMed up to 23 July 2021 with no language restrictions using the search terms: (COVID-19 or SARS-CoV-2 or 2019-nCoV or “novel coronavirus”) AND (vaccine or vaccination) AND (“non pharmaceutical interventions” OR “non-pharmaceutical interventions) AND (model*). We found nine studies that analysed the relaxation of controls with vaccination roll-out. However, none explicitly analysed real-world evidence balancing lifting of interventions, vaccination, and emergence of the Delta variant.

*Added value of this study:* Our data synthesis approach combines real-world evidence from multiple data sources to retrospectively evaluate how relaxation of COVID-19 measures have been balanced with vaccination roll-out. We explicitly capture the emergence of the Delta variant, its transmissibility over Alpha, and quantify its impact on the roadmap. We show the benefits of maintaining NPIs whilst vaccine coverage continues to increase and capture key uncertainties in the epidemic trajectory after NPIs are lifted.

*Implications of all the available evidence:* Our study shows that lifting interventions must be balanced carefully and cautiously with vaccine roll-out. In the presence of a new, highly transmissible variant, vaccination alone may not be enough to control COVID-19. Careful monitoring of vaccine uptake, effectiveness, variants, and changes in contact patterns as restrictions are lifted will be critical in any exit strategy.

## Introduction

Despite the UK being the first country to start nationwide vaccination campaigns^1^, the emergence of the Alpha variant of concern (VOC) drove the severe second wave over the 2020-2021 winter leading to a third lockdown in England from 5 January 2021^2^. Informed by mathematical modelling, the UK government published a “roadmap out of lockdown” for England, setting out the conditions for and expected timeline of a step-wise lifting of non-pharmaceutical interventions (NPIs)^3^. Between 29 March and 19 July 2021, NPIs were incrementally lifted as vaccination coverage increased. By 19 July, 87·5% of the adult population in England had received at least one dose of vaccine, and 68·2% had received two doses^4^. The impact of each roadmap step was assessed in real-time against the government’s “four tests” before further interventions were lifted: i) continued success of the vaccine programme; ii) evidence of the effectiveness of vaccines against hospitalisation; iii) no risk of overwhelming the NHS; and iv) new VOCs do not change the risk assessment^3^.

The emergence of VOCs, notably the lineages Alpha, Beta, Gamma, and Delta first detected in the UK,^5^ South Africa,^6^ Brazil,^7^ and India^8^ respectively – has posed recurring challenges for pandemic control efforts globally. The Delta variant was designated a VOC on 6 May 2021 by Public Health England (PHE) and quickly became the dominant variant in the UK^9^. Delta is substantially more transmissible than Alpha^9,10^, a variant already 50-80% more transmissible than previously circulating variants^5,11^. Delta is also estimated to have a 1·85 to 2·6-fold increase in the risk of hospitalisation^12,13^. It is also associated with partial immune escape^14^ and consequent reductions in vaccine effectiveness (VE) and cross-protection from prior non-Delta infections^13–18^.

Delta’s emergence in the UK in mid-April^19^ drove a rapid increase in cases and hospitalisations across all areas of England^9^, prompting the final roadmap step (step four) being delayed by a month to 19 July^20^. Daily case numbers in England started increasing from mid-May 2021, and grew exponentially from mid-June to mid-July, reaching a peak of 56,282 on 15 July. Numbers then unexpectedly and synchronously fell to half that value over the following two weeks, before plateauing and beginning to rise slowly again in August^21^.

Here we quantify the impact of each step of the roadmap from i) school re-opening; ii) outdoor hospitality and non-essential retail; and iii) indoor hospitality.^3^ Our model framework was developed and adapted throughout the epidemic to provide quantitative evidence and epidemiological insights to the UK government. As an update to this work, we show what the impact of the full roadmap would have been in the absence of Delta, and summarise the real-time modelling of policy options that informed the delaying of the final step four.^22–24^ Finally, we assess the potential epidemic magnitude, timing, and main sources of uncertainty after step four under different VE and immune escape assumptions of Delta and the level of transmissibility after NPIs are lifted^25,26^, taking into account recent trends in case incidence and hospitalisations.

## Methods

### Epidemiological model and fitting

We extended a previously described stochastic SARS-CoV-2 transmission model to include vaccination and to capture multiple variants^27^. We used Bayesian methods to fit a single strain transmission model to multiple data sources, including daily deaths, hospital admissions and bed occupancy, serological data, and population-level PCR tests, from each English NHS region up to 8 March 2021. By fitting a piece-wise linear time-varying transmission rate with change points aligned to policy change dates, the model reliably captures the age-specific scale and timing of the first two waves in England^27^. To model vaccine rollout, we assumed an 11-week interval between first and second vaccine doses with the distribution of doses by vaccine type and uptake by age informed by detailed NHS data on vaccine administration. To explicitly model the emergence of Delta, we then fitted a two-strain version of the same model to date from 8 March to 31 July, using information propagated from the first inference step and additionally fitting to the PHE variant and mutation (VAM) dataset. The VAM dataset lists all genotyped or sequenced SARS-CoV-2 cases in England by region and date of specimen. By fitting to the frequency of Delta among Alpha and Delta cases over time, we were able to estimate the transmission advantage of Delta over Alpha.

We then used the fitted two-strain model to project the epidemic trajectory after 19 July under different scenarios. For these forward projections, we accounted for seasonality in SARS-CoV-2 transmission and the impact of school holidays on transmission (for full model description and data sources see appendix 1)^28^.

### Characteristics of the Delta variant

In our model fitting and forward projections, we explored plausible ranges of key epidemiological characteristics of Delta. First, we used estimates of the transmission advantage of Delta over Alpha which were informed by fitting the two-strain model to VAM data. Second, we allowed for imperfect cross-immunity between Alpha and Delta: infection with a pre-Delta variant (e.g. Alpha) provides only 75-100% protection against infection with Delta. Third, we explored central, optimistic, and pessimistic assumptions for VE against Delta, informed by recent studies^13,29,30^. Fourth, we assumed that vaccines induced long-lasting immunity, but examined varying assumptions about the duration of infection-induced immunity: lifelong or average of six- or three-year duration (appendix 1, section 4.4)^31–33^. Finally, we accounted for the increased severity of Delta relative to Alpha by assuming a 1·85-fold increased risk of hospitalisation^13^.

### Assessing the impact of Delta on the roadmap

We explored counterfactual scenarios of the impact of the roadmap on the epidemic trajectory in the presence and absence of Delta. We compared the projected number of infections, hospital admissions, and deaths for scenarios with and without Delta, varying cross-protection, VE, and waning immunity as described above. To capture the easing of restrictions at step four, we sampled from a range of values for the reproduction number *R*, the average number of secondary infections generated by one case, in the absence of natural- and vaccine-induced immunity (*R*_*excl_immunity*_) that could occur at that stage. Our central scenario assumes contacts increase gradually over an 11-week period after step 4 to a maximum of 40% (low), 70% (central), or 100% (high) above contact rates estimated for the period step three was in place^34^. Finally, we assessed the impact of delaying step four, planned initially for “not before” 21 June, until 19 July.

### Sensitivity Analysis

To understand the main drivers of uncertainty of the magnitude of the third wave, in the forward projections we systematically varied i) VE against Delta; ii) cross-protection against Delta from prior infection with non-Delta variants; iii) the duration of natural immunity; and iv) the level of transmissibility after step four (appendix 1, section 5.4). We also assessed a further counterfactual scenario where contacts increased to maximum levels immediately after the step 4 date, rather than gradually as in the central scenario.

### Ethics

Ethics permission was sought for the study via Imperial College London’s standard ethical review processes and was approved by the College’s Research Governance and Integrity Team (ICREC reference: 21IC6945).

## Results

The model reproduces national (Figure 1) and regional (appendix 1, section 7.1) trends in the SARS-CoV-2 epidemic in England well, including the emergence and eventual dominance of the Delta variant (Figure 1C,F) and the July dip in case numbers, hospitalisations (Figure 1A,B) and hospital deaths (Figure 1D,E). Figure 2 shows estimates of how the reproduction number and population immunity changed over time since December 2020. Overall, while *R*_*excl_immunity*_ increased markedly from March to July, as a result of the relaxation of lockdown and then the importation of the Delta variant, the rapid rollout of vaccines progressively increases the gap between *R*_*excl_immunity*_ and the effective reproduction number (*R*_*t*_^*eff*^, which accounts for the impact of natural- and vaccine-induced immunity on transmission). The Christmas school holidays and accompanying near-lockdown social distancing followed by the third national lockdown in January successfully brought *R*_*t*_^*eff*^ below the critical threshold of one. This period coincided with a rapid expansion of the national vaccination programme. By 8 March (step one of the roadmap) when educational institutions re-opened, 43% and 2% of eligible adults (>18 years) had received their first and second vaccine dose respectively^4^. Cases, hospitalisations, and deaths continued to decline and remained at low levels even after schools re-opened (Figure 1). Although we estimated a slight increase in *R*_*t*_^*eff*^ after step one, Easter holidays and the roll-out of vaccination maintained *R*_*t*_^*eff*^ below one (Figure 1A) when non-essential retail opened (61% and 15% first and second dose coverage respectively by 12 April, step two)^4^. The Delta variant, detected in early April predominantly in London and the North West regions (Figure 1F), was designated as a “variant under investigation” on 15 April after increasing numbers of locally-acquired infections with that variant were detected (Figure 1C)^19^.

**Figure 1:**
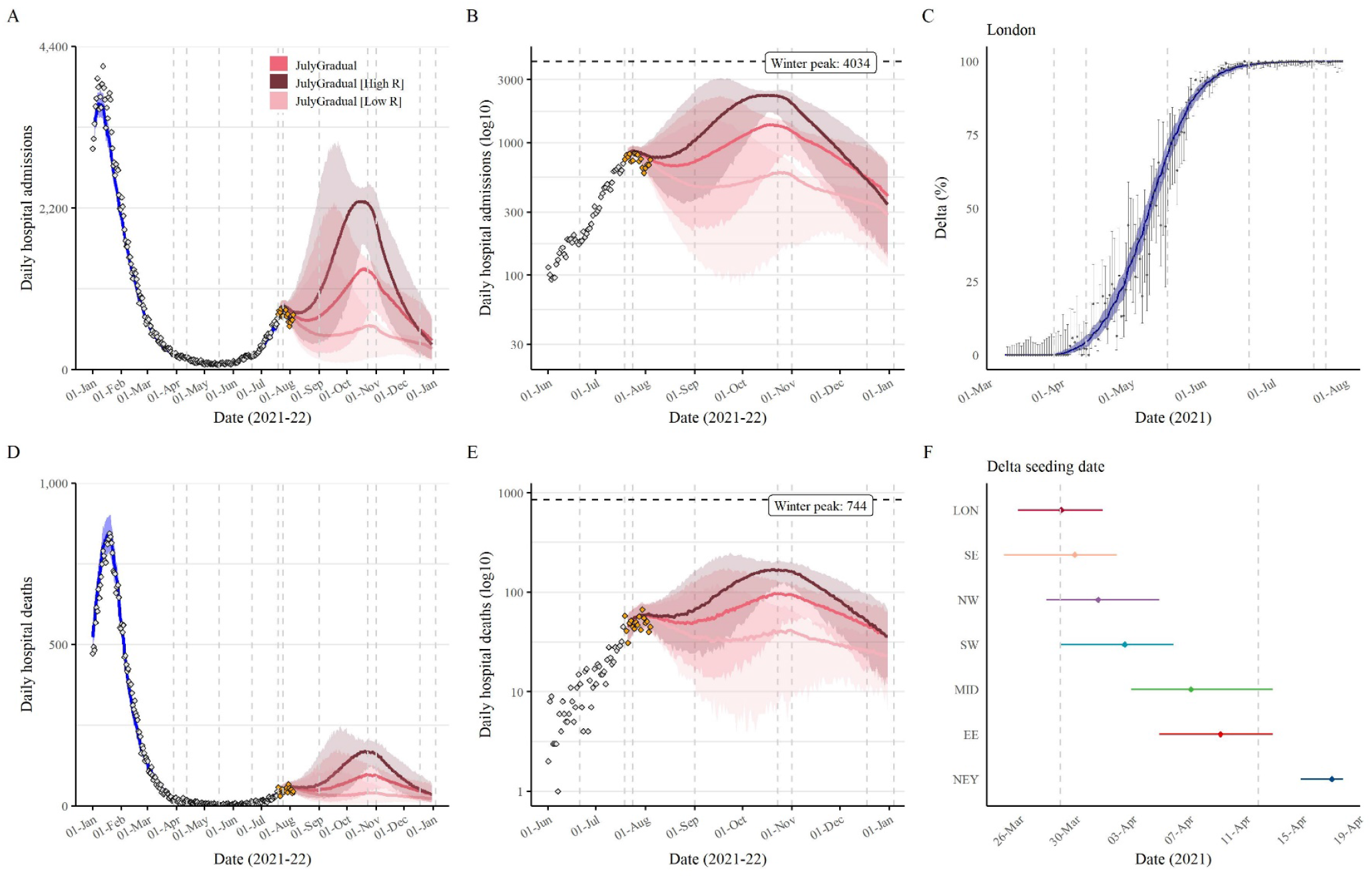
Trajectory of the COVID-19 epidemic in England and the emergence of the Delta variant. (A-B) Observed (grey points) daily hospital admissions and (D-E) daily hospital deaths. The blue line shows the model fit up to 19 July and the red lines the projected admissions and deaths assuming a gradual increase in contacts and a return to high (dark), central (red), or low (light) transmissibility (contact rates) after NPIs are lifted. The orange points in panels B/E show the most recent data which were not fitted to. (C) Model fit to the daily proportion of Delta cases (variant and mutation data, VAM) over time from 8 March-19 July in London as an example. The points show the data, the bar the 95% CI, the blue line the model fit, and the shaded area the 95% CrI (see appendix 1 Figure S17 for other regions). (F) Estimated Delta seeding date by NHS region. Vertical dashed line shows the prior.

**Figure 2:**
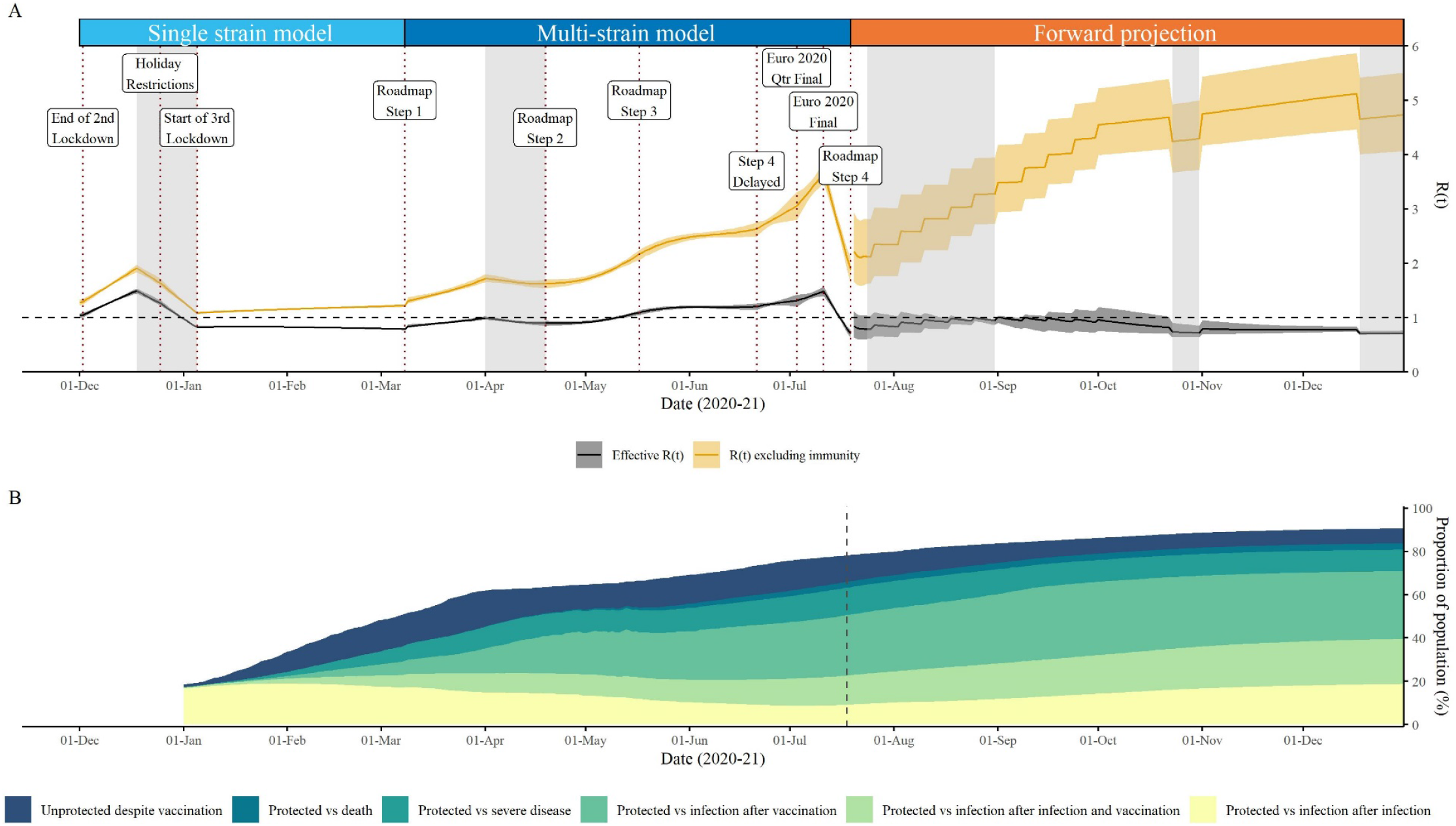
(A) Prevalence weighted (black) effective reproduction number and the reproduction number excluding infection- or vaccine-induced immunity (*Rexcl_immunity*, orange) over time: estimated values from the end of the second national lockdown up to 19 July 2021 (“roadmap step 4” vertical line) and assumed values thereafter. The solid line shows the R(t) median and the shaded area the 95% CrI. The vertical dashed lines show key dates of the roadmap steps and the shaded area the school holidays (appendix 1, sections 4.5 and 5.1.2). The bold dashed lines show when the two-strain model is fitted. (B) Proportion of the English population protected after infection and/or vaccination over time against infection, severe disease, or death. The vertical dashed line denotes the separation between the observed vaccination schedule up to 31 July followed by the simulated schedule assuming a gradual return to central transmissibility after NPIs are lifted.

We estimated that *R*_*t*_^*eff*^ for Alpha remained below one (appendix 1 Figure S17) after step two due to increasing population immunity from vaccination, with uptake increasing to 69% and 39% first and second doses respectively by 17 May (step three, resumption of indoor hospitality and inter-household mixing)^4^. However, after initial seeding between late-March and early April (Figure 1F) the proportion of Delta variant cases was increasing rapidly across all regions in this period (Figure 1C and appendix 1 Figure S17). This rapidly drove *R*_*t*_^*eff*^ above one by mid-May – reflecting the 73% (95%CrI: 68%-79%) transmission advantage of Delta compared with Alpha we estimate (Figure 2A).

The increase in contacts after step three continued to be offset by the increasing vaccine-induced population immunity (Figure 2, 82% and 60% first and second doses respectively by 21 June)^4^. However, the net impact of this differed qualitatively by variant: we estimated that *R*_*t*_^*eff*^ for Alpha remained below one through to mid-July, while *R*_*t*_^*eff*^ for Delta remained above one (appendix 1 Figure S17).

A further sharp increase in *R*_*t*_^*eff*^ was then seen in the first half of July; at the time, we and other modelling groups advising the UK government concluded this was a belated result of step 3, but the rapid drop following 11 July (and other data indicating a sex imbalance in case incidence)^35^, suggest that the increase in transmission rates seen in that period resulted from a transient increase in population contact rates, particularly in young men, likely associated with the Euros football tournament. At the time of writing, that decline has reversed, with a gradual increase in case incidence being seen in the first two weeks of August.^21^

Figure 2 shows that despite high vaccine coverage in adults, sufficient population susceptibility remains for a third wave to occur as contact rates rise. The proportion protected against infection after vaccination against Delta is substantially lower than for Alpha, and most individuals <18 years have neither been vaccinated or infected. In this context, Delta’s high transmissibility means that population immunity is insufficient to keep *R*_*t*_^*eff*^ below one.

With the emergence of Delta, our projections show that had step four occurred on 21 June as initially planned, it may have caused a substantial third wave of hospitalisations and deaths, but with wide uncertainty regarding the magnitude of that wave (Figure 3). Projected total deaths between 21 June 2021 and 1 June 2022 ranged from 11,400 (95%CrI: 6,600-17,600) in the most optimistic (high VE, 40% increase in contacts) scenario to 40,600 (95%CrI: 31,700-50,200) in the most pessimistic (low VE, 100% increase in contacts) scenario (appendix 1, sections 3.1 and 4.4).

**Figure 3:**
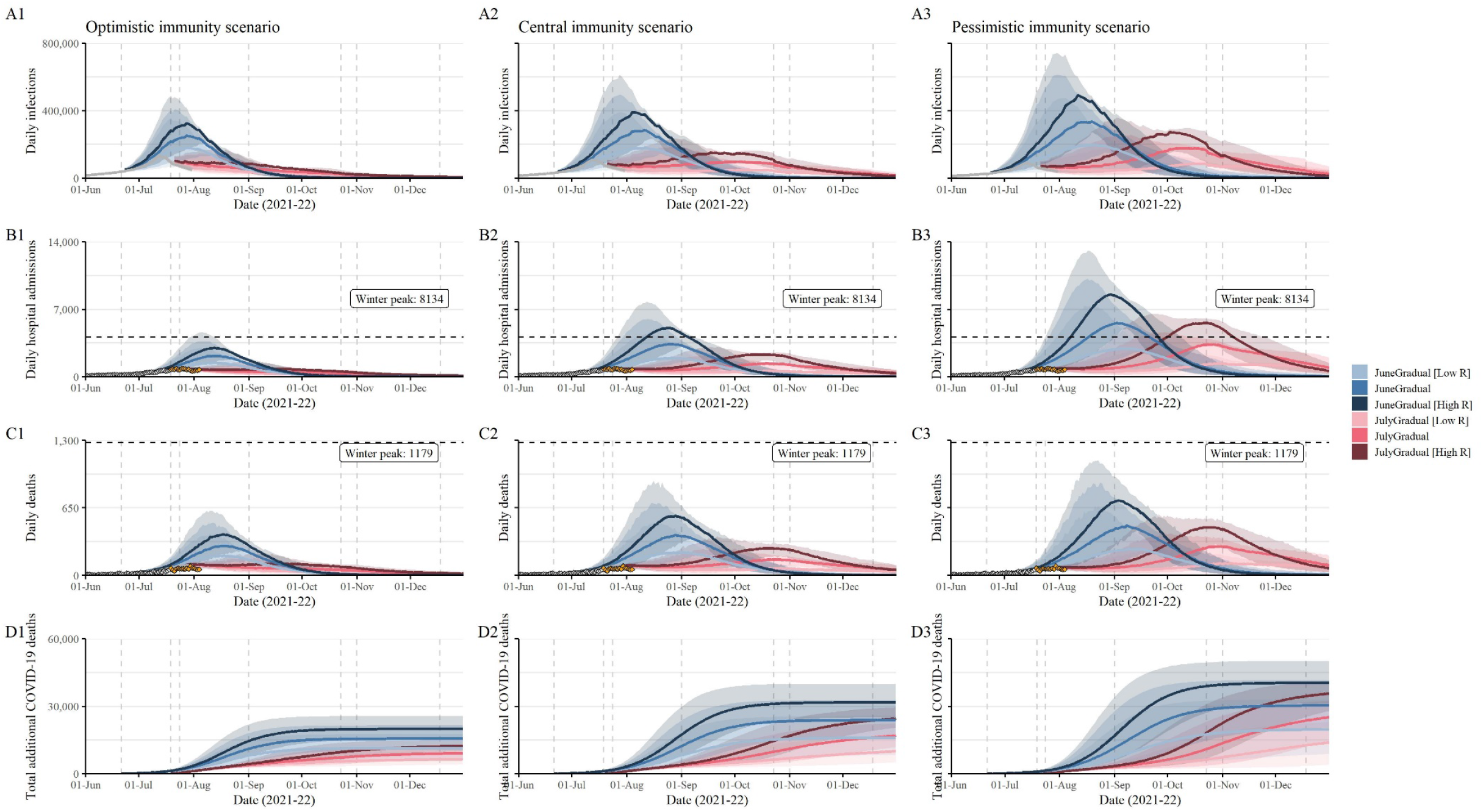
England COVID-19 daily infections (A1-3), hospital admissions (B1-3), deaths (C1-3), and cumulative deaths between 21 June 2021 and 1 Jan 2022 (D1-3) assuming all remaining NPIs were lifted on 21 June (blue) or 19 July (red) with a gradual increase in contacts over 11 weeks thereafter, and a return to low (light colour), central (medium colour), or high (dark colour) transmissibility (contact rates). The grey points show the fitted data and orange the most recent trends (not fitted). Each column shows projections assuming Delta with (A1-A1) optimistic; (A2-D2) central; and (A3-D3) pessimistic vaccine effectiveness, cross-protection, and waning immunity assumptions (appendix 1, section 3.1 and 4.4).

We found that delaying step four until 19 July was beneficial for all scenarios considered, with a much smaller projected wave, although still highly uncertain and sensitive to assumptions around VE against Delta and mixing after NPIs are lifted (Figure 3). The delay allowed the distribution of an additional 2.8 million and 3.8 million first and second doses respectively between 21 June and 19 July^4^, reducing the projected future peak of daily hospital admissions by three-fold from 3,400 (95%CrI: 1,300-4,400) to 1,400 (95%CrI: 700-1,500) in the central scenario of gradually increasing contact rates. It also reduced the total predicted deaths between 21 June 2021 and 1 June 2022 by approximately 15% (appendix 1 Table S16). Had contact rates increased more abruptly after step 4, the reductions in peak hospital admissions and deaths caused by delaying until 19 July would have been substantially greater (see appendix 1, section 7.2). However, trends in cases, hospital admission and death data in the months up to mid-August 2021 suggest that contacts have increased gradually since step 4, in line with the assumptions of our central scenario (Figure 1B,E).

In sensitivity analyses, we found that VE was the main driver of uncertainty of the magnitude of the third wave, followed by the level of mixing after NPIs are lifted, the level of cross-immunity, and waning immunity. The rate at which prior immunity waned had a greater impact on expected total infections but only minimally impacted expected hospitalisations or deaths (Figure 3 and appendix 1, section 7.3).

We project future deaths predominantly in fully vaccinated individuals 75+ years of age, given the high vaccine uptake in the most vulnerable age groups. We also anticipate a substantial number of deaths amongst fully vaccinated adults aged 50 – 74 years (appendix 1, Figure S25).

## Discussion

Mathematical models are valuable tools to inform the design, monitoring, and evaluation of vaccination programmes^36^. In this study, we retrospectively assessed England’s “roadmap out of lockdown”. By extending our evidence synthesis framework to account for vaccinations and multiple variants, we reliably captured the past epidemic and quantified the impact of each phase of lifting interventions and the emergence of the Delta variant on transmission^27^.

We show that the roadmap was successful in mitigating the increase in mixing due to lifting NPIs, by increasing population-level immunity through the mass vaccination programme. Our projections show that in the absence of the Delta variant, lifting NPIs at the planned earliest date of the final step (21 June) would not have resulted in a substantial third wave (appendix 1 Figure S24). This emphasises the importance of carefully aligning the lifting of interventions with immunity levels in the population. Israel rapidly vaccinated a high proportion of their population throughout their re-opening plan, and then implemented vaccination “passes” for entry to high-risk settings^37,38^. Conversely, case incidence and hospitalisations are currently surging in several US states, due to restrictions being released when vaccination coverage was too low to give sufficient population immunity^39^.

Our analyses emphasise the significant impact that the emergence of Delta had on the planned roadmap (Figure 2 and Figure 3). Similar to previous studies, we estimated that Delta has an average 73% (95%CrI: 68%-79%) transmissibility advantage over Alpha^10^. Together with reductions in VE for Delta, this drove a rapid increase in cases and hospitalisations from mid-May that was not offset by the vaccination programme^16,17^. A similar displacement of Alpha and a surge in cases was observed in India where Delta was first detected^40^. The dominance of Delta has now led to a tightening of restrictions in many countries, including in Israel, the country with the fastest vaccination rollout^41^.

Across all the scenarios we examined, delaying step four until 19 July was beneficial (Figure 3) as it allowed more individuals to be vaccinated. In the central scenario, this delayed and reduced the peak of hospitalisations by three-fold and reduced total deaths between June 2021-2022 by approximately 15%.

We project a substantial autumn wave of transmission, but with large uncertainty around the resulting peak hospitalisations and total deaths. This uncertainty is driven by imperfect knowledge of VE against Delta (appendix 1, Figure S3) and thus the overall level of population immunity (accounting for waning and imperfect cross-protection – see appendix 1, section 7.3). At high vaccine coverage, even a difference of 98% vs 95% in VE against mortality translates to a doubling of projected deaths.^42^

Furthermore, the transmission intensity (*R*_*t*_^*eff*^) in the coming months will depend on and how high and how quickly population contact rates will increase, the continued use of face coverings^43^, social distancing, and adherence to case isolation^44,45^. The rapid exponential growth in case incidence in the first half of July illustrates the high transmission levels that could be reached if contact rates approach pre-pandemic levels in the coming months. Fortunately, that increase in contact rates, likely caused by the Euro 2020 football tournament^35^, proved transient, and was then followed by the synchronous self-isolation of contacts alerted through NHS test and trace^46^ and the start of school holidays a week later – driving transmission down for the last two weeks of July. Polls showed that 57% of UK adults were worried about the removal of legal restrictions and 66% would continue to wear a face covering after 19 July^47^. The average number of contacts is still much lower than pre-pandemic levels^48^, and our central scenario which assumes a gradual increase in contact rates most closely reflects current trends (Figure 1). We estimate that this slow increase in contact rates after 19 July will dampen the peak of hospitalisations and total deaths (Figure 3 and appendix 1 Figure S24) compared to an abrupt increase.

We have focused on deaths and hospital admissions as primary outcomes in our analysis, given the impact of hospitalisation on NHS capacity. However, an estimated 1.5% of individuals in the UK were experiencing symptoms of long-COVID (symptoms lasting more than 4 weeks) on 4 July 2021^49^. Although our estimates of infections and cases over time may capture this wider burden of disease,^50^ we did not explicitly quantify this. Overall infection levels also determine the risks of new VOCs emerging within the UK with the risk increasing with transmission levels.

Independently of VOCs emerging, the duration of infection- and vaccine-induced immunity for all SARS-CoV-2 lineages remains a key unknown and will determine long-term transmission dynamics. Better characterising the duration of immunity against both infection and severe disease will be important for informing vaccination boosterC programmes.

Our analysis has a number of limitations. We did not consider re-introduction of NPIs, vaccination of <18 year-olds, nor booster doses in our projections. These measures, if introduced, may partially mitigate the third wave. Vaccination of 16-17 year olds is planned to start in late August, and clinically vulnerable 12-15 year-olds are also eligible for vaccination^51,52^. However, given the age-profile of projected hospitalisations and deaths (appendix 1, Figure S25) we anticipate expansion of eligibility may only have a moderate impact. Although we modelled heterogeneity by age in mixing patterns and in vaccine uptake, we did not explicitly model other factors (e.g. by occupation, sociodemographic, and ethnic groups^53,54^) which may affect both the risk of infection and vaccine uptake. Vaccine uptake was also assumed to be independent of mixing patterns or viral transmission. Groups of individuals who are both at high risk of infection and less likely to take the vaccine may lead to continued outbreaks amongst vulnerable populations and reduce the overall impact of vaccination. Although we explored the impact of waning of natural immunity, here we assumed vaccine-induced immunity did not wane over the timescales we considered in this paper. Our analysis focused only on outcomes directly related to COVID-19: we did not consider the impact on health services, other diseases, mental health, or the economic impact of measures.

In summary, our study shows how the phased lifting of NPIs in England, coordinated with vaccine roll-out, has been largely successful at keeping hospitalisation and deaths at low levels since March 2021. However, our projections show that the high transmissibility of Delta, imperfect VE, and future increases in contact rates are likely to lead to a substantial wave of transmission in the autumn, albeit of highly uncertain magnitude. Overall, our analysis highlights the clear benefit of early and accessible national vaccination programmes which allow population immunity to increase to high levels before NPIs are lifted. Finally, we have showed that vaccination alone in the absence of NPIs may not be sufficient to control Delta even with high vaccination coverage. We quantified how the emergence of Delta affected the progress of the roadmap and the benefit of delaying step four of that roadmap by one month. The experience of Delta highlights the threat posed by any future VOCs and underscores the need for global collaborative efforts to control transmission and mitigate the risk of emergence of new VOCs through equitable global access to vaccines.

## Data Availability

All data files and source code are available here: https://github.com/mrc-ide/sarscov2-roadmap-england

## Acknowledgements

We thank all the colleagues at Public Health England (PHE) and frontline health professionals who not only have driven and continue to drive the daily response to the epidemic but also provided the necessary data to inform this study. This work would not have been possible without the dedication and expertise of said colleagues and professionals. The use of pillar-2 PCR testing data, vaccination data, and the variant and mutation data was made possible thanks to PHE colleagues, and we extend our thanks to N. Gent and A. Charlett for facilitation and insights into these data. The use of serological data was made possible by colleagues at PHE Porton Down, Colindale, and the NHS Blood Transfusion Service. We are particularly grateful to G. Amirthalingam and N. Andrews for helpful discussions around these data. We thank all the REal-time Assessment of Community Transmission (REACT) Study investigators for sharing PCR prevalence data. We also thank the entire Imperial College London Covid-19 Response Team for support and feedback throughout. The views expressed are those of the authors and not necessarily those of the U.K. Department of Health and Social Care, the National Health Service, the National Institute for Health Research (NIHR), PHE, UK MRC, UKRI, or European Union.

## Author contributions

Conceptualization – Ideas; formulation or evolution of overarching research goals and aims: RS, PNPG, ESK, LKW, NI, EMV, ACG, NMF, MB, and AC; Data curation – Management activities to annotate (produce metadata), scrub data and maintain research data: KAMG, WH, BAD, NI, RGF, PNPG, and ESK; Formal analysis – Application of statistical, mathematical, computational, or other formal techniques to analyse or synthesize study data: RS, LKW, PNPG, ESK, KAMG, MB, and AC; Funding acquisition: MB, NMF, and AC; Investigation – Conducting a research and investigation process, specifically performing the experiments, or data/evidence collection: RS, LKW, PNPG, ESK, TR, MB, and AC; Methodology – Development or design of methodology; creation of models: RS, LKW, PNPG, ESK, MB, NMF, and AC; Project administration – Management and coordination responsibility for the research activity planning and execution: NI, RS, NMF, MB, and AC; Resources – Provision of computing resources, or other analysis tools: JAL and RGF; Software – Programming, software development; designing computer programs; implementation of the computer code and supporting algorithms; testing of existing code components: RS, LKW, PNPG, ESK, TR, WH, JAL, RGF, MB, and AC; Supervision – Oversight and leadership responsibility for the research activity planning and execution, including mentorship external to the core team: NMF, MB, and AC; Validation – Verification, whether as a part of the activity or separate, of the overall replication/reproducibility of results/experiments and other research outputs: RS, LKW, NI, PNPG, ESK, TR, NMF, MB, and AC; Visualization – Preparation, creation and/or presentation of the published work, specifically visualization/data presentation: RS, LKW, PNPG, TR, and WH; Writing – original draft – Preparation, creation and/or presentation of the published work, specifically writing the initial draft: NI, TR, RS, ESK, LKW, MB, and AC; Writing – review & editing – Preparation, creation and/or presentation of the published work by those from the original research group, specifically critical review, commentary or revision – including pre- or post-publication stages: NI, RS, LKW, TR, ESK, PNPG, DTK, KAMG, JAL, ACG, MB, AC, and NMF.

## Supplementary Materials

Appendix 1

