## Appendix 1 for "Non-pharmaceutical interventions, vaccination and the Delta variant: epidemiological insights from modelling England’s COVID-19 roadmap out of lockdown"

### Supplementary Material

Symbol Glossary 67

List of Figures 69

List of Tables 70

#### 1 Model Description and Definitions

In this section we provide a brief description to our model along with key definitions. Full details about the fitting procedure, parameter assumptions, and model equations are provided in Section 4.

##### 1.1 Overview of the model

We used a discrete-time stochastic compartmental model of SARS-CoV-2 transmission, illustrated in Figure S1, which has previously been described in detail in Knock et al. (2021) [1]. In short, the model is an extended SEIR-type model, stratified into 16 five-year age groups (0-4, 5-9, . . . , 75-79), 80+, a group of care home residents (CHR) and a group of care home workers (CHW). Mixing between these groups is informed by survey data [2]. Upon infection with SARS-CoV-2, individuals enter an exposed compartment, before becoming infectious. A proportion of infectious individuals are assumed to develop symptoms, while the rest remain asymptomatic. All asymptomatic cases and a fraction of symptomatic cases recover naturally, while the rest of the symptomatic cases develop severe disease requiring hospitalisation. Of these, a proportion die at home, while the remainder are admitted to hospital. Hospital pathways are described in detail, with patients being either triaged before ICU admission, then admitted to ICU, before being transferred into general wards for stepdown recovery, or remaining in general beds throughout. Hospitalised cases are either confirmed as SARS-CoV-2 cases upon admission or may be tested and confirmed later during their stay.

Each compartment in the model is further stratified to account for vaccination status. We used four vaccination strata (Table S2 and Figure S1), which describe the recommended two-dose vaccination regimen (common to the three double-dose vaccines currently licensed and available for use in England: Oxford-AstraZeneca ChAdOx1 nCoV-19 (AZD1222) [3], Pfizer-BioNTech COVID-19 Vaccine BNT162b2 [4], and Moderna mRNA-1273 [5], henceforth referred to as AZ, PF, and Mod respectively), capturing a delay between receiving a dose and onset of dose-specific effectiveness.

As presented in Knock et al. (2021) [1], the model is utilised in two stages, an initial model fitting stage, to build posterior estimations of model parameters fitting to multiple epidemiological data streams, followed by a forecasting ('forward projection') stage, whereby the posterior estimates inform medium- and long-term projections for the pandemic trajectory as well as counterfactual "what if" scenarios. PCR and serology status are modelled with parallel flows.

The model has since been extended to include the spread of variants of concern (VOC). In the context of this paper, we consider Alpha (B.1.1.7) coexisting with the Delta variant (B.1.617.2). All references to 'Alpha' here refer to the Alpha variant and all other previously circulating variants. Before the emergence of Delta, we fit a one-variant model, and then switch to a two-variant model on 8 March 2021, where Delta is seeded at a region-specific date determined by the model fit. Additionally we added a second parallel serology flow, which allows us to fit to samples using two different serology assays, with different durations of seropositivity.

We now include waning of infection-induced immunity in the model. Individuals who have recovered from COVID-19 infection are protected against reinfection with the same variant of SARS-CoV-2 for an exponentially distributed duration with mean 6 years, after which they move back to the susceptible compartment. Other waning rates are considered within sensitivity analyses (Table S7). Further, we model asymmetrical cross-immunity between SARS-CoV-2 variants (Section 2.5). We will use the term 'susceptible' only to refer to individuals in 'S', whereas 'uninfected' will refer to those in 'S' or 'R'.

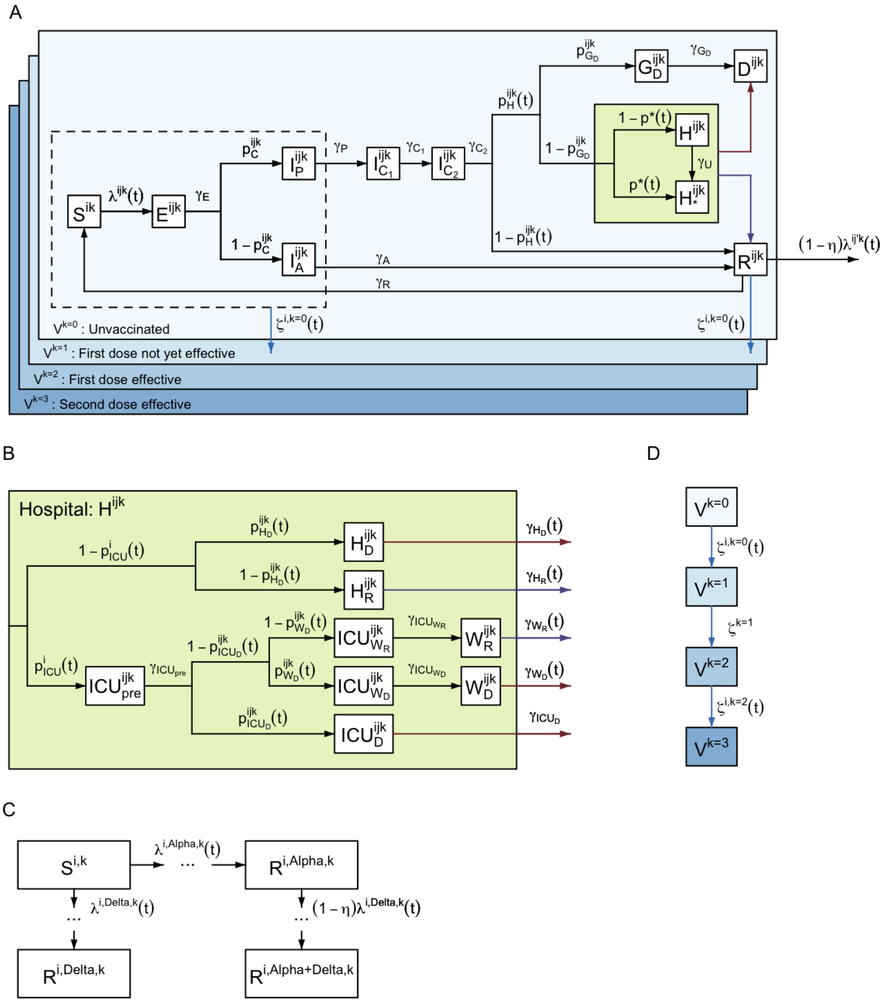

**Figure S1:** Model structure flow diagram with rates of transition between states. (A) Extended SEIR transmission model flow diagram overview. (B) Hospital flow diagram. (C) Vaccination flow diagram. (D) Multi-variant flow diagram showing possible infection with $Alpha$, $Delta$, or both in turn ($Alpha$➔$Delta$). All variables and parameters defined in tables S5, S6, and S7. Superscripts refer to the age or care home group ($i \in [0-4], [5-9], \ldots, [75-79], [80+], CHW, CHR$), variant ($j = Alpha, Delta$, or $Alpha$➔$Delta$), and vaccination stratum ($k = 0, \ldots, 3$).

##### 1.2 Reproduction number

We use two definitions of the reproduction number throughout. We denote $R^j(t)$ as the reproduction number for variant $j$ ($j = Alpha, Delta$) in the absence of immunity at time $t$. This is defined as the average number of secondary infections that an individual infected at time $t$ with variant $j$ would generate in an entirely susceptible and unvaccinated population. In contrast, the effective reproduction number, $R_e^j(t)$, for variant $j$

at time $t$ is the number of secondary infections in the actual uninfected population, accounting for immunity (natural and vaccine-induced). Hence, by definition, $R_e^j(t) \leq R^j(t)$.

##### 1.3 Fitting to data

The model is fitted to multiple data streams from each NHS region in England, as described in Knock et al. (2021), this is summarised in Table S1.

#### 2 The Delta Variant

The model switches to a two-variant set-up at 8 March 2021 to capture the emergence and spread of Delta, due to its proliferation to being the established dominant variant in all NHS regions of England. Key differences between the modelled variants are summarised in the next subsections, these are:

1. Date of introduction.
2. Transmissibility of Delta compared to Alpha.
3. Vaccine effectiveness against Delta.
4. Severity of Delta compared to Alpha.
5. Cross immunity conferred by prior infection with Alpha.

##### 2.1 Transition from single to two-variant model

To account for the emergence of the Delta variant while keeping the fitting process as efficient as possible, our dynamic transmission model is calibrated in two parts. We first run the inference using a single variant model until 8 March 2021. While the model only considers one variant in this part, some of the fitted epidemiological parameters (such as the transmission parameters or the probability of dying in hospitals) are time-varying. These time-varying parameters capture the trends observed in the data following changes in behaviour, in treatment practice, or the replacement by the Alpha variant of previously circulating variants. At 8 March 2021, we switch to a two-variant model and propagate the information from the first inference part in two ways. At the connection timepoint, models should be equivalent as there are no individuals infected with the Delta variant at this date. Infections with the Delta variant are then introduced on a region-specific date ($t_{Delta}$) determined by the model fits, 20 cases per million inhabitants are then seeded to the regions, distributed across the 7 days immediately following the initial seeding date $t_{Delta}$, and distributed evenly across the regional 5-year age groups as described by ONS population estimates [9].

We propagate the sample of filtered end model states on 8 March (obtained with our particle MCMC algorithm [1]) and use it as the initial distribution of the two-variant model. All compartments associated with being infected with the Delta variant are set to 0. Second, we propagate the information from the fitted posterior distributions of parameters common between parts 1 and 2. We fit a functional form (Table S8) to each of the parameters' marginal posterior distribution obtained in part 1 using maximum likelihood. These distributions are then used as priors for part 2.

The two-variant model was fitted to the same data streams as in Table S1, as well as the variant and mutation (VAM) dataset, which contains the daily number of variant tests by NHS region identified as Delta and as identified as Alpha (or other non-Delta circulating variant).

| Data | Description | Source | Reference |
|---|---|---|---|
| Hospital deaths | Daily number of COVID-19 deaths reported by NHS England within 28 days of a positive result | PHE | These data underlie the Gov.uk dashboard data [6] |
| Care home deaths | Daily number of deaths with COVID-19 mentioned as a cause on the death certificate and "care home", "hospice" or "other institution" as the place of death | ONS | These data underlie the Gov.uk dashboard data [6] |
| Community deaths | Daily number of deaths with COVID-19 mentioned as a cause on the death certificate and any place of death that is not one of "hospital", "care home", "hospice" or "other institution" | ONS | These data underlie the Gov.uk dashboard data [6] |
| ICU occupancy | Daily number of confirmed COVID-19 patients in ICU | Gov.uk Dashboard | [6] |
| General bed occupancy | Daily number of confirmed COVID-19 patients in non-ICU beds | Gov.uk Dashboard | [6] |
| Admissions | Daily number of confirmed COVID-19 patients admitted to hospital | Gov.uk Dashboard | [6] |
| Pillar 2 testing | Daily number of positive (cases) and negative PCR test results for individuals aged 25 or over | PHE | These data underlie the Gov.uk dashboard data [6] |
| REACT-1 testing | Daily number of positive and negative PCR test results | REACT | [7] |
| Serology | Serology survey conducted on blood donors aged 15-65. Results using the EuroImmun and Roche N assays are used, fitting to each assay separately. EuroImmun results are only used up to (and including) 14th January 2020, | PHE | These data are collected as part of [8]. |
| Vaccinations by age | Daily number of first and second-vaccine doses - reported in 5-year age groups | PHE | [6] |

**Table S1:** Data sources and definitions.

##### 2.2 Transmissibility compared to Alpha

We fit region-specific transmission advantages, $\sigma$, of the Delta variant compared to Alpha. These are initialised with uniform priors between 0 and 3 (Table S8), with an initial value of 1.

##### 2.3 Vaccine effectiveness

On top of the innate transmission advantage considered for Delta, transmission will also be boosted due to reduced vaccine effectiveness against Delta. Both the transmission advantage, $\sigma$, and variant-specific vaccine effectiveness are captured in the force of infection terms presented in Section 4.2. For full details on vaccine effectiveness against Delta, see Section 3 and in particular Table S3.

##### 2.4 Increased severity

Following Sheikh et al. (2021) [10], we additionally multiply the relative probability of hospitalisation by 1.85 for those infected by Delta.

##### 2.5 Protection from previous infection

The level of cross-protection from prior infection with non-Delta variants is difficult to quantify but in-vitro neutralisation studies found Delta was less susceptible to antibodies from previous infections [11]. We model asymmetric cross immunity between the two variants and assume that infection with Delta confers perfect immunity to infection with Alpha, whilst infection with Alpha is only partially protective against infection with Delta (see Table S7). In addition, for individuals infected by Delta following on from an infection with Alpha ($I_{C_2}^{i,Alpha \to Delta,k}$), we assume that, if the second infection is symptomatic, the probability of hospitalisation is reduced compared to individuals with no prior infection history. We consider this factor reduction equivalent to the conditional effectiveness against severe disease dependent on being symptomatic afforded by one dose of PF against Delta ($e_{SD|sympt}$ in Table S4 below). Probability of infection or hospitalisation by either variant 'resets' to base assumptions once an individual's acquired immunity wanes and they re-enter the susceptible compartment, $S^{i,k}$.

#### 3 Vaccination

The Medicines and Healthcare Products Regulatory Agency issued temporary authorisation grants for both the Oxford-AstraZeneca (AZ) and the Pfizer-BioNTech (PF) vaccines in December 2020 [3, 4], and approved the Moderna (Mod) vaccine shortly after in January 2021 [5]. All three vaccines require two doses to be administered, with increasing levels of vaccine effectiveness seen after each dose. As such, our model considers four distinct vaccination strata ($V_k$, for $k \in \{0,1,2,3\}$) representing the four stages of vaccine effectiveness available, as detailed in Table S2 and illustrated in Figure S2. We model the UK vaccination strategy, i.e. to delay the second dose to, on average, 11 weeks after the first dose [12].

| Vaccination stratum name | Number of doses | Vaccine effectiveness for that group | Mean duration | References |
|---|---|---|---|---|
| $V_0$ | 0 | None | Determined by first dose vaccine roll-out | Section 3.4 |
| $V_1$ | 1 | None | 3 weeks | [13, 14] |
| $V_2$ | 1 | Full first dose effectiveness | In model-fit stage, determined by second dose vaccine roll-out; in simulation-stage, 8 weeks | [12, 13, 14] |
| $V_3$ | 2 | Full second dose effectiveness | Infinite | Section 3.4 |

**Table S2:** Vaccination strata considered for individuals with the mean duration an individual spends in each strata and vaccine effectiveness at each stage.

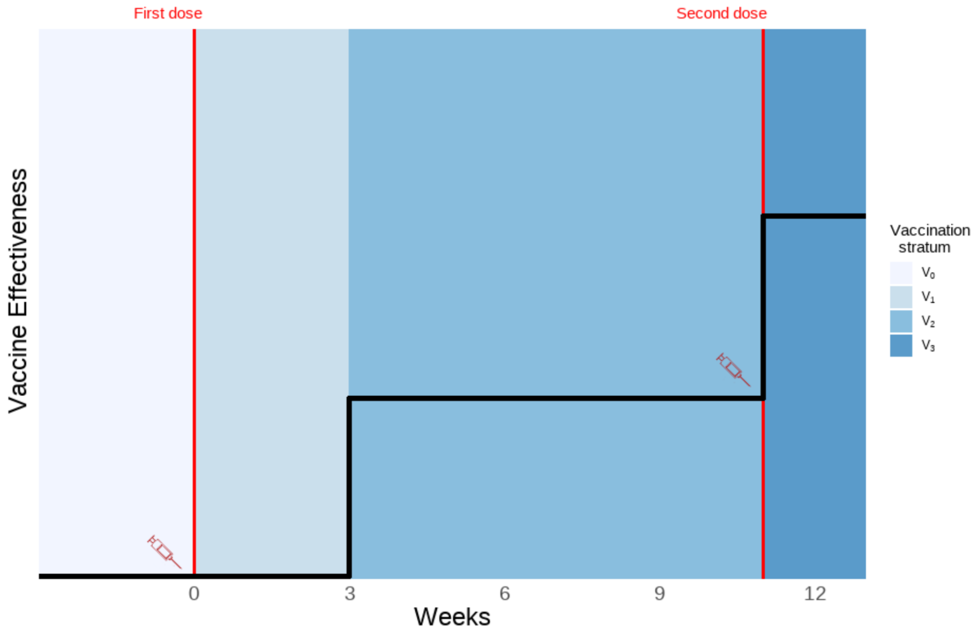

**Figure S2:** Vaccination strata duration and associated illustrative vaccine effectiveness. Red lines depict points at which a vaccine dose is administered. y-axis is an illustration of increasing vaccine effectiveness. Vaccination strata are defined in Table S2.

Individuals in our model move out of an unvaccinated ($V_0$) stratum at a rate determined by the age-specific vaccine roll-out and prioritisation strategy adopted by the UK government (Section 3.4). We only allow vaccination of individuals who are not symptomatic and not hospitalised, i.e. only individuals in the following compartments can be vaccinated: susceptible ($S$), exposed ($E$), infected asymptomatic ($I_A$), infected pre-symptomatic ($I_P$) or recovered ($R$). Other compartments are also stratified by our four vaccination strata but for those there is no movement between vaccine strata ($V_k$).

Phase 2 trials for the AZ and PF vaccines suggested substantial increases in immunogenicity started approximately two to three weeks after receiving the first dose [13, 14]. We therefore assumed a 21-day delay between first dose injection and onset of dose-specific effectiveness. In our model, after receiving their first dose, individuals remain in $V_1$ for 21 days on average, during which the vaccine offers no protection. They then move on to the $V_2$ strata, where they stay for eight weeks on average and are protected with one dose. After this eight week period, individuals move on to the $V_3$ stratum where they are assumed to achieve maximal protection offered by the two doses. This is illustrated in Figure S2. Note that whilst the data shows an average of 10 days between receiving the second dose and it taking full effect, the difference in our model reflects individuals receiving their second dose earlier than 11 weeks after the first. We assume no waning of vaccine-induced immunity on the time horizon of the analysis.

##### 3.1 Vaccine effectiveness

The assumed values for vaccine effectiveness (VE) are derived from both vaccine efficacy measured in clinical trials and vaccine effectiveness studies. Where possible, data from the UK have been used and represent effectiveness of dosing schedules with an 11 week gap between doses. We assumed that there are no significant differences in vaccine effectiveness by age, sex or underlying health conditions [15, 16]. Table S3 summarises our vaccine effectiveness assumptions for the PF, AZ and Mod vaccines. We assume that vaccine protection against symptomatic disease, as determined from the original trials and real-world data, also provides a similar level of protection against infection. We further assume that, in those individuals who do become infected after vaccination, onward transmission is also reduced [17]. Finally, we incorporate a higher overall level of vaccine effectiveness against hospitalisation and against death. Due to a high degree of uncertainty in the literature surrounding the effectiveness against the Delta variant, we adopted central, optimistic, and pessimistic values to be considered through sensitivity analysis. Figure S3 demonstrates our assumed values against the 95% CrI of the associated literature, where UK-based studies are prioritised where available.

| Vaccine effectiveness | Vaccine (dose) | Alpha | Delta (Central) | Delta (Pessimistic/Optimistic) | Informed by |
|---|---|---|---|---|---|
| Against death | AZ (1) | 80% | 80% | 75%/81% | [18, 19] |
| | AZ (2) | 95% | 95% | 95%/95% | [18, 19, 20] |
| | PF (1) | 85% | 85% | 80%/93% | [18, 19] |
| | PF (2) | 95% | 95% | 95%/98% | [18, 19] |
| | Mod | Assume same as PF for 1 and 2 doses | | | |
| Against severe disease | AZ (1) | 80% | 80% | 75%/81% | [21, 22] |
| | AZ (2) | 90% | 90% | 85%/94% | [22, 23, 24], Assumed greater than mild disease. |
| | PF (1) | 85% | 85% | 80%/93% | [23] |
| | PF (2) | 95% | 95% | 90%/98% | [24, 25], Assumed greater than mild disease. |
| | Mod | Assume same as PF for 1 and 2 doses | | | |
| Against mild disease | AZ (1) | 50% | 33% | 20%/45% | [15, 24] |
| | AZ (2) | 74% | 58% | 45%/70% | [15, 26, 27] |
| | PF (1) | 50% | 33% | 20%/45% | [10, 22, 25, 27] |
| | PF (2) | 93% | 85% | 78%/90% | [10, 25, 27, 28] |
| | Mod | Assume same as PF for 1 and 2 doses | | | |
| Against infection | AZ (1) | 50% | 33% | 20%/45% | [10] Assumed same as effectiveness against disease. |
| | AZ (2) | 74% | 58% | 45%/70% | [10] |
| | PF (1) | 50% | 33% | 20%/45% | [10] |
| | PF (2) | 93% | 85% | 78%/90% | [10] |
| | Mod | Assume same as PF for 1 and 2 doses | | | |
| Against infectiousness if infected | AZ (1) | 45% | 40% | 35%/45% | [17] / Assumed |
| | AZ (2) | 45% | 40% | 35%/45% | [17] / Assumed |
| | PF (1) | 45% | 40% | 35%/45% | [17] / Assumed |
| | PF (2) | 45% | 40% | 35%/45% | [17] / Assumed |
| | Mod | Assume same as PF for 1 and 2 doses | | | |

**Table S3:** Vaccine effectiveness assumptions for AstraZeneca (AZ), Pfizer (PF), and Moderna (Mod).

We model cases that require hospitalisation and are hospitalised, as well as cases that require hospitalisation but are not hospitalised, for this reason we refer to vaccine effectiveness against *severe disease* and not *hospitalisation*. Vaccine effectiveness against severe disease, conditional on symptoms, acts on transition to both this compartment of individuals and those admitted to hospital.

We do not model individual vaccines separately, instead vaccine compartments are type-agnostic, and for vaccine effectiveness we compute an age-dependent weighted mean of each vaccine's effectiveness (where weights for a given age group are the proportion of each vaccine type administered to that age group as of 19 July 2021). Whilst we assume vaccine effectiveness does not vary by age, the proportion of each vaccine (PF, AZ or Mod) administered to each age group varied substantially (Figure S4) and vaccine effectiveness varies between vaccines (Table S3), therefore our weighted vaccine-effectiveness varies by age.

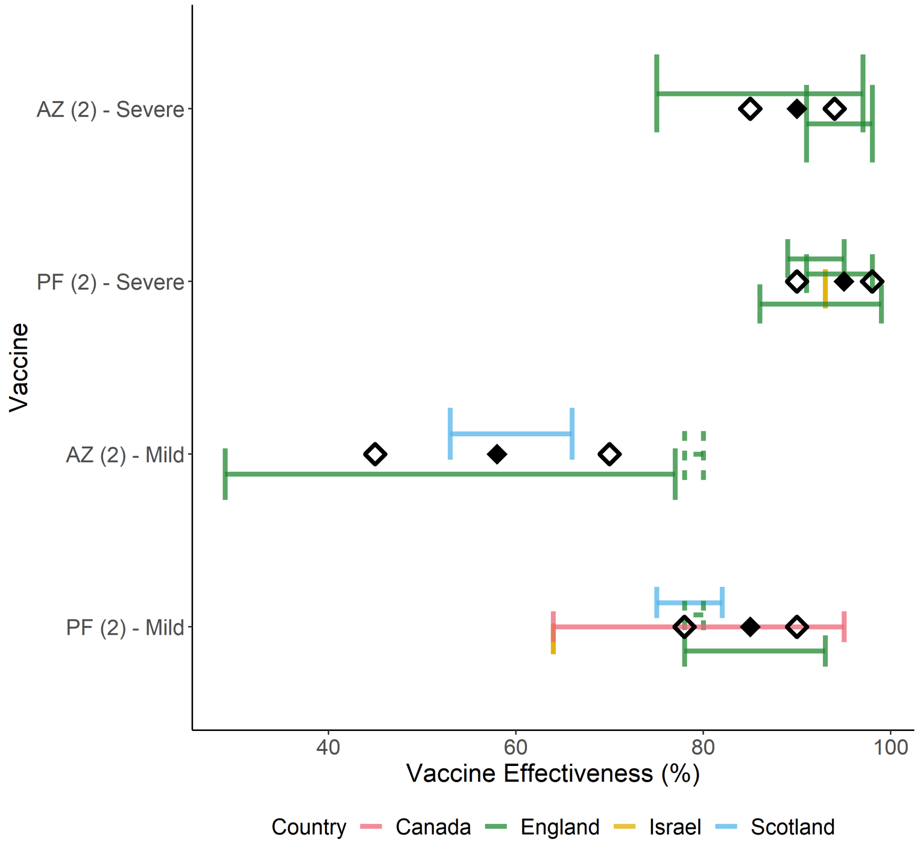

**Figure S3:** Comparison of our chosen Delta-specific VE values for two vaccine doses (Table S3) against associated 95% CrI values from the literature. Estimates taken from studies conducted in Canada [28], England [24, 22, 20, 27], Scotland [10], and Israel [29]. Black diamonds indicate our central assumptions, and hollow diamonds our pessimistic/optimistic assumptions. Dashed lines indicate a study that reported combined AZ/PF effect.

##### 3.2 Conditional dependencies of vaccine-immunity

We present unconditional VE in Table S3 however our model is framed as a compartmental cascade of symptom severity, hence we convert unconditional effectiveness to conditional as detailed in Table S4.

##### 3.3 Vaccine-induced immunity

In the absence of long-term follow-up studies on the duration of COVID-19 vaccine-induced immunity, we assume that IgG and T cell responses will be maintained for longer than the time horizon modelled, and as such do not consider the waning of vaccine-protection [30].

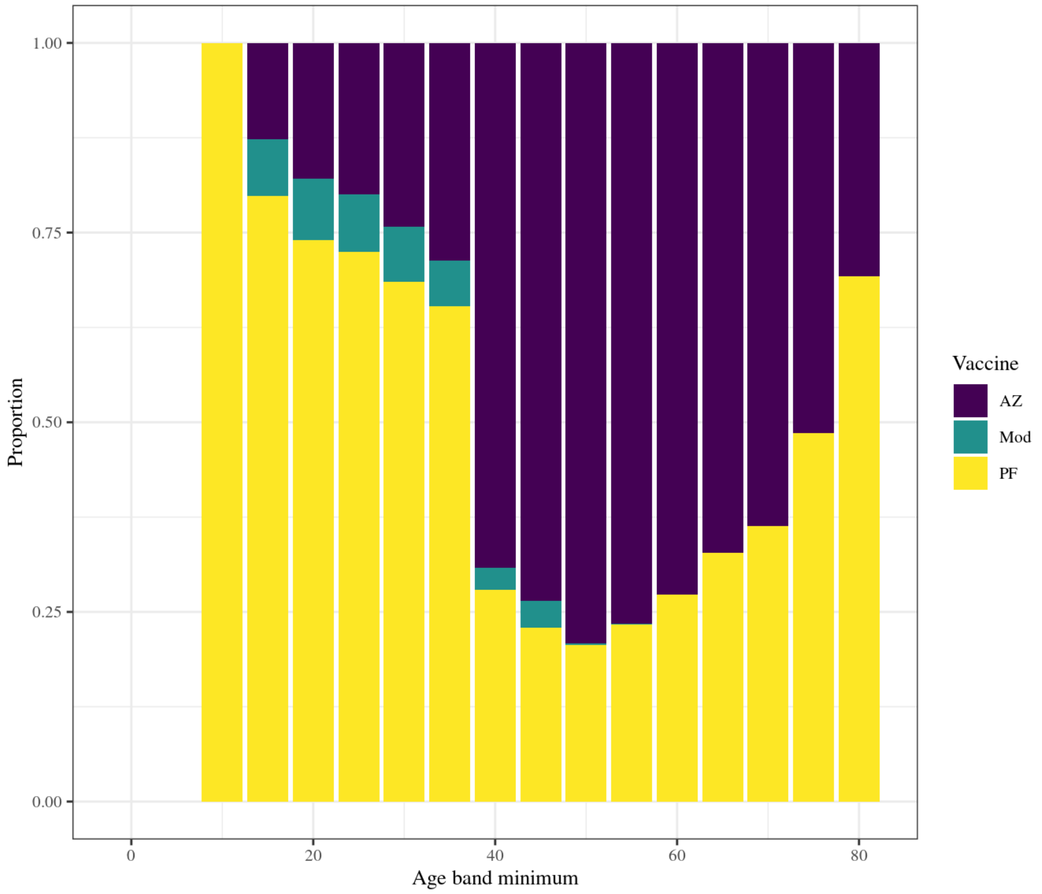

**Figure S4:** Proportion of each vaccine type: (Oxford-AstraZeneca (AZ), Pfizer-BioNTech (PF), Moderna (Mod)) dispensed to each five-year age band as of 31 July 2021. Data taken from PHE Immunisations database for vaccine delivery and ONS population estimates for each age group.

##### 3.4 Vaccine roll-out

The Joint Committee on Vaccination and Immunisation (JCVI) established an ordered list of individuals prioritised for vaccination in the UK, prioritising first care home residents and care home workers, and then other adults by decreasing age and clinical vulnerability [31, 32].

As our model is stratified using 5-year age classes, we model the vaccination of individuals aged 18-19 by assuming the uptake in the 15-19 age group is 2/5 of the uptake in the 20-24 year olds. Children under 18 years are not vaccinated in our model. We assume first and second doses were delivered in England between 8 December 2020 and 19 July 2021 as reported in age-stratified data received from PHE and DHSC via SPI-M (Figure S5) as reported on the COVID-19 dashboard [6]. It is not possible to identify individuals in our CHR and CHW groups from the data however as these are the first two priority groups we assume that all individuals vaccinated under 65 years were carehome workers and all individuals 65 years or over were carehome residents, up until the desired uptake in CHR and CHW is reached.

| VE vs. | Symbol / Calculation |
|---|---|
| Infection | $e_{inf}$ |
| Symptoms | $e_{sympt}$ |
| Severe disease | $e_{SD}$ |
| Death | $e_{death}$ |
| Symptoms given infection | $e_{sympt\|inf} = \dfrac{e_{sympt} - e_{inf}}{1 - e_{inf}}$ |
| Severe disease given symptoms | $e_{SD\|sympt} = \dfrac{e_{SD} - e_{sympt}}{(1 - e_{inf})(1 - e_{sympt\|inf})}$ |
| Death given severe disease | $e_{death\|SD} = \dfrac{e_{death} - e_{SD}}{(1 - e_{inf})(1 - e_{sympt\|inf})(1 - e_{SD\|sympt})}$ |

**Table S4:** Conditional vaccine effectiveness values that we model.

In forward projections we assume an average vaccine dose roll out of 1.9 million doses per week, based on past schedule, split between first and second doses to prioritise the latter, so that individuals having already received one dose receive their second dose approximately 11 weeks after their first. We assumed doses are split between NHS regions in proportion of their population size.

#### 4 Model Fitting

##### 4.1 Model compartments and parameters

In the following, $i$ denotes the age or care home group of individuals ($i = [0,5), [5,10), \ldots, [75-80), [80+)$, $CHW, CHR$), and $j$ denotes their variant status ($j = Alpha$ for infection with Alpha, $Delta$ for infection with the Delta variant, or $Alpha \rightarrow Delta$ if an individual is infected with Delta immediately following infection from Alpha). Finally, $k$ denotes the index of the vaccination stratum of individuals (with $k$ corresponding to $V_k$ as defined in Table S2).

$\zeta^{i,k}(t)$ is the rate of movement from vaccination stratum $k$ to vaccination stratum $k+1$ at time $t$, for individuals in group $i$. This was calculated dynamically to match the number of daily doses aimed to be given to each group at time step $t$, given the number of daily doses available for distribution each day, the JCVI priority order, and the uptake in each group. Note that there is no movement out of vaccination stratum 3, so by definition $\zeta^{i,3}(t) = 0$. Additionally we have no movement into vaccination stratum 0, so for ease of notation and equation simplicity we let $k = -1$ be a dummy vaccination strata with empty compartments and let $\zeta^{i,-1}(t) = 0$.

We define all model compartments and parameters in Table S5 below, and illustrate the model structure and flows between compartments in Figure S1 (this figure is copied again below for easy reference). The model assumes discrete time and four time steps are taken per day.

###### 4.1.1 Parallel flows

In addition to compartments involved in the transmission dynamics and clinical progression, there are three parallel flows which we use for fitting to testing data from surveys: (i) one for PCR testing and (ii) two

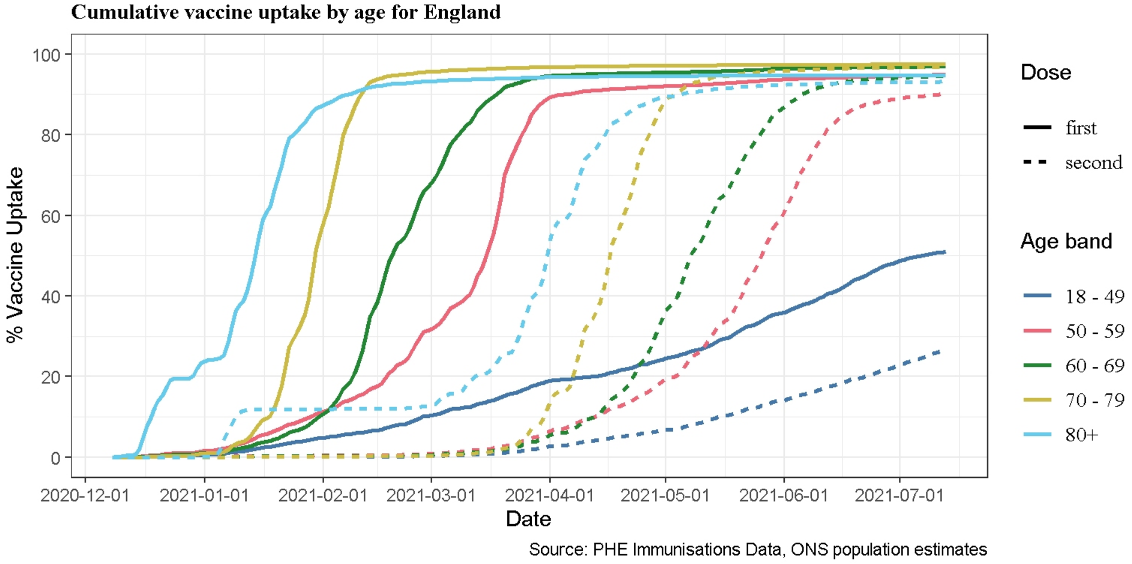

**Figure S5:** Cumulative vaccine uptake by age for England shown for first (solid lines) and second (dashed lines) doses, up to 19 July 2021. Shown as the proportion of the population age group of England (ONS).

for serology testing (Figure S7), with separate flows used for testing with the EuroImmun and Roche N assays.

The PCR flow is used for fitting to data from the REACT-1 study. Upon infection, an individual enters the PCR flow in a pre-positivity compartment ($T_{PCR_{pre}}$) before moving into the PCR positivity compartment ($T_{PCR_{pos}}$) and then ultimately into the PCR negativity compartment ($T_{PCR_{neg}}$).

We have two serology flows to allow us to assume different distributions for the time to seroreversion when fitting to samples tested with two different assays: EuroImmun and Roche N. EuroImmun was used for testing NHSBT samples from the first wave onwards, while Roche N only started being used in November 2020. Roche N tests only for seropositivity resulting from infection, whereas EuroImmun does not distinguish between seropositivity resulting from infection or from vaccination. Since our serology flows only are designed to capture seroconversion resulting from infection, we do not fit to samples using the EuroImmun assay from 15th January 2021 onwards as we can expect the vaccination to impact beyond this. After a seroconversion period ($T_{sero^1_{pre}}$ for EuroImmun, $T_{sero^2_{pre}}$ for Roche N), individuals can seroconvert ($T_{sero^1_{pos}}$ for EuroImmun, $T_{sero^2_{pos}}$ for Roche N) or not ($T_{sero^1_{neg}}$ for EuroImmun, $T_{sero^2_{neg}}$ for Roche N) ; if they do seroconvert, they eventually serorevert to $T_{sero^1_{neg}}$ or $T_{sero^2_{neg}}$ accordingly.

#### 4.2 Equations

##### 4.2.1 Force of infection

We let $\chi^{i,j,k}$ be the susceptibility to variant $j$ of a susceptible individual in group $i$ and vaccine stratum $k$, relative to a non vaccinated individual (so that $\chi^{i,j,0} = 1$ for all $i$ and $j$), given by

$$\chi^{i,j,k} = (1 - e^{i,j,k}_{inf}), \tag{1}$$

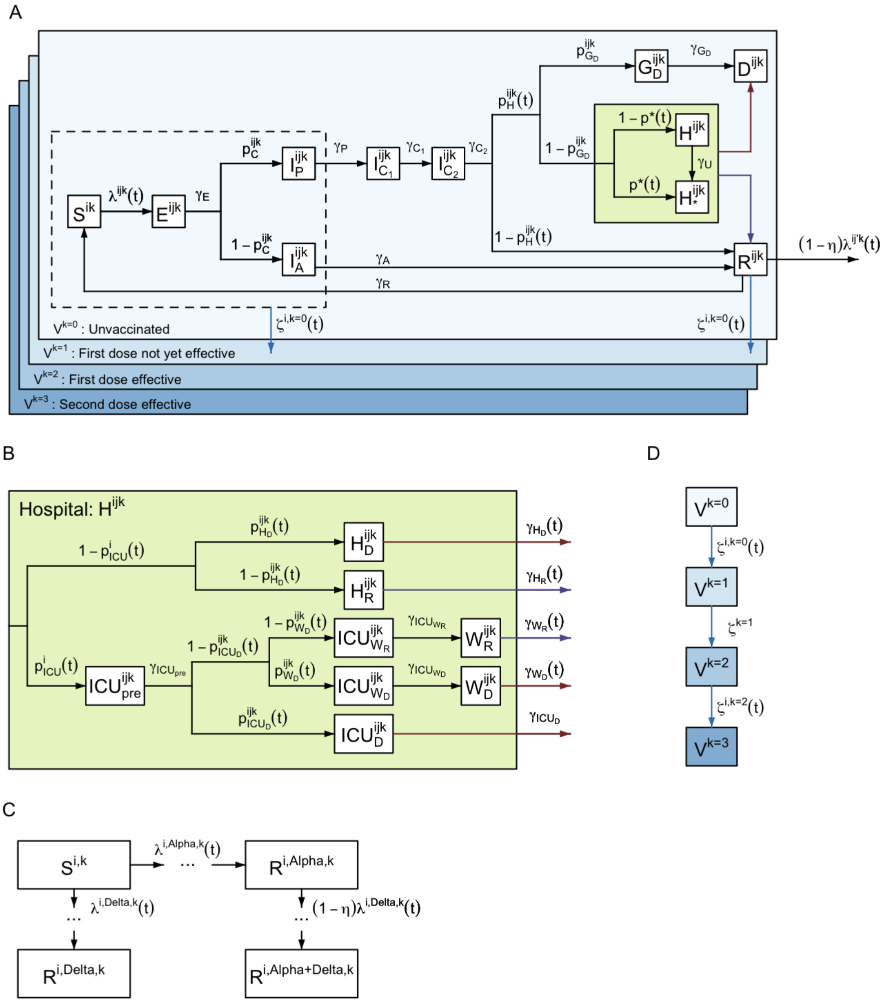

**Figure S6:** See Figure S1 for full details.

where $e_{inf}^{i,j,k}$ is the vaccine effectiveness against infection of variant $j$ in vaccine strata $k$ (Table S4), scaled across vaccine types according to the distribution presented in Figure S4.

We let $\xi^{i,j,k}$ be the infectivity of an individual in group $i$ and vaccine stratum $k$ infected with variant $j$ relative to a non vaccinated individual infected with the Alpha variant (so that $\xi^{i,Alpha,0} = 1$). This infectivity captures both the vaccine effectiveness against infectiousness as presented in Table S3 and also the increased transmissibility of Delta compared to Alpha. As such $\xi^{i,j,k}$ is equal to

$$\xi^{i,j,k} = \begin{cases} (1 - e_{ins}^{i,j,k}), & \text{if } j = Alpha, \\ \\ (1 - e_{ins}^{i,j,k})\sigma, & \text{if } j \in \{Delta, Alpha\rightarrow Delta\}, \end{cases} \tag{2}$$

| Compartment | Definition |
|---|---|
| $S^{i,k}(t)$ | Susceptible |
| $E^{i,j,k}(t)$ | Exposed (latent infection). |
| $I_A^{i,j,k}(t)$ | Asymptomatic infected. |
| $I_P^{i,j,k}(t)$ | Presymptomatic infected (infectious). |
| $I_{C_1}^{i,j,k}(t)$ | Symptomatic infected (infectious). |
| $I_{C_2}^{i,j,k}(t)$ | Symptomatic infected (not infectious). |
| $G_D^{i,j,k}(t)$ | Severely diseased, leading to death (at home). |
| $D^{i,j,k}(t)$ | Deceased (as a result of COVID-19). |
| $H_D^{i,j,k}(t)$ | Hospitalised on general ward leading to death. |
| $H_R^{i,j,k}(t)$ | Hospitalised on general ward leading to recovery. |
| $ICU_{pre}^{i,j,k}(t)$ | Awaiting admission to ICU. |
| $ICU_D^{i,j,k}(t)$ | Hospitalised in ICU, leading to death. |
| $ICU_{W_R}^{i,j,k}(t)$ | Hospitalised in ICU, leading to recovery. |
| $ICU_{W_D}^{i,j,k}(t)$ | Hospitalised in ICU, leading to death following step-down from ICU. |
| $W_R^{i,j,k}(t)$ | Step-down recovery period. |
| $W_D^{i,j,k}(t)$ | Step-down post-ICU period, leading to death. |
| $R^{i,j,k}(t)$ | Recovered. |

**Table S5:** Definitions of model compartments shown in Figure S1. $i$ defines age group ( $i \in \{[0,5),[5,10),\ldots,[75-80),[80+),CHW,CHR\}$ ), $j$ denotes the variant ($j \in \{$ *Alpha*, *Delta*, *Alpha➔Delta* $\}$ as defined in Section 4.1), $k$ denotes vaccination strata ($k \in \{V_0,V_1,V_2,V_3\}$ as defined in Table S2). See Knock et al. (2021) [1] for further details.

where $e_{ins}^{i,j,k}$ is the vaccine effectiveness against infectiousness of variant $j$ in vaccine strata $k$ as defined in Table S3, scaled across vaccine types according to the distribution presented in Figure S4, while $\sigma$ is the region-specific transmission advantage of Delta as described in Section 2.2.

We let $\Theta_{i,j,k}(t)$ be the number of infectious individuals with variant $j$ in group $i$ and vaccination stratum $k$, weighted by infectivity, given by:

$$\Theta_{i,j,k}(t) = \xi^{i,j,k}\left(\theta_{I_A} I_A^{i,j,k}(t) + I_P^{i,j,k}(t) + I_{C_1}^{i,j,k}(t)\right). \quad (3)$$

where $\theta_{I_A}$ is the infectivity of an asymptomatic infected individual, relative to a symptomatic individual infected with the same variant, and in the same vaccination strata.

The force of infection, $\lambda^{i,j,k}(t)$, of variant $j \in \{Alpha, Delta\}$ on a susceptible individual in group $i \in \{[0,5), \ldots,[75,80),[80+), CHW, CHR\}$ and vaccination stratum $k = 0,1,\ldots,3$ is then given by

$$\lambda^{i,Alpha,k}(t) = \chi^{i,Alpha,k} \sum_{i'} m_{i,i'}(t) \sum_{k'} \Theta_{i',Alpha,k'}(t) \quad (4)$$

| Parameter | Definition |
| --- | --- |
| $\lambda^{i,j,k}(t)$ | Force of infection. |
| $\gamma_x$ | Rate of progression from compartment $x$. |
| $\gamma_U$ | Rate at which unconfirmed hospital patients are confirmed as infected. |
| $p_C^{i,j,k}$ | Probability of being symptomatic if infected. |
| $p_H^{i,j,k}$ | Probability of admission to hospital, conditional on symptomatic infection. |
| $p_{G_D}^{i,j,k}$ | Probability of death for severe symptomatic cases outside of hospital. |
| $p^*(t)$ | Probability of COVID-19 diagnosis confirmed prior to admission to hospital. |
| $p_{ICU}^{i}(t)$ | Probability of admission to ICU, conditional on hospitalisation. |
| $p_{H_D}^{i,j,k}(t)$ | Probability of death for hospitalised cases not in ICU. |
| $p_{ICU_D}^{i,j,k}(t)$ | Probability of death for cases in ICU. |
| $p_{W_D}^{i,j,k}(t)$ | Probability of death for cases after discharge from ICU. |
| $\zeta^{i,k}(t)$ | Rate of movement from vaccine strata $k$ to $k+1$. |
| $\eta$ | Protection from infection with *Delta* for those recovered from *Alpha* ($R^{i,Alpha,k}$), relative to those in the susceptible class ($S^{i,k}$). |

**Table S6:** Definitions of model parameters shown in Figure S1. These parameters define the routes of transmission through model compartments defined in Table S5. $i$ defines age group ( $i \in \{[0,5),[5,10),\ldots,[75-80),[80+),CHW,CHR\}$ ), $j$ denotes variant status $j$ denotes the variant ($j \in \{$ *Alpha*, *Delta*, *Alpha→Delta* $\}$ as defined in Section 4.1), $k$ denotes vaccination strata ($k \in \{V_0,V_1,V_2,V_3\}$ as defined in Table S2). Refer to Knock et al. (2021) [1] for further details of parameter fitting.

$$\lambda^{i,Delta,k}(t) = \chi^{i,Delta,k}\sum_{i'} m_{i,i'}(t)\sum_{k'}\left(\Theta_{i',Delta,k'}(t)+\Theta_{i',Alpha\rightarrow Delta,k'}(t)\right) \quad (5)$$

where $m_{i,i'}(t)$ is the (symmetric) time-varying person-to-person transmission rate from group $i'$ to group $i$.

We let $\Lambda^{i,k}(t)$ be the total force of infection on a susceptible individual in group $i$ and vaccination stratum $k$, i.e.

$$\Lambda^{i,k}(t) = \lambda^{i,Alpha,k}(t)+\lambda^{i,Delta,k}(t). \quad (6)$$

Note that there is zero force of infection of *Alpha* on all recovered individuals. The force of infection of *Delta* on an individual recovered from *Alpha* in group $i$ and vaccine stratum $k$ is $(1-\eta)\lambda^{i,Delta,k}(t)$, where $\eta$ is the cross immunity parameter Table S6. There is zero force of infection of *Delta* on all individuals recovered from *Delta* (or in the *Alpha→Delta* class).

Transmission between different age groups $(i,i') \in \{[0,5),\ldots,[75,80),[80+)\}^2$ was parameterised as follows:

$$m_{i,i'}(t) = \beta(t)c_{i,i'}, \quad (7)$$

where $c_{i,i'}$ is the (symmetric) person-to-person contact rate between age group $i$ and $i'$, derived from pre-pandemic data from the POLYMOD survey [2] for the United Kingdom. For each region, the `socialmixr`

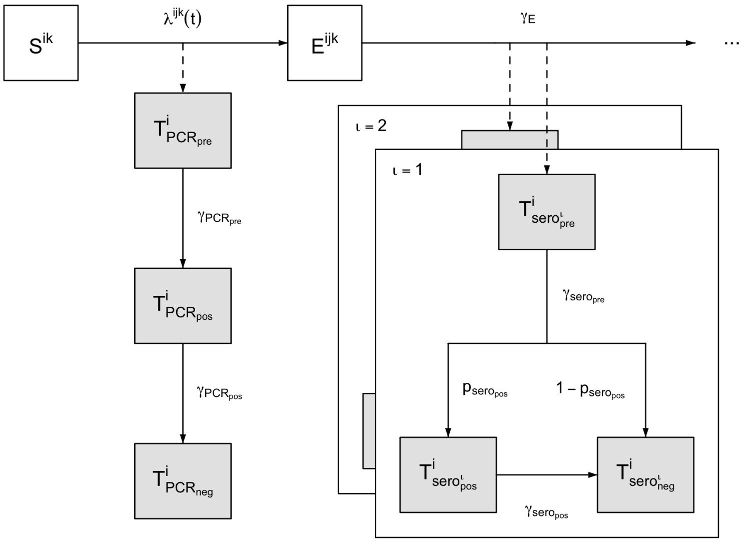

**Figure S7:** PCR positivity and seropositivity model structure flow diagram. Upon infection, an individual enters the pre-positive PCR compartment ($T_{PCR_{pre}}$) before moving into the PCR positivity compartment ($T_{PCR_{pos}}$) and then into the PCR negativity compartment ($T_{PCR_{neg}}$). After a seroreversion period ($T_{sero_{pre}}$), individuals can seroconvert ($T_{sero_{pos}}$) or not ($T_{sero_{neg}}$); if they do seroconvert, they eventually serorevert to $T_{sero_{pos}}$.

package [33] was used to derive the contact matrix between different age groups $(i,i') \in \{[0,5),\ldots,[75,80),[80+)\}^2$, which was then scaled by the regional population demography to yield the required person-to-person daily contact rate matrix, $c_{i,i'}$.

$\beta(t)$ is the time-varying transmission rate which encompasses both changes over time in transmission efficiency (e.g. due to temperature) and temporal changes in the overall level of contacts in the population (due to changes in policy and behaviours).

We assumed $\beta(t)$ to be piecewise linear:

$$\beta(t) = \begin{cases} \beta_i, & \text{if } t \le t_i,\ i = 1 \\ \dfrac{t_i - t}{t_i - t_{i-1}}\beta_{i-1} + \dfrac{t - t_{i-1}}{t_i - t_{i-1}}\beta_i, & \text{if } t_{i-1} < t \le t_i,\ i \in \{2,\ldots,24\} \\ \beta_i & \text{if } t > t_i,\ i = 24 \end{cases} \tag{8}$$

with 24 change points $t_i$ corresponding to major announcements and changes in COVID-19 related policy (Table S8).

We defined parameters representing transmission rates within care homes (between and among workers and residents), which were assumed to be constant over time. Parameter $m_{CHW}$ represents the person-to-person transmission rate among care home workers and between care home workers and residents; $m_{CHR}$ represents the person-to-person transmission rate among care home residents; these are defined as:

$$m_{CHW,CHW}(t) = m_{CHW,CHR}(t) = m_{CHW} \tag{9}$$

$$m_{CHR,CHR}(t) = m_{CHR} \tag{10}$$

Transmission between the general population and care home workers was assumed to be similar to that within the general population, accounting for the average age of care home workers, with, for $i \in \{[0,5), \ldots, [75,80), [80+)\}$,

$$m_{i,CHW}(t) = \beta(t) c_{i,CHW}, \tag{11}$$

where $c_{i,CHW}$ is the mean of $c_{i,[25,30)}, c_{i,[30,35)}, \ldots, c_{i,[60,65)}$ (i.e. of the age groups that the care home workers are drawn from).

Transmission between the general population and care home residents was assumed to be similar to that between the general population and the 80+ age group, adjusted by a reduction factor $\varepsilon$ (which was inferred, see Table S8), such that, for $i \in \{[0,5), \ldots, [75,80), [80+)\}$,

$$m_{i,CHR} = \varepsilon \beta(t) c_{i,80+}. \tag{12}$$

These represent contact between visitors from the general community and care home residents. This might involve a slightly different age profile than the age profile of the contact made by people in the 80+ age group.

###### 4.2.2 Pathway probabilities and rates

The movement between model compartments is primarily dictated by the parameters $p_X^{i,j,k}$, defining the probability of progressing to compartment $x$ (Table S6), as well as rate parameters $\gamma$. These parameters vary between age groups ($i$), variant of infection ($j$), and vaccine strata ($k$). This section outlines how these differences are formally defined and calculated.

The probability that an individual will have a symptomatic infection given that they have been infected is given by

$$p_C^{i,j,k} = p_C^i \left(1 - e_{sympt|inf}^{i,j,k}\right), \tag{13}$$

where $p_C^i$ is given in [1].

The probability that an individual has severe disease requiring hospitalisation given that they have symptoms is given by

$$p_H^{i,j,k}(t) = h_H(t) \psi_H^i \left(1 - e_{SD|sympt}^{i,j,k}\right), \tag{14}$$

where $\psi_H^i$ is given in [1] (with a slight amendment that now $\psi_H^{CHR} = 1$) and $h_H(t)$ has a piecewise linear form with changepoints defined as follows:

$$h_H = (t) \begin{cases} p_{H,1}^{max} & \text{on (and before) 2020-10-01,} \\ p_{H,2}^{max} & \text{on 2020-12-15,} \\ p_{H,3}^{max} & \text{on (and after) 2021-02-02.} \end{cases} \tag{15}$$

The probability that an individual dies in the community (or a care home if $i = CHR$) given they have severe disease is

$$p_{G_D}^i(t) = \begin{cases} p_{G_D} & \text{if } i \neq CHR, \\ p_{G_D}^{CHR} & \text{if } i = CHR. \end{cases} \tag{16}$$

The probability that an individual will be admitted to ICU given that they have been hospitalised is

$$p^i_{ICU}(t) = h_{ICU}(t)\psi^i_{ICU}, \tag{17}$$

where $\psi^i_{ICU}$ is given in [1] and $h_{ICU}(t)$ has a piecewise linear form with changepoints defined as follows:

$$h_{ICU} = (t)\begin{cases} p^{max}_{ICU,1} & \text{on (and before) 2020-04-01,} \\ p^{max}_{ICU,2} & \text{on 2020-06-01.} \end{cases} \tag{18}$$

The probability that an individual will die in general beds given that they are not admitted to ICU is

$$p^{i,j,k}_{H_D}(t) = \min\left\{p^{max}_{H_D} h_D(t)\psi^i_{H_D} e^{i,j,k}_{death|SD}\omega_j, 1\right\}, \tag{19}$$

where $\psi^i_{H_D}$ is given in [1], $h_D(t)$ has a piecewise linear form with changepoints defined as follows:

$$h_D = (t)\begin{cases} 1 & \text{on (and before) 2020-04-01,} \\ \mu^{max}_{D,1} & \text{on 2020-06-01,} \\ \mu^{max}_{D,1} & \text{on 2020-10-01,} \\ \mu^{max}_{D,2} & \text{on 2020-12-15,} \\ \mu^{max}_{D,2} & \text{on 2021-01-15,} \\ \mu^{max}_{D,1} & \text{on (and after) 2021-02-01,} \end{cases} \tag{20}$$

and $\omega_j$ is a multiplier accounting for the increased severity of $Delta$ (see Section 2.4):

$$\omega_j = \begin{cases} 1 & \text{if } j = Alpha, \\ 1.85 & \text{if } j = Delta. \end{cases} \tag{21}$$

The probability that an individual dies in ICU given that they are not admitted to ICU is

$$p^{i,j,k}_{ICU_D}(t) = p^{max}_{ICU_D} h_D(t)\psi^i_{ICU_D} e^{i,j,k}_{death|SD} \tag{22}$$

where $\psi^i_{ICU_D}$ is given in [1].

The probability that an individual who has been in ICU dies in stepdown beds given that they have not died in ICU is

$$p^{i,j,k}_{W_D}(t) = p^{max}_{W_D} h_D(t)\psi^i_{W_D} e^{i,j,k}_{death|SD}, \tag{23}$$

where $\psi^i_{W_D}$ is given in [1].

Finally, the probability of individuals having had a COVID-19 diagnosis confirmed prior to admission to hospital, $p^*(t)$ has a piecewise linear form with changepoints defined as follows:

$$p^*(t) = \begin{cases} 0.1 & \text{on (and before) 2020-03-15,} \\ 0.42 & \text{on 2020-07-01,} \\ 0.2 & \text{on 2020-09-09,} \\ 0.45 & \text{on (and after) 2021-06-27.} \end{cases} \tag{24}$$

These were informed by data on COVID-19 admissions and inpatient diagnoses for England from NHS England [34].

In addition, the duration rates for some hospital compartments are time-varying to account for changes in length of stay over time. We let

$$\gamma_{H_R}(t) = h_\gamma(t)\gamma_{H_R} \tag{25}$$
$$\gamma_{H_D}(t) = h_\gamma(t)\gamma_{H_D} \tag{26}$$
$$\gamma_{W_R}(t) = h_\gamma(t)\gamma_{W_R} \tag{27}$$
$$\gamma_{W_D}(t) = h_\gamma(t)\gamma_{W_D} \tag{28}$$
$$\tag{29}$$

where $h_\gamma(t)$ has a piecewise linear form with changepoints given by

$$h_\gamma(t) = \begin{cases} 1 & \text{on (and before) 2020-12-01,} \\ 1/\mu_{\gamma_H,1} & \text{on 2021-01-01,} \\ 1/\mu_{\gamma_H,2} & \text{on 2021-03-01,} \\ 1/\mu_{\gamma_H,3} & \text{on (and after) 2021-06-15.} \end{cases} \tag{30}$$

###### 4.2.3 Compartmental model equations

To clearly illustrate the model dynamics, we describe a deterministic version of the model in differential equations (32)-(72), followed by the stochastic implementation used in the analysis. Full definitions of compartments and model parameters are set out in Tables S5 and S6. Unless otherwise specified, $\sum_j$ refers to the sum across all combinations of variants (i.e. $j \in \{Alpha, Delta, Alpha\text{→}Delta\}$), and $\sum_k$ refers to the sum across all vaccination strata ($k \in \{0,1,2,3\}$).

In the following model equations we use $\mathbb{1}_A$ as an indicator function, such that

$$\mathbb{1}_A(j) := \begin{cases} 1 & \text{if } j \in A, \\ 0 & \text{if } j \notin A. \end{cases} \tag{31}$$

Further note that we split some compartments in two distinct compartments. For example, the exposed class, $E^{i,j,k}$, is modelled via two separate compartments, $E^{i,j,k,1}$ and $E^{i,j,k,2}$ (equations (33) and (34)). This is to be able to capture a non exponentially distributed duration of stay in certain compartments; the split allows to model the duration of stay as an Erlang distribution instead (sum of independent exponential distributions) [35].

$$\frac{dS^{i,k}(t)}{dt} = \zeta^{i,k-1}(t)S^{i,k-1}(t) - \left(\zeta^{i,k}(t) + \Lambda^{i,k}(t)\right)S^{i,k}(t) + \gamma_R \sum_j R^{i,j,k}(t) \tag{32}$$

$$\begin{aligned}\frac{dE^{i,j,k,1}(t)}{dt} &= \mathbb{1}_{\{Alpha,Delta\}}(j)\lambda^{i,j,k}(t)S^{i,k}(t) + \mathbb{1}_{\{Alpha\text{→}Delta\}}(j)\,(1-\eta)\,\lambda^{i,Delta,k}(t)R^{i,Alpha,k}(t) \\ &\quad + \zeta^{i,k-1}(t)E^{i,j,k-1,1}(t) - \left(\gamma_E + \zeta^{i,k}(t)\right)E^{i,j,k,1}(t)\end{aligned} \tag{33}$$

$$\frac{dE^{i,j,k,2}(t)}{dt} = \gamma_E E^{i,j,k,1}(t) + \zeta^{i,k-1}(t)E^{i,j,k-1,2}(t) - \left(\gamma_E + \zeta^{i,k}(t)\right)E^{i,j,k,2}(t) \tag{34}$$

$$\frac{dI_A^{i,j,k}(t)}{dt} = \left(1 - p_C^{i,j,k}\right)\gamma_E E^{i,j,k,2}(t) + \zeta^{i,k-1}(t)I_A^{i,j,k-1}(t) - \left(\gamma_A + \zeta^{i,k}(t)\right)I_A^{i,j,k}(t) \tag{35}$$

$$\frac{dI_P^{i,j,k}(t)}{dt} = p_C^{i,j,k}\gamma_E E^{i,j,k,2}(t) + \zeta^{i,k-1}(t)I_P^{i,j,k-1}(t) - \left(\gamma_P + \zeta^{i,k}(t)\right)I_P^{i,j,k}(t) \tag{36}$$

$$\frac{dI_{C_1}^{i,j,k}(t)}{dt} = \gamma_P I_P^{i,j,k}(t) - \gamma_{C_1} I_{C_1}^{i,j,k}(t) \tag{37}$$

$$\frac{dI_{C_2}^{i,j,k}(t)}{dt} = \gamma_{C_1} I_{C_1}^{i,j,k}(t) - \gamma_{C_2} I_{C_2}^{i,j,k}(t) \tag{38}$$

$$\frac{dG_D^{i,j,k,1}(t)}{dt} = p_H^{i,j,k}(t) p_{G_D}^{i,j,k} \gamma_{C_2} I_{C_2}^{i,j,k}(t) - \gamma_{G_D} G_D^{i,j,k,1}(t) \tag{39}$$

$$\frac{dG_D^{i,j,k,2}(t)}{dt} = \gamma_{G_D} G_D^{i,j,k,1}(t) - \gamma_{G_D} G_D^{i,j,k,2}(t) \tag{40}$$

$$\begin{aligned}\frac{dICU_{pre}^{i,j,k}(t)}{dt} &= p_H^{i,j,k}(t)\left(1 - p_{G_D}^{i,j,k}\right)(1 - p^*(t))\, p_{ICU}^{i}(t) \gamma_{C_2} I_{C_2}^{i,j,k}(t) \\ &\quad - \left(\gamma_{ICU_{pre}} + \gamma_U\right) ICU_{pre}^{i,j,k}(t)\end{aligned} \tag{41}$$

$$\begin{aligned}\frac{dICU_{pre^*}^{i,j,k}(t)}{dt} &= p_H^{i,j,k}(t)\left(1 - p_{G_D}^{i,j,k}\right) p^*(t) p_{ICU}^{i}(t) \gamma_{C_2} I_{C_2}^{i,j,k}(t) - \gamma_{ICU_{pre}} ICU_{pre^*}^{i,j,k}(t) \\ &\quad + \gamma_U ICU_{pre}^{i,j,k}(t)\end{aligned} \tag{42}$$

$$\begin{aligned}\frac{dICU_{W_R}^{i,j,k}(t)}{dt} &= \left(1 - p_{ICU_D}^{i,j,k}(t)\right)\left(1 - p_{W_D}^{i,j,k}(t)\right) \gamma_{ICU_{pre}} ICU_{pre}^{i,j,k}(t) \\ &\quad - \left(\gamma_{ICU_{W_R}} + \gamma_U\right) ICU_{W_R}^{i,j,k}(t)\end{aligned} \tag{43}$$

$$\begin{aligned}\frac{dICU_{W_{R^*}}^{i,j,k}(t)}{dt} &= \left(1 - p_{ICU_D}^{i,j,k}(t)\right)\left(1 - p_{W_D}^{i,j,k}(t)\right) \gamma_{ICU_{pre}} ICU_{pre^*}^{i,j,k}(t) - \gamma_{ICU_{W_R}} ICU_{W_{R^*}}^{i,j,k}(t) \\ &\quad + \gamma_U ICU_{W_R}^{i,j,k}(t)\end{aligned} \tag{44}$$

$$\frac{dICU_{W_D}^{i,j,k}(t)}{dt} = \left(1 - p_{ICU_D}^{i,j,k}(t)\right) p_{W_D}^{i,j,k}(t) \gamma_{ICU_{pre}} ICU_{pre}^{i,j,k}(t) - \left(\gamma_{ICU_{W_D}} + \gamma_U\right) ICU_{W_D}^{i,j,k}(t) \tag{45}$$

$$\begin{aligned}\frac{dICU_{W_{D^*}}^{i,j,k}(t)}{dt} &= \left(1 - p_{ICU_D}^{i,j,k}(t)\right) p_{W_D}^{i,j,k}(t) \gamma_{ICU_{pre}} ICU_{pre^*}^{i,j,k}(t) - \gamma_{ICU_{W_D}} ICU_{W_{D^*}}^{i,j,k}(t) \\ &\quad + \gamma_U ICU_{W_D}^{i,j,k}(t)\end{aligned} \tag{46}$$

$$\frac{dICU_D^{i,j,k,1}(t)}{dt} = p_{ICU_D}^{i,j,k}(t) \gamma_{ICU_{pre}} ICU_{pre}^{i,j,k}(t) - \left(\gamma_{ICU_D} + \gamma_U\right) ICU_D^{i,j,k,1}(t) \tag{47}$$

$$\frac{dICU_D^{i,j,k,2}(t)}{dt} = \gamma_{ICU_D} ICU_D^{i,j,k,1}(t) - \left(\gamma_{ICU_D} + \gamma_U\right) ICU_D^{i,j,k,2}(t) \tag{48}$$

$$\frac{dICU_{D^*}^{i,j,k,1}(t)}{dt} = p_{ICU_D}^{i,j,k}(t) \gamma_{ICU_{pre}} ICU_{pre^*}^{i,j,k}(t) - \gamma_{ICU_D} ICU_{D^*}^{i,j,k,1}(t) + \gamma_U ICU_D^{i,j,k,1}(t) \tag{49}$$

$$\frac{dICU_{D^*}^{i,j,k,2}(t)}{dt} = \gamma_{ICU_D} ICU_{D^*}^{i,j,k,1}(t) - \gamma_{ICU_D} ICU_{D^*}^{i,j,k,2}(t) + \gamma_U ICU_D^{i,j,k,2}(t) \tag{50}$$

$$\frac{dW_R^{i,j,k,1}(t)}{dt} = \gamma_{ICU_{W_R}} ICU_{W_R}^{i,j,k}(t) - \left(\gamma_{W_R}(t) + \gamma_U\right) W_R^{i,j,k,1}(t) \tag{51}$$

$$\frac{dW_R^{i,j,k,2}(t)}{dt} = \gamma_{W_R}(t) W_R^{i,j,k,1}(t) - \left(\gamma_{W_R}(t) + \gamma_U\right) W_R^{i,j,k,2}(t) \tag{52}$$

$$\frac{dW_{R^*}^{i,j,k,1}(t)}{dt} = \gamma_{ICU_{W_R}} ICU_{W_{R^*}}^{i,j,k}(t) - \gamma_{W_R}(t) W_{R^*}^{i,j,k,1}(t) + \gamma_U W_R^{i,j,k,1}(t) \tag{53}$$

$$\frac{dW_{R^*}^{i,j,k,2}(t)}{dt} = \gamma_{W_R}(t)W_{R^*}^{i,j,k,1}(t) - \gamma_{W_R}(t)W_{R^*}^{i,j,k,2}(t) + \gamma_U W_R^{i,j,k,2}(t) \tag{54}$$

$$\frac{dW_D^{i,j,k}(t)}{dt} = \gamma_{ICU_{W_D}} ICU_{W_D}^{i,j,k}(t) - (\gamma_{W_D}(t) + \gamma_U) W_D^{i,j,k}(t) \tag{55}$$

$$\frac{dW_{D^*}^{i,j,k}(t)}{dt} = \gamma_{ICU_{W_D}} ICU_{W_{D^*}}^{i,j,k}(t) - \gamma_{W_D}(t)W_{D^*}^{i,j,k}(t) + \gamma_U W_D^{i,j,k}(t) \tag{56}$$

$$\begin{aligned}\frac{dH_R^{i,j,k}(t)}{dt} &= p_H^{i,j,k}(t)\left(1 - p_{G_D}^{i,j,k}\right)(1 - p^*(t))\left(1 - p_{ICU}^i(t)\right)\left(1 - p_{H_D}^{i,j,k}(t)\right)\gamma_{C_2} I_{C_2}^{i,j,k}(t) \\ &\quad - (\gamma_{H_R}(t) + \gamma_U) H_R^{i,j,k}(t)\end{aligned} \tag{57}$$

$$\begin{aligned}\frac{dH_{R^*}^{i,j,k}(t)}{dt} &= p_H^{i,j,k}(t)\left(1 - p_{G_D}^{i,j,k}\right)p^*(t)\left(1 - p_{ICU}^i(t)\right)\left(1 - p_{H_D}^{i,j,k}(t)\right)\gamma_{C_2} I_{C_2}^{i,j,k}(t) \\ &\quad + \gamma_U H_R^{i,j,k}(t) - \gamma_{H_R}(t) H_{R^*}^{i,j,k}(t)\end{aligned} \tag{58}$$

$$\begin{aligned}\frac{dH_D^{i,j,k,1}(t)}{dt} &= p_H^{i,j,k}(t)\left(1 - p_{G_D}^{i,j,k}\right)(1 - p^*(t))\left(1 - p_{ICU}^i(t)\right)p_{H_D}^{i,j,k}(t)\gamma_{C_2} I_{C_2}^{i,j,k}(t) \\ &\quad - (\gamma_{H_D}(t) + \gamma_U) H_D^{i,j,k,1}(t)\end{aligned} \tag{59}$$

$$\frac{dH_D^{i,j,k,2}(t)}{dt} = \gamma_{H_D}(t)H_D^{i,j,k,1}(t) - (\gamma_{H_D}(t) + \gamma_U) H_D^{i,j,k,2}(t) \tag{60}$$

$$\begin{aligned}\frac{dH_{D^*}^{i,j,k,1}(t)}{dt} &= p_H^{i,j,k}(t)\left(1 - p_{G_D}^{i,j,k}\right)p^*(t)\left(1 - p_{ICU}^i(t)\right)\; p_{H_D}^{i,j,k}(t)\gamma_{C_2} I_{C_2}^{i,j,k}(t) + \gamma_U H_D^{i,j,k,1}(t) \\ &\quad - \gamma_{H_D}(t) H_{D^*}^{i,j,k,1}(t)\end{aligned} \tag{61}$$

$$\frac{dH_{D^*}^{i,j,k,2}(t)}{dt} = \gamma_{H_D}(t)H_{D^*}^{i,j,k,1}(t) - \gamma_{H_D}(t)H_{D^*}^{i,j,k,2}(t) + \gamma_U H_D^{i,j,k,2}(t) \tag{62}$$

$$\begin{aligned}\frac{dR^{i,j,k}(t)}{dt} &= \gamma_A I_A^{i,j,k}(t) + \left(1 - p_H^{i,j,k}(t)\right)\gamma_{C_2} I_{C_2}^{i,j,k}(t) + \gamma_{H_R}(t)\left(H_R^{i,j,k}(t) + H_{R^*}^{i,j,k}(t)\right) \\ &\quad + \gamma_{W_R}(t)\left(W_R^{i,j,k,2}(t) + W_{R^*}^{i,j,k,2}(t)\right) + \zeta^{i,k-1}(t)R^{i,j,k-1}(t) - \left(\gamma_R + \zeta^{i,k}(t)\right)R^{i,j,k}(t) \\ &\quad - \mathbb{1}_{\{Alpha\}}(j)\,(1-\eta)\,\lambda^{i,Delta,k}(t)R^{i,j,k}(t)\end{aligned} \tag{63}$$

$$\frac{dT_{sero_{pre}^1}^i(t)}{dt} = -\gamma_{sero_{pre}} T_{sero_{pre}^1}^i(t) + \sum_j \sum_k \gamma_E E^{i,j,k,2}(t) \tag{64}$$

$$\frac{dT_{sero_{pos}^1}^i(t)}{dt} = p_{sero_{pos}} \gamma_{sero_{pre}} T_{sero_{pre}^1}^i(t) - \gamma_{sero_{pos}^1} T_{sero_{pos}^1}^i(t) \tag{65}$$

$$\frac{dT_{sero_{neg}^1}^i(t)}{dt} = \left(1 - p_{sero_{pos}}\right)\gamma_{sero_{pre}} T_{sero_{pre}^1}^i(t) + \gamma_{sero_{pos}^1} T_{sero_{pos}^1}^i(t) \tag{66}$$

$$\frac{dT_{sero_{pre}^2}^i(t)}{dt} = -\gamma_{sero_{pre}} T_{sero_{pre}^2}^i(t) + \sum_j \sum_k \gamma_E E^{i,j,k,2}(t) \tag{67}$$

$$\frac{dT_{sero_{pos}^2}^i(t)}{dt} = p_{sero_{pos}} \gamma_{sero_{pre}} T_{sero_{pre}^2}^i(t) - \gamma_{sero_{pos}^2} T_{sero_{pos}^2}^i(t) \tag{68}$$

$$\frac{dT_{sero_{neg}^2}^i(t)}{dt} = \left(1 - p_{sero_{pos}}\right)\gamma_{sero_{pre}} T_{sero_{pre}^2}^i(t) + \gamma_{sero_{pos}^2} T_{sero_{pos}^2}^i(t) \tag{69}$$

$$\frac{dT^i_{PCR_{pre}}(t)}{dt} = -\gamma_{PCR_{pre}} T^i_{PCR_{pre}}(t) + \sum_k \left(\lambda^{i,Alpha,k}(t) + \lambda^{i,Delta,k}(t)\right) S^{i,k}(t) \tag{70}$$

$$\frac{dT^i_{PCR_{pos}}(t)}{dt} = \gamma_{PCR_{pre}} T^i_{PCR_{pre}}(t) - \gamma_{PCR_{pos}} T^i_{PCR_{pos}}(t) \tag{71}$$

$$\frac{dT^i_{PCR_{neg}}(t)}{dt} = \gamma_{PCR_{pos}} T^i_{PCR_{pos}}(t). \tag{72}$$

We used the tau-leap method [36] to create a stochastic, time-discretised version of the model described in equations (75) - (206), taking four update steps per day ($dt = 0.25$ days).

For each time step, the model iterated through the procedure described below. In the following, we introduce a small abuse of notation: for transitions involving multiple onward compartments (e.g. transition from compartment $E$ to compartments $I_A$ or $I_P$ or to the next vaccination strata within $E$), for conciseness, we write

$$\left(d^{i,j,k}_{E,I_A},\, d^{i,j,k}_{E,I_P},\, d^{i,j,k}_{E,v}\right) \sim \text{Multinom}\left(E^{i,j,k,2}(t),\, q^{i,j,k}_{E,I_A},\, q^{i,j,k}_{E,I_P},\, q^{i,j,k}_{E,v}\right) \tag{73}$$

instead of

$$\left(d^{i,j,k}_{E,I_A},\, d^{i,j,k}_{E,I_P},\, d^{i,j,k}_{E,v},\, d^{i,j,k}_{nomove}\right) \sim \text{Multinom}\left(E^{i,j,k,2}(t),\, q^{i,j,k}_{E,I_A},\, q^{i,j,k}_{E,I_P},\, q^{i,j,k}_{E,v},\, 1 - \sum_{x \in I_A, I_P, v} q^{i,j,k}_{E,x}\right) \tag{74}$$

where $d^{i,j,k}_{nomove}$ is a dummy variable counting the number of individuals remaining in compartment $E^{i,j,k,2}$. We also omit the time dependency i.e. we use $d^{i,j,k}_{E,I_A}$ or $q^{i,j,k}_{E,I_A}$ instead of $d^{i,j,k}_{E,I_A}(t)$ or $q^{i,j,k}_{E,I_A}(t)$.

Using this convention, transition variables are drawn from the following distributions, with probabilities defined below:

$$q^{i,Alpha,k}_{S,E} = \left(1 - e^{-\Lambda^{i,k}(t)dt}\right) \frac{\lambda^{i,Alpha,k}(t)}{\Lambda^{i,k}(t)} \tag{75}$$

$$q^{i,Delta,k}_{S,E} = \left(1 - e^{-\Lambda^{i,k}(t)dt}\right) \frac{\lambda^{i,Delta,k}(t)}{\Lambda^{i,k}(t)} \tag{76}$$

$$q^{i,k}_{S,v} = e^{-\Lambda^{i,k}(t)dt} \left(1 - e^{-\zeta^{i,k}(t)dt}\right) \tag{77}$$

$$\left(d^{i,Alpha,k}_{S,E},\, d^{i,Delta,k}_{S,E},\, d^{i,k}_{S,v}\right) \sim \text{Multinom}\left(S^{i,k}(t),\, q^{i,Alpha,k}_{S,E},\, q^{i,Delta,k}_{S,E},\, q^{i,k}_{S,v}\right) \tag{78}$$

$$\left(q^{i,j,k}_{E,E},\, q^{i,j,k,1}_{E,v}\right) = \left(1 - e^{-\gamma_E dt},\, e^{-\gamma_E dt}\left(1 - e^{-\zeta^{i,k}(t)dt}\right)\right) \tag{79}$$

$$\left(d^{i,j,k}_{E,E},\, d^{i,j,k,1}_{E,v}\right) \sim \text{Multinom}\left(E^{i,j,k,1}(t),\, q^{i,j,k}_{E,E},\, q^{i,j,k,1}_{E,v}\right) \tag{80}$$

$$q^{i,j,k}_{E,I_A} = \left(1 - p^{i,j,k}_C\right)\left(1 - e^{-\gamma_E dt}\right) \tag{81}$$

$$q^{i,j,k}_{E,I_P} = p^{i,j,k}_C \left(1 - e^{-\gamma_E dt}\right) \tag{82}$$

$$q^{i,j,k,2}_{E,v} = e^{-\gamma_E dt}\left(1 - e^{-\zeta^{i,k}(t)dt}\right) \tag{83}$$

$$\left(d^i_{E,I_A},\, d^i_{E,I_P},\, d^{i,j,k,2}_{E,v}\right) \sim \text{Multinom}\left(E^{i,j,k,2}(t),\, q^{i,j,k}_{E,I_A},\, q^{i,j,k}_{E,I_P},\, q^{i,j,k,2}_{E,v}\right) \tag{84}$$

$$\left(q_{I_A,R}^{i,j,k},\ q_{I_A,v}^{i,j,k}\right) = \left(1-e^{-\gamma_A dt},\ e^{-\gamma_A dt}\left(1-e^{-\zeta^{i,k}(t)dt}\right)\right) \tag{85}$$

$$\left(d_{I_A,R}^{i,j,k},\ d_{I_A,v}^{i,j,k}\right) \sim \text{Multinom}\left(I_A^i(t),\ q_{I_A,R}^{i,j,k},\ q_{I_A,v}^{i,j,k}\right) \tag{86}$$

$$\left(q_{I_P,I_{C_1}}^{i,j,k},\ q_{I_P,v}^{i,j,k}\right) = \left(1-e^{-\gamma_P dt},\ e^{-\gamma_P dt}\left(1-e^{-\zeta^{i,k}(t)dt}\right)\right) \tag{87}$$

$$\left(d_{I_P,I_{C_1}}^{i,j,k},\ d_{I_P,v}^{i,j,k}\right) \sim \text{Multinom}\left(I_P^i(t),\ q_{I_P,I_{C_1}}^{i,j,k},\ q_{I_P,v}^{i,j,k}\right) \tag{88}$$

$$d_{I_{C_1},I_{C_2}}^{i,j,k} \sim \text{Binom}\left(I_{C_1}^{i,j,k}(t),\ 1-e^{-\gamma_{C_1} dt}\right) \tag{89}$$

$$q_{I_{C_2},G_D}^{i,j,k} = p_H^{i,j,k}(t)p_{G_D}^{i,j,k}\left(1-e^{-\gamma_{C_2} dt}\right) \tag{90}$$

$$q_{I_{C_2},R}^{i,j,k} = \left(1-p_H^{i,j,k}(t)\right)\left(1-e^{-\gamma_{C_2} dt}\right) \tag{91}$$

$$q_{I_{C_2},ICU_{pre}}^{i,j,k} = p_H^{i,j,k}(t)\left(1-p_{G_D}^{i,j,k}\right)\left(1-p^*(t)\right)p_{ICU}^i(t)\left(1-e^{-\gamma_{C_2} dt}\right) \tag{92}$$

$$q_{I_{C_2},ICU_{pre^*}}^{i,j,k} = p_H^{i,j,k}(t)\left(1-p_{G_D}^{i,j,k}\right)p^*(t)p_{ICU}^i(t)\left(1-e^{-\gamma_{C_2} dt}\right) \tag{93}$$

$$q_{I_{C_2},H_R}^{i,j,k} = p_H^{i,j,k}(t)\left(1-p_{G_D}^{i,j,k}\right)\left(1-p^*(t)\right)\left(1-p_{ICU}^i(t)\right)\left(1-p_{H_D}^{i,j,k}(t)\right)\left(1-e^{-\gamma_{C_2} dt}\right) \tag{94}$$

$$q_{I_{C_2},H_{R^*}}^{i,j,k} = p_H^{i,j,k}(t)\left(1-p_{G_D}^{i,j,k}\right)\ p^*(t)\left(1-p_{ICU}^i(t)\right)\left(1-p_{H_D}^{i,j,k}(t)\right)\left(1-e^{-\gamma_{C_2} dt}\right) \tag{95}$$

$$q_{I_{C_2},H_D}^{i,j,k} = p_H^{i,j,k}(t)\left(1-p_{G_D}^{i,j,k}\right)\left(1-p^*(t)\right)\left(1-p_{ICU}^i(t)\right)p_{H_D}^{i,j,k}(t)\left(1-e^{-\gamma_{C_2} dt}\right) \tag{96}$$

$$q_{I_{C_2},H_{D^*}}^{i,j,k} = p_H^{i,j,k}(t)\left(1-p_{G_D}^{i,j,k}\right)p^*(t)\left(1-p_{ICU}^i(t)\right)p_{H_D}^{i,j,k}(t)\left(1-e^{-\gamma_{C_2} dt}\right) \tag{97}$$

$$\left(d_{I_{C_2},G_D}^{i,j,k},\ldots,d_{I_{C_2},H_{D^*}}^{i,j,k}\right) \sim \text{Multinom}\left(I_{C_2}^{i,j,k}(t),\ q_{I_{C_2},G_D}^{i,j,k},\ldots,q_{I_{C_2},H_{D^*}}^{i,j,k}\right) \tag{98}$$

$$d_{G_D,G_D}^{i,j,k} \sim \text{Binom}\left(G_D^{i,j,k,1}(t),\ 1-e^{-\gamma_{G_D} dt}\right) \tag{99}$$

$$d_{G_D,D}^{i,j,k} \sim \text{Binom}\left(G_D^{i,j,k,2}(t),\ 1-e^{-\gamma_{G_D} dt}\right) \tag{100}$$

$$q_{ICU_{pre},ICU_{W_R}}^{i,j,k} = \left(1-p_{ICU_D}^{i,j,k}(t)\right)\left(1-p_{W_D}^{i,j,k}(t)\right)\left(1-e^{-\gamma_{ICU_{pre}} dt}\right)e^{-\gamma_U dt} \tag{101}$$

$$q_{ICU_{pre},ICU_{W_{R^*}}}^{i,j,k} = \left(1-p_{ICU_D}^{i,j,k}(t)\right)\left(1-p_{W_D}^{i,j,k}(t)\right)\left(1-e^{-\gamma_{ICU_{pre}} dt}\right)\left(1-e^{-\gamma_U dt}\right) \tag{102}$$

$$q_{ICU_{pre},ICU_{W_D}}^{i,j,k} = \left(1-p_{ICU_D}^{i,j,k}(t)\right)p_{W_D}^{i,j,k}(t)\left(1-e^{-\gamma_{ICU_{pre}} dt}\right)e^{-\gamma_U dt} \tag{103}$$

$$q_{ICU_{pre},ICU_{W_{D^*}}}^{i,j,k} = \left(1-p_{ICU_D}^{i,j,k}(t)\right)p_{W_D}^{i,j,k}(t)\left(1-e^{-\gamma_{ICU_{pre}} dt}\right)\left(1-e^{-\gamma_U dt}\right) \tag{104}$$

$$q_{ICU_{pre},ICU_D}^{i,j,k} = p_{ICU_D}^{i,j,k}(t)\left(1-e^{-\gamma_{ICU_{pre}} dt}\right)e^{-\gamma_U dt} \tag{105}$$

$$q_{ICU_{pre},ICU_{D^*}}^{i,j,k} = p_{ICU_D}^{i,j,k}(t)\left(1-e^{-\gamma_{ICU_{pre}} dt}\right)\left(1-e^{-\gamma_U dt}\right) \tag{106}$$

$$q_{ICU_{pre},ICU_{pre^*}}^{i,j,k} = e^{-\gamma_{ICU_{pre}} dt}\left(1-e^{-\gamma_U dt}\right) \tag{107}$$

$$\begin{aligned}\left(d_{ICU_{pre},ICU_{W_R}}^{i,j,k},\ \ldots,\ d_{ICU_{pre},ICU_{pre^*}}^{i,j,k}\right)& \\ \sim \text{Multinom}&\left(ICU_{pre}^{i,j,k}(t),\ q_{ICU_{pre},ICU_{W_R}}^{i,j,k},\ldots,q_{ICU_{pre},ICU_{pre^*}}^{i,j,k}\right)\end{aligned} \tag{108}$$

$$q_{ICU_{pre^*},ICU_{W_{R^*}}}^{i,j,k} = \left(1-p_{ICU_D}^{i,j,k}(t)\right)\left(1-p_{W_D}^{i,j,k}(t)\right)\left(1-e^{-\gamma_{ICU_{pre}} dt}\right) \tag{109}$$

$$q^{i,j,k}_{ICU_{pre^*},ICU_{W_{D^*}}} = \left(1 - p^{i,j,k}_{ICU_D}(t)\right) p^{i,j,k}_{W_D}(t) \left(1 - e^{-\gamma_{ICU_{pre}} dt}\right) \tag{110}$$

$$q^{i,j,k}_{ICU_{pre^*},ICU_{D^*}} = p^{i,j,k}_{ICU_D}(t) \left(1 - e^{-\gamma_{ICU_{pre}} dt}\right) \tag{111}$$

$$\begin{aligned} &\left(d^{i,j,k}_{ICU_{pre^*},ICU_{W_{R^*}}}, \ \ldots, \ d^{i,j,k}_{ICU_{pre^*},ICU_{D^*}}\right) \\ &\qquad \sim \text{Multinom}\left(ICU^{i,j,k}_{pre^*}(t), \ q^{i,j,k}_{ICU_{pre^*},ICU_{W_{R^*}}}, \ldots, q^{i,j,k}_{ICU_{pre^*},ICU_{D^*}}\right) \end{aligned} \tag{112}$$

$$q^{i,j,k}_{H_D,H_D} = \left(1 - e^{-\gamma_{H_D}(t) dt}\right) e^{-\gamma_U dt} \tag{113}$$

$$q^{i,j,k,1,1}_{H_D,H_{D^*}} = e^{-\gamma_{H_D}(t) dt} \left(1 - e^{-\gamma_U dt}\right) \tag{114}$$

$$q^{i,j,k,1,2}_{H_D,H_{D^*}} = \left(1 - e^{-\gamma_{H_D}(t) dt}\right) \left(1 - e^{-\gamma_U dt}\right) \tag{115}$$

$$\begin{aligned} &\left(d^{i,j,k}_{H_D,H_D}, \ d^{i,j,k,1,1}_{H_D,H_{D^*}}, \ d^{i,j,k,1,2}_{H_D,H_{D^*}}\right) \\ &\qquad \sim \text{Multinom}\left(H^{i,j,k,1}_D(t), \ q^{i,j,k}_{H_D,H_D}, \ q^{i,j,k,1,1}_{H_D,H_{D^*}}, \ q^{i,j,k,1,2}_{H_D,H_{D^*}}\right) \end{aligned} \tag{116}$$

$$d^{i,j,k}_{H_{D^*},H_{D^*}} \sim \text{Binom}\left(H^{i,j,k,1}_{D^*}(t), \ 1 - e^{-\gamma_{H_D}(t) dt}\right) \tag{117}$$

$$\left(d^{i,j,k}_{H_D,D}, \ d^{i,j,k,2,2}_{H_D,H_{D^*}}\right) \sim \text{Multinom}\left(H^{i,j,k,2}_D(t), \ 1 - e^{-\gamma_{H_D}(t) dt}, \ e^{-\gamma_{H_D}(t) dt}\left(1 - e^{-\gamma_U dt}\right)\right) \tag{118}$$

$$d^{i,j,k}_{H_{D^*},D} \sim \text{Binom}\left(H^{i,j,k,2}_{D^*}(t), \ 1 - e^{-\gamma_{H_D}(t) dt}\right) \tag{119}$$

$$\left(d^{i,j,k}_{H_R,R}, \ d^{i,j,k}_{H_R,H_{R^*}}\right) \sim \text{Multinom}\left(H^{i,j,k}_R(t), \ 1 - e^{-\gamma_{H_R}(t) dt}, \ e^{-\gamma_{H_R}(t) dt}\left(1 - e^{-\gamma_U dt}\right)\right) \tag{120}$$

$$d^{i,j,k}_{H_{R^*},R} \sim \text{Binom}\left(H^{i,j,k}_{R^*}(t), \ 1 - e^{-\gamma_{H_R}(t) dt}\right) \tag{121}$$

$$q^{i,j,k}_{ICU_{W_R},W_R} = \left(1 - e^{-\gamma_{ICU_{W_R}} dt}\right) e^{-\gamma_U dt} \tag{122}$$

$$q^{i,j,k}_{ICU_{W_R},ICU_{W_{R^*}}} = e^{-\gamma_{ICU_{W_R}} dt} \left(1 - e^{-\gamma_U dt}\right) \tag{123}$$

$$q^{i,j,k}_{ICU_{W_R},W_{R^*}} = \left(1 - e^{-\gamma_{ICU_{W_R}} dt}\right) \left(1 - e^{-\gamma_U dt}\right) \tag{124}$$

$$\begin{aligned} &\left(d^{i,j,k}_{ICU_{W_R},W_R}, \ \ldots, d^{i,j,k}_{ICU_{W_R},W_{R^*}}\right) \\ &\qquad \sim \text{Multinom}\left(ICU^{i,j,k}_{W_R}(t), \ q^{i,j,k}_{ICU_{W_R},W_R}, \ldots, q^{i,j,k}_{ICU_{W_R},W_{R^*}}\right) \end{aligned} \tag{125}$$

$$d^{i,j,k}_{ICU_{W_{R^*}},W_{R^*}} \sim \text{Binom}\left(ICU^{i,j,k}_{W_{R^*}}(t), \ 1 - e^{-\gamma_{ICU_{W_R}} dt}\right) \tag{126}$$

$$q^{i,j,k}_{ICU_{W_D},W_D} = \left(1 - e^{-\gamma_{ICU_{W_D}} dt}\right) e^{-\gamma_U dt} \tag{127}$$

$$q^{i,j,k}_{ICU_{W_D},ICU_{W_{D^*}}} = e^{-\gamma_{ICU_{W_D}} dt} \left(1 - e^{-\gamma_U dt}\right) \tag{128}$$

$$q^{i,j,k}_{ICU_{W_D},W_{D^*}} = \left(1 - e^{-\gamma_{ICU_{W_D}} dt}\right) \left(1 - e^{-\gamma_U dt}\right) \tag{129}$$

$$\begin{aligned} &\left(d^{i,j,k}_{ICU_{W_D},W_D}, \ \ldots, d^{i,j,k}_{ICU_{W_D},W_{D^*}}\right) \\ &\qquad \sim \text{Multinom}\left(ICU^{i,j,k}_{W_D}(t), \ q^{i,j,k}_{ICU_{W_D},W_D}, \ldots, q^{i,j,k}_{ICU_{W_D},W_{D^*}}\right) \end{aligned} \tag{130}$$

$$d^{i,j,k}_{ICU_{W_{D^*}},W_{D^*}} \sim \text{Binom}\left(ICU^{i,j,k}_{W_{D^*}}(t), \ 1 - e^{-\gamma_{ICU_{W_D}} dt}\right) \tag{131}$$

$$q_{ICU_D,ICU_D}^{i,j,k} = \left(1-e^{-\gamma_{ICU_D}dt}\right)e^{-\gamma_U dt} \tag{132}$$

$$q_{ICU_D,ICU_{D^*}}^{i,j,k,1,1} = e^{-\gamma_{ICU_D}dt}\left(1-e^{-\gamma_U dt}\right) \tag{133}$$

$$q_{ICU_D,ICU_{D^*}}^{i,j,k,1,2} = \left(1-e^{-\gamma_{ICU_D}dt}\right)\left(1-e^{-\gamma_U dt}\right) \tag{134}$$

$$\begin{aligned}&\left(d_{ICU_D,ICU_D}^{i,j,k},\ d_{ICU_D,ICU_{D^*}}^{i,j,k,1,1},\ d_{ICU_D,ICU_{D^*}}^{i,j,k,1,2}\right)\\ &\qquad\sim \text{Multinom}\left(ICU_D^{i,j,k,1}(t),\ q_{ICU_D,ICU_D}^{i,j,k},\ q_{ICU_D,ICU_{D^*}}^{i,j,k,1,1},\ q_{ICU_D,ICU_{D^*}}^{i,j,k,1,2}\right)\end{aligned} \tag{135}$$

$$d_{ICU_{D^*},ICU_{D^*}}^{i,j,k} \sim \text{Binom}\left(ICU_{D^*}^{i,j,k,1}(t),\ 1-e^{-\gamma_{ICU_D}dt}\right) \tag{136}$$

$$\left(q_{ICU_D,D}^{i,j,k},\ q_{ICU_D,ICU_{D^*}}^{i,j,k,2,2}\right) = \left(\ 1-e^{-\gamma_{ICU_D}dt},\ e^{-\gamma_{ICU_D}dt}\left(1-e^{-\gamma_U dt}\right)\right) \tag{137}$$

$$\left(d_{ICU_D,D}^{i,j,k},\ d_{ICU_D,ICU_{D^*}}^{i,j,k,2,2}\right) \sim \text{Multinom}\left(ICU_D^{i,j,k,2}(t),\ q_{ICU_D,D}^{i,j,k},\ q_{ICU_D,ICU_{D^*}}^{i,j,k,2,2}\right) \tag{138}$$

$$d_{ICU_{D^*},D}^{i,j,k} \sim \text{Binom}\left(ICU_{D^*}^{i,j,k,2}(t),\ 1-e^{-\gamma_{ICU_D}dt}\right) \tag{139}$$

$$q_{W_R,W_R}^{i,j,k} = \left(1-e^{-\gamma_{W_R}(t)dt}\right)e^{-\gamma_U dt} \tag{140}$$

$$q_{W_R,W_{R^*}}^{i,j,k,1,1} = e^{-\gamma_{W_R}(t)dt}\left(1-e^{-\gamma_U dt}\right) \tag{141}$$

$$q_{W_R,W_{R^*}}^{i,j,k,1,2} = \left(1-e^{-\gamma_{W_R}(t)dt}\right)\left(1-e^{-\gamma_U dt}\right) \tag{142}$$

$$\begin{aligned}&\left(d_{W_R,W_R}^{i,j,k},\ d_{W_R,W_{R^*}}^{i,j,k,1,1},\ d_{W_R,W_{R^*}}^{i,j,k,1,2}\right)\\ &\qquad\sim\ \text{Multinom}\left(W_R^{i,j,k,1}(t),\ q_{W_R,W_R}^{i,j,k},\ q_{W_R,W_{R^*}}^{i,j,k,1,1},\ q_{W_R,W_{R^*}}^{i,j,k,1,2}\right)\end{aligned} \tag{143}$$

$$d_{W_{R^*},W_{R^*}}^{i,j,k} \sim \text{Binom}\left(W_{R^*}^{i,j,k,1}(t),\ 1-e^{-\gamma_{W_R}(t)dt}\right) \tag{144}$$

$$\left(q_{W_R,R}^{i,j,k},\ q_{W_R,W_{R^*}}^{i,j,k,2,2}\right) = \left(1-e^{-\gamma_{W_R}(t)dt},\ e^{-\gamma_{W_R}(t)dt}\left(1-e^{-\gamma_U\ dt}\right)\right) \tag{145}$$

$$\left(d_{W_R,R}^{i,j,k},\ d_{W_R,W_{R^*}}^{i,j,k,2,2}\right) \sim \text{Multinom}\left(W_R^{i,j,k,2}(t),\ q_{W_R,R}^{i,j,k},\ q_{W_R,W_{R^*}}^{i,j,k,2,2}\right) \tag{146}$$

$$d_{W_{R^*},R}^{i,j,k} \sim \text{Binom}\left(W_{R^*}^{i,j,k,2}(t),\ 1-e^{-\gamma_{W_R}(t)dt}\right) \tag{147}$$

$$\left(q_{W_D,D}^{i,j,k},\ q_{W_D,W_{D^*}}^{i,j,k}\right) = \left(1-e^{-\gamma_{W_D}(t)dt},\ e^{-\gamma_{W_D}(t)dt}\left(1-e^{-\gamma_U dt}\right)\right) \tag{148}$$

$$\left(d_{W_D,D}^{i,j,k},\ d_{W_D,W_{D^*}}^{i,j,k}\right) \sim \text{Multinom}\left(W_D^{i,j,k}(t),\ q_{W_D,D}^{i,j,k},\ q_{W_D,W_{D^*}}^{i,j,k}\right) \tag{149}$$

$$d_{W_{D^*},D}^{i,j,k} \sim \text{Binom}\left(W_{D^*}^{i,j,k}(t),\ 1-e^{-\gamma_{W_D}(t)dt}\right) \tag{150}$$

$$\gamma_{R,E}^{i,j,k} = \mathbb{1}_{\{Alpha\}}(j)(1-\eta)\lambda^{i,Delta,k}(t) \tag{151}$$

$$q_{R,S}^{i,j,k} = \left(1-e^{-\left(\gamma_R+\gamma_{R,E}^{i,j,k}\right)dt}\right)\frac{\gamma_R}{\gamma_R+\gamma_{R,E}^{i,j,k}} \tag{152}$$

$$q_{R,E}^{i,j,k} = \left(1-e^{-\left(\gamma_R+\gamma_{R,E}^{i,j,k}\right)dt}\right)\frac{\gamma_{R,E}^{i,j,k}}{\gamma_R+\gamma_{R,E}^{i,j,k}} \tag{153}$$

$$q_{R,v}^{i,j,k} = e^{-\left(\gamma_R+\gamma_{R,E}^{i,j,k}\right)dt}\left(1-e^{-\zeta^{i,k}(t)dt}\right) \tag{154}$$

$$\left(d_{R,S}^{i,j,k},\, d_{R,E}^{i,j,k},\, d_{R,v}^{i,j,k}\right) \sim \text{Multinom}\left(R^{i,j,k}(t),\, q_{R,S}^{i,j,k},\, q_{R,E}^{i,j,k},\, q_{R,v}^{i,j,k}\right) \tag{155}$$

$$q^i_{T_{sero^1_{pre}},T_{sero^1_{pos}}} = p_{sero_{pos}}\left(1-e^{-\gamma_{sero_{pre}}dt}\right) \tag{156}$$

$$q^i_{T_{sero^1_{pre}},T_{sero^1_{neg}}} = \left(1-p_{sero_{pos}}\right)\left(1-e^{-\gamma_{sero_{pre}}dt}\right) \tag{157}$$

$$\begin{aligned}\left(d^i_{T_{sero^1_{pre}},T_{sero^1_{pos}}},\, d^i_{T_{sero^1_{pre}},T_{sero^1_{neg}}}\right)& \\ \sim \text{Multinom}&\left(T^i_{sero^1_{pre}}(t),\, q^i_{T_{sero^1_{pre}},T_{sero^1_{pos}}},\, q^i_{T_{sero^1_{pre}},T_{sero^1_{neg}}}\right)\end{aligned} \tag{158}$$

$$d^i_{T_{sero^1_{pos}},T_{sero^1_{neg}}} \sim \text{Binom}\left(T^i_{sero^1_{pos}}(t),\, 1-e^{-\gamma_{sero^1_{pos}}dt}\right) \tag{159}$$

$$q^i_{T_{sero^2_{pre}},T_{sero^2_{pos}}} = p_{sero_{pos}}\left(1-e^{-\gamma_{sero_{pre}}dt}\right) \tag{160}$$

$$q^i_{T_{sero^2_{pre}},T_{sero^2_{neg}}} = \left(1-p_{sero_{pos}}\right)\left(1-e^{-\gamma_{sero_{pre}}dt}\right) \tag{161}$$

$$\begin{aligned}\left(d^i_{T_{sero^2_{pre}},T_{sero^2_{pos}}},\, d^i_{T_{sero^2_{pre}},T_{sero^2_{neg}}}\right)& \\ \sim \text{Multinom}&\left(T^i_{sero^2_{pre}}(t),\, q^i_{T_{sero^2_{pre}},T_{sero^2_{pos}}},\, q^i_{T_{sero^2_{pre}},T_{sero^2_{neg}}}\right)\end{aligned} \tag{162}$$

$$d^i_{T_{sero^2_{pos}},T_{sero^2_{neg}}} \sim \text{Binom}\left(T^i_{sero^2_{pos}}(t),\, 1-e^{-\gamma_{sero^2_{pos}}dt}\right) \tag{163}$$

$$d^i_{T_{PCR_{pre}},T_{PCR_{pos}}} \sim \text{Binom}\left(T^i_{PCR_{pre}}(t),\, 1-e^{-\gamma_{PCR_{pre}}dt}\right) \tag{164}$$

$$d^i_{T_{PCR_{pos}},T_{PCR_{neg}}} \sim \text{Binom}\left(T^i_{PCR_{pos}}(t),\, 1-e^{-\gamma_{PCR_{pos}}dt}\right) \tag{165}$$

Model compartments were then updated as follows (Note that $d_{S,E}^{i,Alpha\to Delta,k} = 0$):

$$S^{i,k}(t+dt) := S^{i,k}(t) - d_{S,E}^{i,Alpha,k} - d_{S,E}^{i,Delta,k} + d_{R,S}^{i,j,k} + d_{S,v}^{i,k-1} - d_{S,v}^{i,k} \tag{166}$$

$$E^{i,j,k,1}(t+dt) := E^{i,j,k,1}(t) + d_{S,E}^{i,j,k} + \mathbb{1}_{\{Alpha\to Delta\}}(j)d_{R,E}^{i,Alpha,k} - d_{E,E}^{i,j,k} + d_{E,v}^{i,j,k-1,1} - d_{E,v}^{i,j,k,1} \tag{167}$$

$$E^{i,j,k,2}(t+dt) := E^{i,j,k,2}(t) + d_{E,E}^{i,j,k} - d_{E,I_A}^{i,j,k} - d_{E,I_P}^{i,j,k} + d_{E,v}^{i,j,k-1,2} - d_{E,v}^{i,j,k,2} \tag{168}$$

$$I_A^{i,j,k}(t+dt) := I_A^{i,j,k}(t) + d_{E,I_A}^{i,j,k} - d_{I_A,R}^{i,j,k} + d_{I_A,v}^{i,j,k-1} - d_{I_A,v}^{i,j,k} \tag{169}$$

$$I_P^{i,j,k}(t+dt) := I_P^{i,j,k}(t) + d_{E,I_P}^{i,j,k} - d_{I_P,I_{C_1}}^{i,j,k} + d_{I_P,v}^{i,j,k-1} - d_{I_P,v}^{i,j,k} \tag{170}$$

$$I_{C_1}^{i,j,k}(t+dt) := I_{C_1}^{i,j,k}(t) + d_{I_P,I_{C_1}}^{i,j,k} - d_{I_{C_1},I_{C_2}}^{i,j,k} \tag{171}$$

$$\begin{aligned}I_{C_2}^{i,j,k}(t+dt) := I_{C_2}^{i,j,k}(t) &+ d_{I_{C_1},I_{C_2}}^{i,j,k} - d_{I_{C_2},G_D}^{i,j,k} - d_{I_{C_2},R}^{i,j,k} - d_{I_{C_2},ICU_{pre}}^{i,j,k} \\ &- d_{I_{C_2},ICU_{pre^*}}^{i,j,k} - d_{I_{C_2},H_R}^{i,j,k} - d_{I_{C_2},H_{R^*}}^{i,j,k} - d_{I_{C_2},H_D}^{i,j,k} - d_{I_{C_2},H_{D^*}}^{i,j,k}\end{aligned} \tag{172}$$

$$G_D^{i,j,k,1}(t+dt) := G_D^{i,j,k,1}(t) + d_{I_{C_2},G_D}^{i,j,k} - d_{G_D,G_D}^{i,j,k} \tag{173}$$

$$G_D^{i,j,k,2}(t+dt) := G_D^{i,j,k,2}(t) + d_{G_D,G_D}^{i,j,k} - d_{G_D,D}^{i,j,k} \tag{174}$$

$$
\begin{aligned}
ICU_{pre}^{i,j,k}(t+dt) &:= ICU_{pre}^{i,j,k}(t) + d^{i,j,k}_{I_{C_2},ICU_{pre}} - d^{i,j,k}_{ICU_{pre},ICU_{W_R}} - d^{i,j,k}_{ICU_{pre},ICU_{W_D}} \\
&\quad - d^{i,j,k}_{ICU_{pre},ICU_D} - d^{i,j,k}_{ICU_{pre},ICU_{pre^*}} - d^{i,j,k}_{ICU_{pre},ICU_{W_{R^*}}} - d^{i,j,k}_{ICU_{pre},ICU_{W_{D^*}}} \\
&\quad - d^{i,j,k}_{ICU_{pre},ICU_{D^*}}
\end{aligned} \tag{175}
$$

$$
\begin{aligned}
ICU_{pre^*}^{i,j,k}(t+dt) &:= ICU_{pre^*}^{i,j,k}(t) + d^{i,j,k}_{I_{C_2},ICU_{pre^*}} - d^{i,j,k}_{ICU_{pre},ICU_{W_{D^*}}} - d^{i,j,k}_{ICU_{pre^*},ICU_{W_{R^*}}} \\
&\quad - d^{i,j,k}_{ICU_{pre^*},ICU_{D^*}}
\end{aligned} \tag{176}
$$

$$
\begin{aligned}
ICU_{W_R}^{i,j,k}(t+dt) &:= ICU_{W_R}^{i,j,k}(t) + d^{i,j,k}_{ICU_{pre},ICU_{W_R}} - d^{i,j,k}_{ICU_{W_R},W_R} - d^{i,j,k}_{ICU_{W_R},ICU_{W_{R^*}}} \\
&\quad - d^{i,j,k}_{ICU_{W_R},W_{R^*}}
\end{aligned} \tag{177}
$$

$$
\begin{aligned}
ICU_{W_{R^*}}^{i,j,k}(t+dt) &:= ICU_{W_{R^*}}^{i,j,k}(t) + d^{i,j,k}_{ICU_{pre^*},ICU_{W_{R^*}}} + d^{i,j,k}_{ICU_{W_R},ICU_{W_{R^*}}} + d^{i,j,k}_{ICU_{pre},ICU_{W_{R^*}}} \\
&\quad - d^{i,j,k}_{ICU_{W_{R^*}},W_{R^*}}
\end{aligned} \tag{178}
$$

$$
\begin{aligned}
ICU_{W_D}^{i,j,k}(t+dt) &:= ICU_{W_D}^{i,j,k}(t) + d^{i,j,k}_{ICU_{pre},ICU_{W_D}} - d^{i,j,k}_{ICU_{W_D},W_D} - d^{i,j,k}_{ICU_{W_D},ICU_{W_{D^*}}} \\
&\quad - d^{i,j,k}_{ICU_{W_D},W_{D^*}}
\end{aligned} \tag{179}
$$

$$
\begin{aligned}
ICU_{W_{D^*}}^{i,j,k}(t+dt) &:= ICU_{W_{D^*}}^{i,j,k}(t) + d^{i,j,k}_{ICU_{pre^*},ICU_{W_{D^*}}} + d^{i,j,k}_{ICU_{W_D},ICU_{W_{D^*}}} + d^{i,j,k}_{ICU_{pre},ICU_{W_{D^*}}} \\
&\quad - d^{i,j,k}_{ICU_{W_{D^*}},W_{D^*}}
\end{aligned} \tag{180}
$$

$$
\begin{aligned}
ICU_{D}^{i,j,k,1}(t+dt) &:= ICU_{D}^{i,j,k,1}(t) + d^{i,j,k}_{ICU_{pre},ICU_D} - d^{i,j,k}_{ICU_D,ICU_D} - d^{i,j,k,1,1}_{ICU_D,ICU_{D^*}} \\
&\quad - d^{i,j,k,1,2}_{ICU_D,ICU_{D^*}}
\end{aligned} \tag{181}
$$

$$
ICU_{D}^{i,j,k,2}(t+dt) := ICU_{D}^{i,j,k,2}(t) + d^{i,j,k}_{ICU_D,ICU_D} - d^{i,j,k}_{ICU_D,D} - d^{i,j,k,2,2}_{ICU_D,ICU_{D^*}} \tag{182}
$$

$$
\begin{aligned}
ICU_{D^*}^{i,j,k,1}(t+dt) &:= ICU_{D^*}^{i,j,k,1}(t) + d^{i,j,k}_{ICU_{pre^*},ICU_{D^*}} + d^{i,j,k,1,1}_{ICU_D,ICU_{D^*}} + d^{i,j,k}_{ICU_{pre},ICU_{D^*}} \\
&\quad - d^{i,j,k}_{ICU_{D^*},ICU_{D^*}}
\end{aligned} \tag{183}
$$

$$
\begin{aligned}
ICU_{D^*}^{i,j,k,2}(t+dt) &:= ICU_{D^*}^{i,j,k,2}(t) + d^{i,j,k}_{ICU_{D^*},ICU_{D^*}} + d^{i,j,k,1,2}_{ICU_D,ICU_{D^*}} + d^{i,j,k,2,2}_{ICU_D,ICU_{D^*}} \\
&\quad - d^{i,j,k}_{ICU_{D^*},D}
\end{aligned} \tag{184}
$$

$$
W_R^{i,j,k,1}(t+dt) := W_R^{i,j,k,1}(t) + d^{i,j,k}_{ICU_{W_R},W_R} - d^{i,j,k}_{W_R,W_R} - d^{i,j,k,1,1}_{W_R,W_{R^*}} - d^{i,j,k,1,2}_{W_R,W_{R^*}} \tag{185}
$$

$$
W_R^{i,j,k,2}(t+dt) := W_R^{i,j,k,2}(t) + d^{i,j,k}_{W_R,W_R} - d^{i,j,k}_{W_R,R} - d^{i,j,k,2,2}_{W_R,W_{R^*}} \tag{186}
$$

$$
W_{R^*}^{i,j,k,1}(t+dt) := W_{R^*}^{i,j,k,1}(t) + d^{i,j,k}_{ICU_{W_{R^*}},W_{R^*}} + d^{i,j,k,1,1}_{W_R,W_{R^*}} + d^{i,j,k}_{ICU_{W_R},W_{R^*}} - d^{i,j,k}_{W_{R^*},W_{R^*}} \tag{187}
$$

$$
W_{R^*}^{i,j,k,2}(t+dt) := W_{R^*}^{i,j,k,2}(t) + d^{i,j,k}_{W_{R^*},W_{R^*}} + d^{i,j,k,2,2}_{W_R,W_{R^*}} + d^{i,j,k,1,2}_{W_R,W_{R^*}} - d^{i,j,k}_{W_{R^*},R} \tag{188}
$$

$$
W_D^{i,j,k}(t+dt) := W_D^{i,j,k}(t) + d^{i,j,k}_{ICU_{W_D},W_D} - d^{i,j,k}_{W_D,D} - d^{i,j,k}_{W_D,W_{D^*}} \tag{189}
$$

$$
W_{D^*}^{i,j,k}(t+dt) := W_{D^*}^{i,j,k}(t) + d^{i,j,k}_{ICU_{W_{D^*}},W_{D^*}} + d^{i,j,k}_{W_D,W_{D^*}} + d^{i,j,k}_{ICU_{W_D},W_{D^*}} - d^{i,j,k}_{W_{D^*},D} \tag{190}
$$

$$
H_D^{i,1}(t+dt) := H_D^{i,j,k,1}(t) + d^{i,j,k}_{I_{C_2},H_D} - d^{i,j,k}_{H_D,H_D} - d^{i,j,k,1,1}_{H_D,H_{D^*}} - d^{i,j,k,1,2}_{H_D,H_{D^*}} \tag{191}
$$

$$
H_D^{i,j,k,2}(t+dt) := H_D^{i,j,k,2}(t) + d^{i,j,k}_{H_D,H_D} - d^{i,j,k}_{H_D,D} - d^{i,j,k,2,2}_{H_D,H_{D^*}} \tag{192}
$$

$$
H_{D^*}^{i,j,k,1}(t+dt) := H_{D^*}^{i,j,k,1}(t) + d^{i,j,k}_{I_{C_2},H_{D^*}} + d^{i,j,k,1,1}_{H_D,H_{D^*}} - d^{i,j,k}_{H_{D^*},H_{D^*}} \tag{193}
$$

$$H_{D^*}^{i,j,k,2}(t+dt) := H_{D^*}^{i,j,k,2}(t) + d_{H_{D^*},H_{D^*}}^{i,j,k} + d_{H_D,H_{D^*}}^{i,j,k,2,2} + d_{H_D,H_{D^*}}^{i,j,k,1,2} - d_{H_{D^*},D}^{i,j,k} \tag{194}$$

$$H_R^{i,j,k}(t+dt) := H_R^{i,j,k}(t) + d_{I_{C_2},H_R}^{i,j,k} - d_{H_R,R}^{i,j,k} - d_{H_R,H_{R^*}}^{i,j,k} \tag{195}$$

$$H_{R^*}^{i}(t+dt) := H_{R^*}^{i,j,k}(t) + d_{I_{C_2},H_{R^*}}^{i,j,k} + d_{H_R,H_{R^*}}^{i,j,k} - d_{H_{R^*},R}^{i,j,k} \tag{196}$$

$$\begin{aligned} R^{i,j,k}(t+dt) := {} & R^{i,j,k}(t) + d_{I_A,R}^{i,j,k} + d_{I_{C_2},R}^{i,j,k} + d_{H_R,R}^{i,j,k} + d_{H_{R^*},R}^{i,j,k} + d_{W_R,R}^{i,j,k} + d_{W_{R^*},R}^{i,j,k} \\ & - d_{R,S}^{i,j,k} - \mathbb{1}_{\{Alpha\}} d_{R,E}^{i,Alpha,k} + d_{R,v}^{i,j,k-1} - d_{R,v}^{i,j,k} \end{aligned} \tag{197}$$

$$T_{sero^1_{pre}}^{i}(t+dt) := T_{sero^1_{pre}}^{i}(t) - d_{T_{sero^1_{pre}},T_{sero^1_{pos}}}^{i} - d_{T_{sero^1_{pre}},T_{sero^1_{neg}}}^{i} + \sum_j \sum_k d_{E,I_A}^{i,j,k} + d_{E,I_P}^{i,j,k} \tag{198}$$

$$T_{sero^1_{pos}}^{i}(t+dt) := T_{sero^1_{pos}}^{i}(t) + d_{T_{sero^1_{pre}},T_{sero^1_{pos}}}^{i} - d_{T_{sero^1_{pos}},T_{sero^1_{neg}}}^{i} \tag{199}$$

$$T_{sero^1_{neg}}^{i}(t+dt) := T_{sero^1_{neg}}^{i}(t) + d_{T_{sero^1_{pre}},T_{sero^1_{neg}}}^{i} + d_{T_{sero^1_{pos}},T_{sero^1_{neg}}}^{i} \tag{200}$$

$$T_{sero^2_{pre}}^{i}(t+dt) := T_{sero^2_{pre}}^{i}(t) - d_{T_{sero^2_{pre}},T_{sero^2_{pos}}}^{i} - d_{T_{sero^2_{pre}},T_{sero^2_{neg}}}^{i} + \sum_j \sum_k d_{E,I_A}^{i,j,k} + d_{E,I_P}^{i,j,k} \tag{201}$$

$$T_{sero^2_{pos}}^{i}(t+dt) := T_{sero^2_{pos}}^{i}(t) + d_{T_{sero^2_{pre}},T_{sero^2_{pos}}}^{i} - d_{T_{sero^2_{pos}},T_{sero^2_{neg}}}^{i} \tag{202}$$

$$T_{sero^2_{neg}}^{i}(t+dt) := T_{sero^2_{neg}}^{i}(t) + d_{T_{sero^2_{pre}},T_{sero^2_{neg}}}^{i} + d_{T_{sero^2_{pos}},T_{sero^2_{neg}}}^{i} \tag{203}$$

$$T_{PCR_{pre}}^{i}(t+dt) := T_{PCR_{pre}}^{i}(t) - d_{T_{PCR_{pre}},T_{PCR_{pos}}}^{i} + \sum_j \sum_k d_{S,E}^{i,j,k} \tag{204}$$

$$T_{PCR_{pos}}^{i}(t+dt) := T_{PCR_{pos}}^{i}(t) + d_{T_{PCR_{pre}},T_{PCR_{pos}}}^{i} - d_{T_{PCR_{pos}},T_{PCR_{neg}}}^{i} \tag{205}$$

$$T_{PCR_{neg}}^{i}(t+dt) := T_{PCR_{neg}}^{i}(t) + d_{T_{PCR_{pos}},T_{PCR_{neg}}}^{i} \tag{206}$$

$$\tag{207}$$

##### 4.3 Reproduction number

Both $R^j(t)$ and $R_e^j(t)$ are calculated using next generation matrix (NGM) methods [37], and only consider mixing in the general population, i.e. we do not consider the $CHW$ and $CHR$ age categories.

Note that in this calculation only, we make a simplifying assumption that individuals cannot change vaccine strata between initial infection and the end of their infectious period (or death).

To compute the next generation matrix, we calculated the mean duration of infectiousness weighted by infectivity (asymptomatic individuals are less infectious than symptomatic individuals by factor $\theta_{I_A}$) for an individual in group $i$ and vaccine stage $k$, $\Delta_I^{i,k}$:

$$\Delta_I^{i,k} = \theta_{I_A}\left(1 - p_C^{i,j,k}\right)\mathbb{E}[\tau_{I_A}] + p_C^{i,j,k}\left(\mathbb{E}[\tau_{I_P}] + \mathbb{E}\left[\tau_{I_{C_1}}\right]\right). \tag{208}$$

Note that $\Delta_I^{i,k}$ does not depend on $j$, as we assume the same duration spent in compartments and probability of being symptomatic between variants. The next generation matrices for the variants were calculated as,

$$NGM_{i,i'}^{Alpha}(t) = m_{i,i'}(t)\Delta_I^{i,0}N^{i'}, \tag{209}$$

$$NGM_{i,i'}^{Delta}(t) = m_{i,i'}(t)\xi^{i,Delta,0}\Delta_I^{i,0}N^{i'}, \tag{210}$$

where $\xi$ is the infectivity of an individual (fully defined in eq. (2)), $N^i$ is the total population of age group $i$, and with $R^j(t)$ taken as the dominant eigenvalue of the 17 by 17 matrix $NGM^j(t)$. The element $NGM^j_{i,i'}(t)$ is therefore defined as the average number of secondary cases that an individual in age group $i'$ infected with variant $j$ at time $t$ would generate among age group $i$.

The effective next generation matrices for the variants were calculated as

$$NGM^{Alpha,e}_{D(i,k),D(i',k')}(t) = m_{i,i'}(t)\chi^{i,Alpha,k}\xi^{i,Alpha,k'}\Delta^{i,k}_I S^{i',k'}(t), \tag{211}$$

$$NGM^{Delta,e}_{D(i,k),D(i',k')}(t) = m_{i,i'}(t)\chi^{i,Delta,k}\xi^{i,Delta,k'}\Delta^{i,k}_I \left(S^{i',k'}(t) + (1-\eta)R^{i',Alpha,k'}(t)\right), \tag{212}$$

where $D : \{[0-4],[5-9],\ldots,[75-79],80+\} \times \{0,1,2,3\} \to \{1,2,\ldots,68\}$ is a one-to-one mapping. Then $R^j_e(t)$ is taken as the dominant eigenvalue of the 68 by 68 matrix $NGM^{j,e}(t)$ .

We calculate the reproduction numbers weighted by the two variants as

$$R(t) = \frac{w_{Alpha}(t)R^{Alpha}(t) + w_{Delta}(t)R^{Delta}(t)}{w_{Alpha}(t) + w_{Delta}(t)} \tag{213}$$

$$R_e(t) = \frac{w_{Alpha}(t)R^{Alpha}_e(t) + w_{Delta}(t)R^{Delta}_e(t)}{w_{Alpha}(t) + w_{Delta}(t)}, \tag{214}$$

where the weightings $w_j(t)$ are weightings based on the infectious prevalence of each variant (accounting for the baseline relative infectivity of each compartment), such that for $j = Alpha, Delta$,

$$w_j(t) = \sum_i \sum_k \left(\theta_{I_A} I^{i,j,k}_A(t) + I^{i,j,k}_P(t) + I^{i,j,k}_{C_1}(t)\right). \tag{215}$$

##### 4.4 Fixed parameters

We used parameter values calibrated to data from 19 July 2021.

| Parameter | Definition | Value | Source |
|---|---|---|---|
| $1/\gamma_U$ | Mean time to confirmation of SARS-CoV-2 diagnosis within hospital. | 2 days | [38] |
| $1/\gamma_R$ | Mean duration of natural immunity following infection. | 3, 6, $\infty$ years | [39], (sensitivity) |
| $p_{sero_{pos}}$ | Probability of seroconversion following infection. | 0.85 | [40] |
| $1/\gamma_{sero_{pre}}$ | Mean time to seroconversion from onset of infectiousness. | 13 days | [41] |
| $1/\gamma_{sero^1_{pos}}$ | Mean duration of seropositivity (Euroimmun assay). | 200 days | [40, 42, 43] |
| $1/\gamma_{sero^2_{pos}}$ | Mean duration of seropositivity (Roche N). | 400 days | [40, 42, 43] |
| $p_{sero_{spec}}$ | Specificity of serology test. | 0.99 | [40] |
| $p_{sero_{sens}}$ | Sensitivity of serology test. | 1 | Assumed |
| $\eta$ | Proportion of cross-immunity to $Delta$ following infection with $Alpha$. | 0.75,0.85,1 | [44], (sensitivity) |
| $\theta_{I_A}$ | Infectivity of an asymptomatic individual, relative to a symptomatic individual. | 0.223 | [1] |

**Table S7:** Fixed model parameter notations, values, and evidence-base.

##### 4.5 Prior distributions

**Table S8:** Inferred model parameter notations and prior distributions. Region codes: NW = North West, NEY = North East and Yorkshire, MID = Midlands, EE = East of England, LON = London, SW = South West, SE = South East

| | Description | Region | Prior distribution |
|---|---|---|---|
| $t_0$ | Start date of regional outbreak (2020-mm-dd) | All | $U[01-01, 03-15]$ |
| $t_{Delta}$ | Delta seeding date (2021-mm-dd) | All | $U[03-10, 05-31]$ |
| $\sigma$ | Delta transmission advantage | All | $U(0,3)$ |
| $\beta(t)$ | Transmission rate (pp) at $t = dd/mm/yy$ | | |
| $\beta_1$ | 16/03/20: PM advises WFH and essential travel only | All | $\Gamma(136, 0.0008)$ |
| $\beta_2$ | 23/03/20: PM announces lockdown 1 | All | $\Gamma(3.73, 0.0154)$ |
| $\beta_3$ | 25/03/20: Lockdown 1 into full effect | All | $\Gamma(4.25, 0.0120)$ |
| $\beta_4$ | 11/05/20: Initial easing of lockdown 1 | All | $\Gamma(4.25, 0.0120)$ |
| $\beta_5$ | 15/06/20: Non-essential shops re-open | All | $\Gamma(4.25, 0.0120)$ |
| $\beta_6$ | 04/07/20: Hospitality re-opens | All | $\Gamma(4.25, 0.0120)$ |
| $\beta_7$ | 01/08/20: “Eat out to help out” scheme starts | All | $\Gamma(4.25, 0.0120)$ |
| $\beta_8$ | 01/09/20: Schools and universities re-open | All | $\Gamma(4.25, 0.0120)$ |
| $\beta_9$ | 14/09/20: “Rule of six” introduced | All | $\Gamma(4.25, 0.0120)$ |
| $\beta_{10}$ | 14/10/20: Tiered system introduced | All | $\Gamma(4.25, 0.0120)$ |
| $\beta_{11}$ | 31/10/20: Lockdown 2 announced | All | $\Gamma(4.25, 0.0120)$ |
| $\beta_{12}$ | 05/11/20: Lockdown 2 starts | All | $\Gamma(4.25, 0.0120)$ |
| $\beta_{13}$ | 02/12/20: Lockdown 2 ends | All | $\Gamma(4.25, 0.0120)$ |
| $\beta_{14}$ | 18/12/20: School holidays start | All | $\Gamma(4.25, 0.0120)$ |
| $\beta_{15}$ | 25/12/20: Last day of holiday season relaxation | All | $\Gamma(4.25, 0.0120)$ |
| $\beta_{16}$ | 05/01/21: Lockdown 3 starts | All | $\Gamma(4.25, 0.0120)$ |
| $\beta_{17}$ | 08/03/21: Roadmap step one - schools reopen | All | $\Gamma(4.25, 0.0120)$ |
| $\beta_{18}$ | 01/04/21: School holidays | All | $\Gamma(4.25, 0.0120)$ |
| $\beta_{19}$ | 19/04/21: Roadmap step two - outdoor rule of 6 (12/04) and schools re-open (19/04) | All | $\Gamma(2.72, 0.0292)$ |
| $\beta_{20}$ | 17/05/21: Roadmap step three - Indoor hospitality opens | All | $\Gamma(2.72, 0.0292)$ |
| $\beta_{21}$ | 21/06/21: Wedding and care home restrictions eased | All | $\Gamma(2.72, 0.0292)$ |
| $\beta_{22}$ | 03/07/21: Euro 2020 quarter finals (cited as significant influence [45]) | All | $\Gamma(2.72, 0.0292)$ |
| $\beta_{23}$ | 11/07/21: End of Euros football tournament | All | $\Gamma(2.72, 0.0292)$ |
| $\beta_{24}$ | 19/07/21: Full lift of NPIs | All | $\Gamma(2.72, 0.0292)$ |
| $\varepsilon$ | Relative reduction in contacts between CHR and the general population | All | $B(1,1)$ |
| $m_{CHW}$ | Transmission rate between CHR and CHW | NW | $\Gamma(5, 4.3 \times 10^{-7})$ |
| | | NEY | $\Gamma(5, 3.7 \times 10^{-7})$ |
| | | MID | $\Gamma(5, 2.9 \times 10^{-7})$ |
| | | EE | $\Gamma(5, 5.2 \times 10^{-7})$ |
| | | LON | $\Gamma(5, 7.6 \times 10^{-7})$ |

Continued on next page

| **Table S8 – continued from previous page** | | | |
|---|---|---|---|
| | Description | Region | Prior distribution |
| | | SW | $\Gamma(5, 4.9\times10^{-7})$ |
| | | SE | $\Gamma(5, 3.1\times10^{-7})$ |
| $m_{CHR}$ | Transmission rate among CHR | NW | $\Gamma(5, 4.3\times10^{-7})$ |
| | | NEY | $\Gamma(5, 3.7\times10^{-7})$ |
| | | MID | $\Gamma(5, 2.9\times10^{-7})$ |
| | | EE | $\Gamma(5, 5.2\times10^{-7})$ |
| | | LON | $\Gamma(5, 7.6\times10^{-7})$ |
| | | SW | $\Gamma(5, 4.9\times10^{-7})$ |
| | | SE | $\Gamma(5, 3.1\times10^{-7})$ |
| $p_{H,1}^{max}, p_{H,2}^{max}, p_{H,3}^{max}$ | The probability of symptomatic individuals developing serious disease requiring hospitalisation, for the group with the largest probability at different timepoints (see Section 4.2.2) | All | $B(15.8, 5.28)$ |
| $p_{G_D}^{max}$ | Probability of death in the community given disease severe enough for hospitalisation | All | $B(1,1)$ |
| $p_{G_D}^{CHR}$ | Probability of death in CHR given disease severe enough for hospitalisation | All | $B(1,1)$ |
| $p_{ICU,1}^{max}, p_{ICU,2}^{max}$ | Probability of triage to ICU for new hospital admissions, for the group with the largest probability at different timepoints (see Section 4.2.2) | All | $B(13.9, 43.9)$ |
| $p_{H_D}^{max}$ | Initial probability of death for general inpatients | All | $B(42.1, 50.1)$ |
| $p_{ICU_D}^{max}$ | Initial probability of death for ICU inpatients | All | $B(60.2, 29.3)$ |
| $p_{W_D}^{max}$ | Initial probability of death for stepdown inpatients | All | $B(28.7, 52.1)$ |
| $\mu_{D,1}, \mu_{D,2}$ | Hospital mortality multipliers due to changes in clinical care at different timepoints (see Section 4.2.2) | All | $B(1,1)$ |
| $\mu_{\gamma_H,1}, \mu_{\gamma_H,2}, \mu_{\gamma_H,3}$ | Mean duration multipliers for non-ICU hospital compartments at different timepoints (see Section 4.2.2) | All | $B(1,1)$ |
| $p_{NC}$ | Prevalence of non-COVID symptomatic illness that could lead to getting a PCR test | All | $B(1,1)$ |
| $p_{NC}^{weekend}$ | Prevalence of non-COVID symptomatic illness that could lead to getting a PCR test on a weekend | All | $B(1,1)$ |
| $\rho_{P2_{test}}$ | Overdispersion of PCR positivity | All | $B(1,1)$ |
| $\alpha_H$ | Overdispersion for hospital data streams | All | $B(1,1)$ |
| $\alpha_D$ | Overdispersion for death data streams | All | $B(1,1)$ |

##### 4.6 Running the model

The model is run three times to cover central, optimistic, and pessimistic scenarios. These are reflected in the fixed parameters (Table S8) and VE (Table S3) tables. Specifically these scenarios are given by setting all fixed parameters above with:

| Assumptions | Cross-immunity, $\eta$ | Mean duration natural immunity, $1/\gamma_R$ | Vaccine effectiveness vs Delta |
|---|---|---|---|
| Optimistic | 1 | $\infty$ | optimistic (Table S3) |
| Central | 0.85 | 6 years | central (Table S3) |
| Pessimistic | 0.75 | 3 years | pessimistic (Table S3) |

**Table S9:** Combinations of parameters that are run in the model in three distinct scenarios. See Table S8 for details of the parameters and Table S3 for the vaccine effectiveness scenarios.

#### 5 Forward Projections

The previous sections provided methodological details of our model structure and how we fit to existing epidemiological data. This section outlines information and assumptions made for simulating forward projections from the fitted model. Simulated scenarios depend primarily on assumptions regarding future transmissibility and vaccine rollout, which we discuss in sections 5.1 and 5.2 respectively. The range of simulated scenarios that we run, including counterfactual runs, are detailed in Section 5.3.

The simulated model is run three times, corresponding to the three model fits from the central/optimistic/pessimistic assumptions (Section 4.6). These assumptions are carried forward into the simulated model, e.g. optimistic assumptions are made in the simulation model when using the optimistic model fits (same for central and pessimistic). As sensitivity analyses we vary these assumptions in the simulations for each of the three model fits (Section 5.4).

##### 5.1 Transmissibility of SARS-CoV-2

There is considerable uncertainty on the level of transmissibility of SARS-CoV-2 following the final stage of NPI lifting, 'step four'. The overall reproduction number will depend on the relative frequency of each SARS-CoV-2 variant in circulation, the ability of the virus to be transmitted upon contact between two individuals (which may depend on environmental factors such as temperature therefore displaying seasonal effects), the level of physical mixing between individuals (which could be low if individuals do not immediately return to their pre-pandemic behaviours, or could be high if individuals suddenly compensate for months of low mixing), and the level of susceptibility in the population.

###### 5.1.1 NPI lifting: the new baseline

The final level of transmissibility once all NPIs are lifted is determined by the assumptions made in the given simulation scenario. To estimate the value of $R^{Alpha}(t)$ in Step 4 we took the average $R^{Alpha}(t)$ over Step 3 in June and July then multiplied this value by 1.4, 1.7, or 2 to correspond with an optimistic, central, or pessimistic contact mixing assumption. Table S10 summarises the estimated average $R^{Alpha}(t)$ values in Step 3 and assumed values in Step 4 for each of the three contact mixing assumptions, as well as for the three model assumptions (Section 4.6). By averaging over all of June and July, the same values for Step 4 can be utilised in all scenarios (independent of the full lift date).

The estimated level of transmissibility depends on the assumptions around VE, immunity and cross protection. In the optimistic scenario (Table S10 row 1), where VE and cross-protection are high and immunity is long-lasting, a high level of transmission is required to explain the level of observed cases. Conversely, in the pessimistic scenario (Table S10 row 3), a lower level of transmission can explain the observed number of cases, due to low VE and cross-protection, as well as immunity waning after 3 years.

| Model assumption | Average $R^{Alpha}_{S3}(t)$ | Assumed $R^{Alpha}_{S4}(t)$ | Average $R^{Delta}_{S3}(t)$ | Assumed $R^{Delta}_{S4}(t)$ |
|---|---|---|---|---|
| Optimistic | 1.74 | {2.44, 2.96, 3.49} | 3.43 | {4.81, 5.84, 6.87} |
| Central | 1.59 | {2.23, 2.70, 3.18} | 2.82 | {3.95, 4.80, 5.65} |
| Pessimistic | 1.45 | {2.02, 2.46, 2.89} | 2.41 | {3.37, 4.09, 4.81} |

**Table S10:** Forward projection scenarios with $R^j(t)$ values based on: model assumptions (column 1) (Section 4.6), average $R^{Alpha}(t)$ in Step 3 between 2021-06-01 and 2021-07-31 (column 3) and same for $R^{Delta}(t)$ (column 5), $R^{Alpha}(t)$ in Step 4 (column 4) and $R^{Delta}(t)$ in Step 4 (column 6) under different contact mixing assumptions (1.4x, 1.7x, 2x Step 3).

$R^j(t)$ is assumed to vary through time in our forward projections, in accordance with school term dates and seasonal variation. To capture this variation we assumed $R^j(t)$ under each level of restrictions was distributed lognormally with means as given in Table S10, and assume a standard deviation of 0.2. For each NPI lifting scenario, we sampled from the relevant distributions of $R^j(t)$ and generated sampled trajectories of $R^j(t)$ over time by matching the ranked values obtained for each step, where distinct steps are defined by the opening and closing of schools as defined in Table S11 below. This constraint was added to ensure that $R^j(t)$ could only increase over time except for decreases due to school closures and seasonality.

##### 5.1.2 School closures

School closures can break the chains of transmission between households and reduces contacts between children, and children and adults. This can therefore reduce the level of transmission of respiratory pathogens such as SARS-CoV-2. We accounted for this by assuming that during school holidays $R^j(t)$ decreases by 0.25 compared to during term-time. We applied this multiplier (and seasonality assumptions detailed below in Section 5.1.3) at each associated term start/end date (Table S11). The magnitude of the difference in $R^j(t)$ is based on the consensus value from SPI-M accounting for the increase in transmission due to the emmergence of the Alpha variant, and is consistent with the impact of school closures estimated during step two from our model fits [46]. School closure dates were taken from the average term dates agreed by councils in England, and presented in Table S11.

| Start/End Dates | Definition | Open/Closed |
|---|---|---|
| 2021-07-24 - 2021-08-31 | Summer holidays | Closed |
| 2021-09-01 - 2021-10-22 | Autumn term | Open |
| 2021-10-23 - 2021-10-31 | Half term | Closed |
| 2021-11-01 - 2021-12-17 | Autumn term | Open |
| 2021-12-18 - 2022-01-03 | Winter holidays | Closed |
| 2022-01-04 - 2022-02-11 | Spring term | Open |
| 2022-02-12 - 2022-02-20 | Half term | Closed |
| 2022-02-21 - 2022-04-01 | Spring term | Open |
| 2022-04-02 - 2022-04-18 | Easter holidays | Closed |
| 2022-04-19 - 2022-05-27 | Summer term | Open |
| 2022-05-28 - 2022-06-05 | Half term | Closed |
| 2022-06-06 - 2022-07-22 | Summer term | Open |

**Table S11:** School term dates for 2021/2022 academic year, based on the average across England.

##### 5.1.3 Seasonality

In our main analyses we assumed a slight seasonal trend in SARS-CoV-2 transmissibility throughout the year in England with 20% relative peak to trough variation. Clear evidence has yet to be demonstrated for seasonality of SARS-CoV-2 transmission, however our 20% variation assumption is considered reasonable based on theoretical modelling approaches and comparisons to pre-existing coronaviruses [47, 48]. We computed a daily multiplier for transmissibility which was:

- Maximal at 1.1 in mid-February of each year (10% relative increase compared to the mean transmissibility)
- Minimal at 0.9 on in mid-August (day 228) of each year (10% relative decrease compared to the mean transmissibility)

We applied this seasonal multiplier (Figure S8) to the value of $R^j(t)$ at each term start/end date.

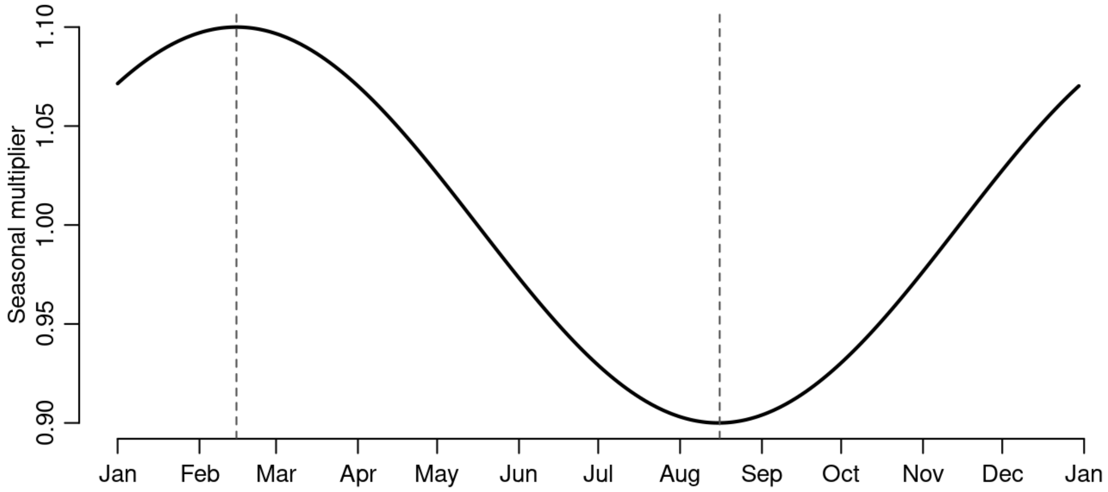

**Figure S8:** Seasonal daily multiplier for transmissibility ($R^j(t)$) applied at future change steps.

As specified above, we assumed $R^j(t)$ under each level of restrictions was distributed lognormally. For each NPI lifting scenario, we sampled from the relevant distributions of $R^j(t)$ and generated sampled trajectories of $R^j(t)$ over time by matching the ranked values obtained for each step.

##### 5.2 Vaccine roll-out

Projections of future vaccine roll-out are assumed to be 1.9 million doses per week on average [49]. For individuals aged over 50 years, we assumed that PF and AZ vaccines continue to be distributed in the proportions observed up to 31 July 2021 in each age group (Figure S4). We assumed a 60% AZ and 40% PF/Mod mix for 40-49 year olds, and assumed all individuals under 40 would receive PF or Mod. We further assume a maximum uptake for each group as summarised in Table S12. All other vaccine assumptions are the same as detailed as in Section 3.

##### 5.3 Counterfactual simulations

As well as simulating the roadmap on the planned dates, we also simulated four counterfactual projections (Table S13). We modelled:

1. The 'truth', where step four of the UK roadmap happened on 19 July 2021 and Delta was present. We assumed the range of $R^j(t)$ stated in Table S10.

| Group | Reported NHS first dose uptake data up to 19 July 2021^ | Maximum uptake in the simulation |
|---|---|---|
| Care home residents (CHR) | - | 95% |
| Care home workers (CHW) | - | 86% |
| 80+ years* | 97.6% | 95% |
| 75-79 years* | 100%+ | 99% |
| 70-74 years* | 98% | 99% |
| 65-69 years* | 95.4% | 97% |
| 60-64 years* | 99.9% | 99% |
| 55-59 years* | 97.7% | 98% |
| 50-54 years* | 92.1% | 95% |
| 45-49 years* | 86.1% | 90% |
| 40-44 years* | 89% | 90% |
| 35-39 years* | 80.7% | 80% |
| 30-34 years* | 75.9% | 80% |
| 25-29 years* | 66.2% | 80% |
| 18-24 years* | 62% | 80% |

* not care home resident/worker.

^ NIMS data, as announced 16 July 2021 [50].

+ Signifies first doses exceeds the latest estimate of the population from ONS for this group.

**Table S12:** Onward vaccine uptake assumptions by group or age.

| Name | Step four date | VOC | $R^{Alpha}(t)$ |
|---|---|---|---|
| Jul-19 | 19 July 2021 | Alpha and Delta | Table S10 |
| July Gradual | 19 July 2021 | Alpha and Delta | Gradual over 11 weeks to Table S10* |
| Jun-21 | 21 June 2021 | Alpha and Delta | Table S10 rows 4-6 |
| June Gradual | 21 June 2021 | Alpha and Delta | Gradual over 11 weeks to Table S10* |
| Alpha Only | 21 June 2021 | Alpha Only | Table S10 |

**Table S13:** Projected scenarios including the name of the scenario used in the manuscript, the assumed date of step four, whether we seed a variant of concern (Delta), and the assumed $R^j(t)$ for Alpha. *For the 'Gradual' scenario, $R^{Alpha}(t)$ gradually increases from the value on 18 July 2021 (just before full lift) to the assumed value in Table S10, in seven linear steps to 1 September 2021.

2. A 'gradual' mixing counterfactual, in which we still assume step four happened on 19 July 2021 and Delta was present, however we assume that social mixing increases gradually so that $R^j(t)$ linearly increases to the assumed Step 4 $R^j(t)$ over 11 weeks (Table S14).

3. A counterfactual in which we assume step four happened on 21 June 2021 and Delta was present. We assumed the range of $R^j(t)$ stated in Table S10.

4. A counterfactual in which we assume step four happened on 21 June 2021 and Delta was present, however we assume that social mixing increases gradually so that $R^j(t)$ linearly increases to the assumed Step 4 $R^j(t)$ over 11 weeks (Table S15).

5. A counterfactual in which we assume step four (full lift) happened on 21 June 2021 but with no seeding of Delta. In this case we do not switch to a two-variant model but instead simulate from 8 March 2021 without seeding Delta. On the date of full lift (21 June) we assume the same values as in Table S10. For simulated days between 8 March 2021 and 20 June 2021 we use the $R^{Alpha}(t)$ values estimated in the two-variant model.

| Date | $R^{Alpha}(t)$ |
|---|---|
| End Step 3 | 1.17 |
| 2021-07-19 | {1.27, 1.31, 1.36} |
| 2021-07-26 | {1.37, 1.45, 1.54} |
| 2021-08-03 | {1.46, 1.59, 1.72} |
| 2021-08-10 | {1.56, 1.73, 1.90} |
| 2021-08-18 | {1.65, 1.87, 2.09} |
| 2021-08-25 | {1.75, 2.01, 2.27} |
| 2021-09-01 | {1.84, 2.15, 2.45} |
| 2021-09-09 | {1.94, 2.29, 2.63} |
| 2021-09-16 | {2.04, 2.43, 2.82} |
| 2021-09-24 | {2.13, 2.56, 3.00} |
| 2021-10-01 | {2.23, 2.70, 3.18} |

**Table S14:** Gradual July scenario $R^{Alpha}(t)$ schedule under central assumptions. The value of $R^{Alpha}(t)$ estimated at 2021-07-18 linearly increases over 11 weeks to the assumed Step 4 $R^{Alpha}(t)$ value (Table S10). The three $R^{Alpha}(t)$ values correspond to optimistic/central/pessimistic mixing assumptions.

| Date | $R^{Alpha}(t)$ |
|---|---|
| End Step 3 | 1.56 |
| 2021-06-21 | {1.62, 1.66, 1.71} |
| 2021-06-28 | {1.68, 1.77, 1.85} |
| 2021-07-05 | {1.74, 1.87, 2.00} |
| 2021-07-13 | {1.80, 1.98, 2.15} |
| 2021-07-20 | {1.86, 2.08, 2.30} |
| 2021-07-27 | {1.92, 2.18, 2.44} |
| 2021-08-03 | {1.98, 2.29, 2.59} |
| 2021-08-10 | {2.04, 2.39, 2.74} |
| 2021-08-18 | {2.11, 2.50, 2.89} |
| 2021-08-25 | {2.17, 2.60, 3.03} |
| 2021-09-01 | {2.23, 2.70, 3.18} |

**Table S15:** Gradual June scenario $R^{Alpha}(t)$ schedule under central assumptions. The value of $R^{Alpha}(t)$ estimated at 2021-06-20 linearly increases over 11 weeks to the assumed Step 4 $R^{Alpha}(t)$ value (Table S10). The three $R^{Alpha}(t)$ values correspond to optimistic/central/pessimistic mixing assumptions.

##### 5.4 Sensitivity analysis

Finally as a sensitivity analysis we project forward the two July scenarios (Table S13, rows 1-2) under different combinations of:

1. Vaccine effectiveness vs Delta - Table S3.
2. Cross immunity vs Delta - $\{1, 0.85, 0.75\}$ (Table S7).
3. Waning immunity - $\{$None, 6 years, 3 years$\}$ (Table S7).
4. Contact mixing - $\{$Optimistic, central, pessimistic$\}$ (Table S10).

#### 6 Software and Implementation

Implementation of the model described above is fully described in FitzJohn et al. [51]. The primary interface to the model is coded in R [52] with functions written in packages `sircovid` and `spimalot`. The model is written in `odin` and run with `dust`, the pMCMC functions are written in `mcstate`. For this paper we used `sircovid` v0.11.30, `spimalot` v0.2.52, `dust` v0.9.11, and `mcstate` v0.6.5. The above packages are publicly available in the *mrc-ide* GitHub organisation (https://github.com/mrc-ide/). The code and scripts used to create the results in this paper are available in https://github.com/mrc-ide/sarscov2-roadmap-england.

### 7 Supplementary Results

#### 7.1 Model fit to data

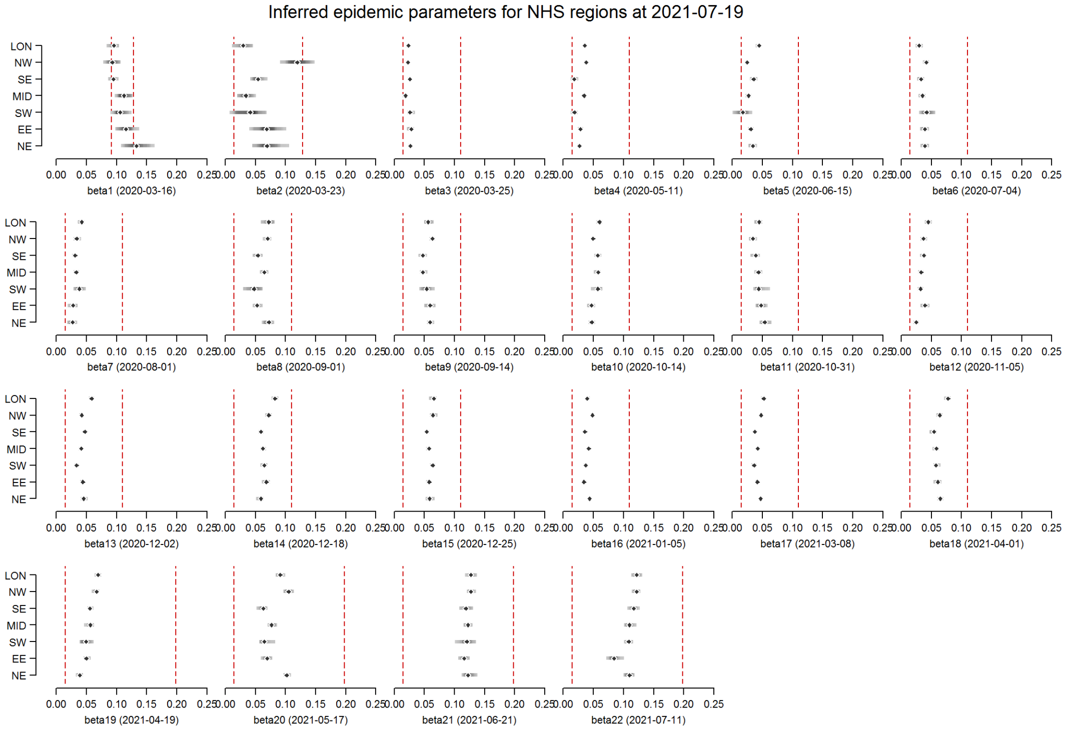

**Figure S9:** Posterior distributions for $\beta_i$ for initial variant-agnostic model fits. Red bars indicate prior distribution range, black bars indicate the 95% CrI. As explained in Section 2.1, the parameters corresponding to after the two-variant model is introduced ($\beta_{17}$ - $\beta_{22}$) are later re-calculated after model restarts, using the posterior distributions presented here as prior distributions. Final posterior distributions for $\beta_{17}$ - $\beta_{24}$ are presented below in Figure S10.

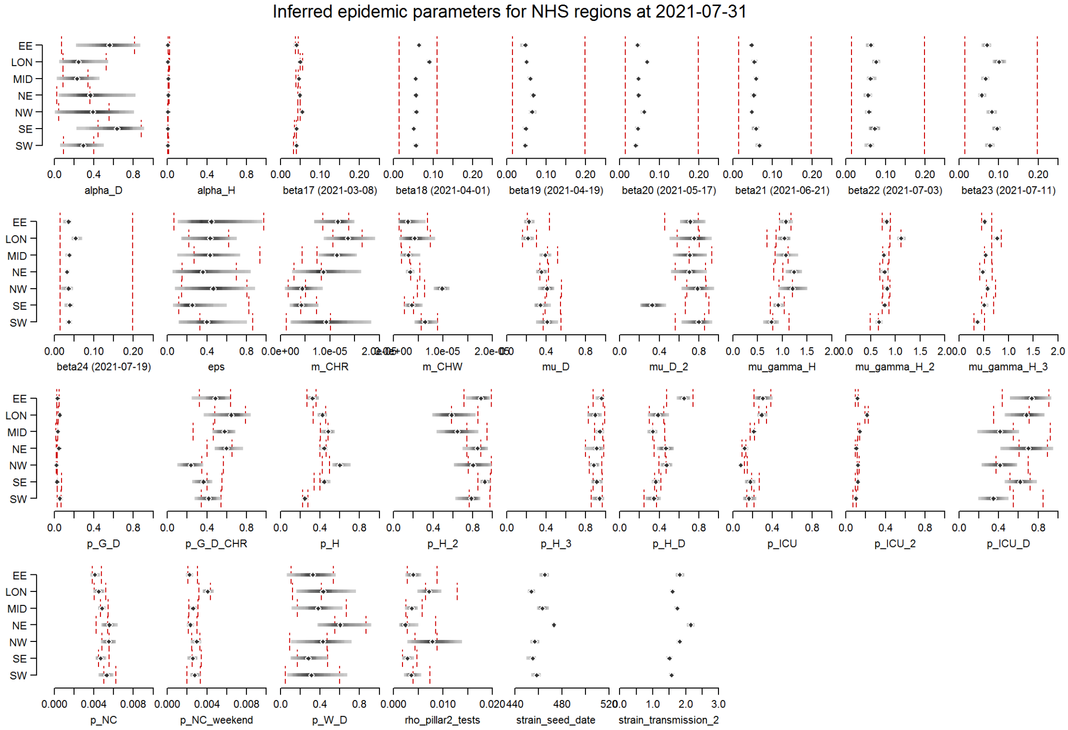

**Figure S10:** Final model parameter posterior distributions. Red bars indicate prior distribution range, black bars indicate the 95% CrI.

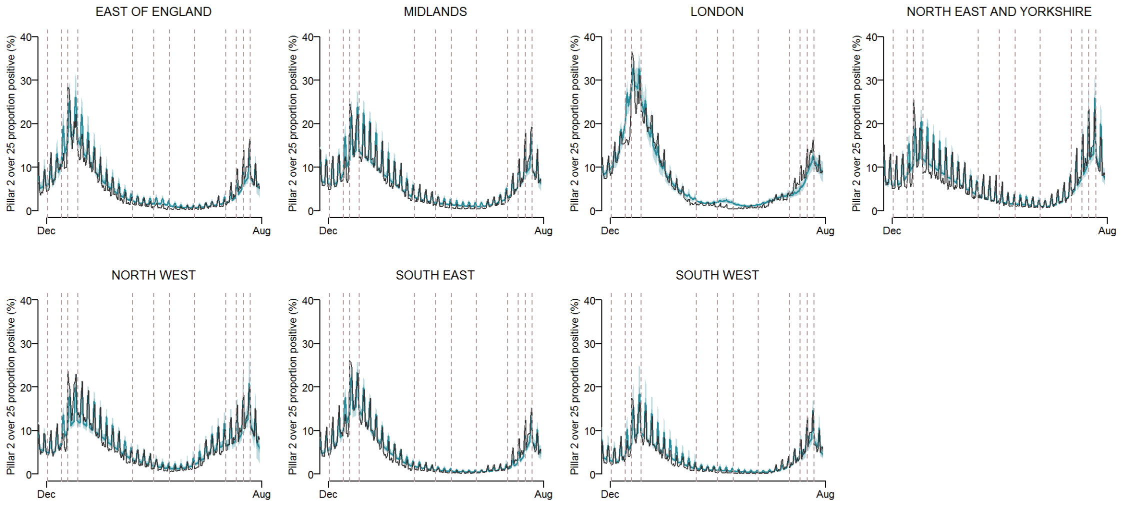

**Figure S11:** Model fits by region to PCR positivity from Pillar 2 for individuals aged > 25 years. The solid blue line shows the median model fit and the shaded area the 95% CrI. The black line depicts the recorded data.

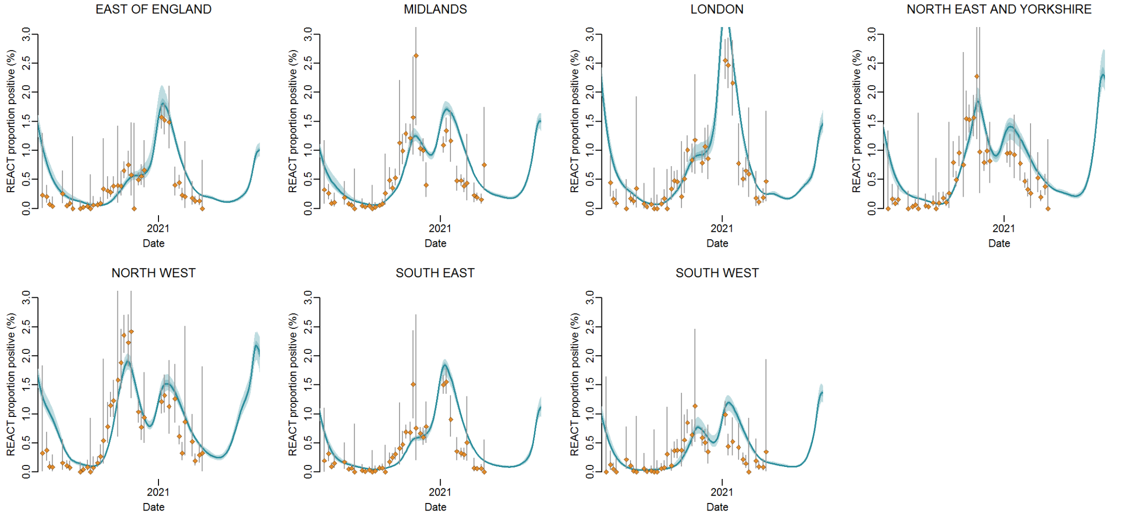

**Figure S12:** Model fits by region to PCR positivity from the REACT-1 study . The points show the data and bars the 95% CI. The solid blue line shows the median model fit and the shaded area the 95% CrI.

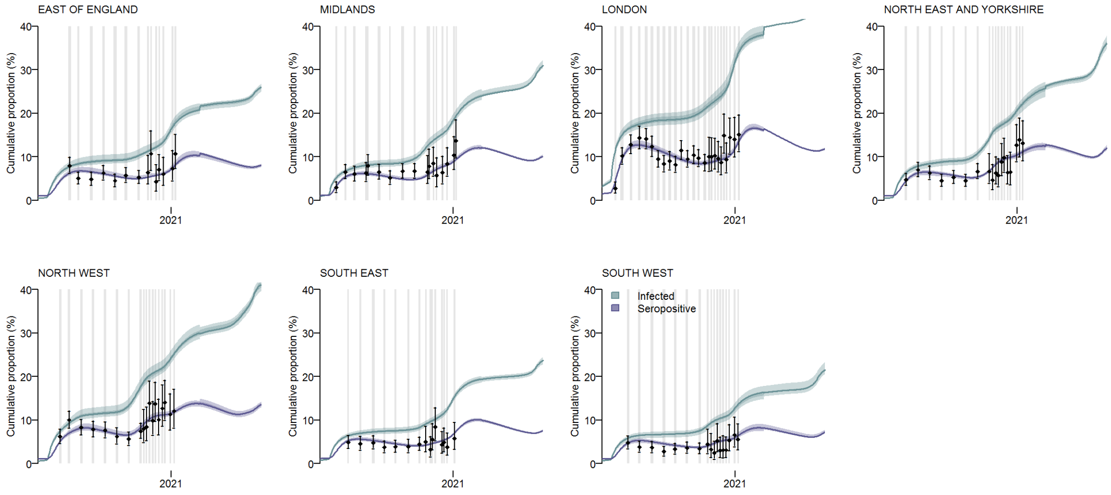

**Figure S13:** Model fits by region to EuroImmun serology assay data (Table S1). The points show the data and bars the 95% CI. The solid purple line shows the median model fit, the solid blue line shows the median cumulative infections, the shaded areas the respective 95% CrI.

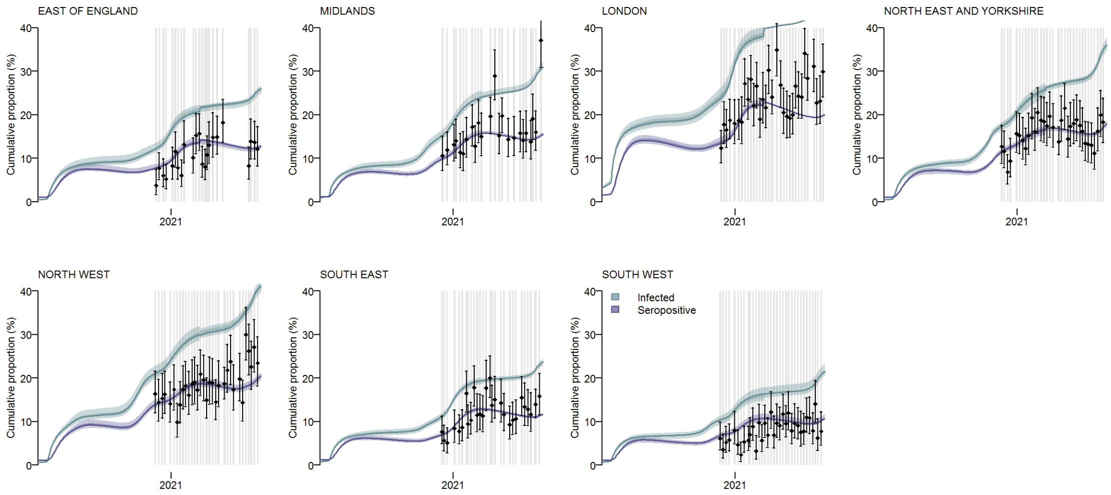

**Figure S14:** Model fits by region to Roche serology assay data (Table S1). The points show the data and bars the 95% CI. The solid purple line shows the median model fit, the solid blue line shows the median cumulative infections, the shaded areas the respective 95% CrI.

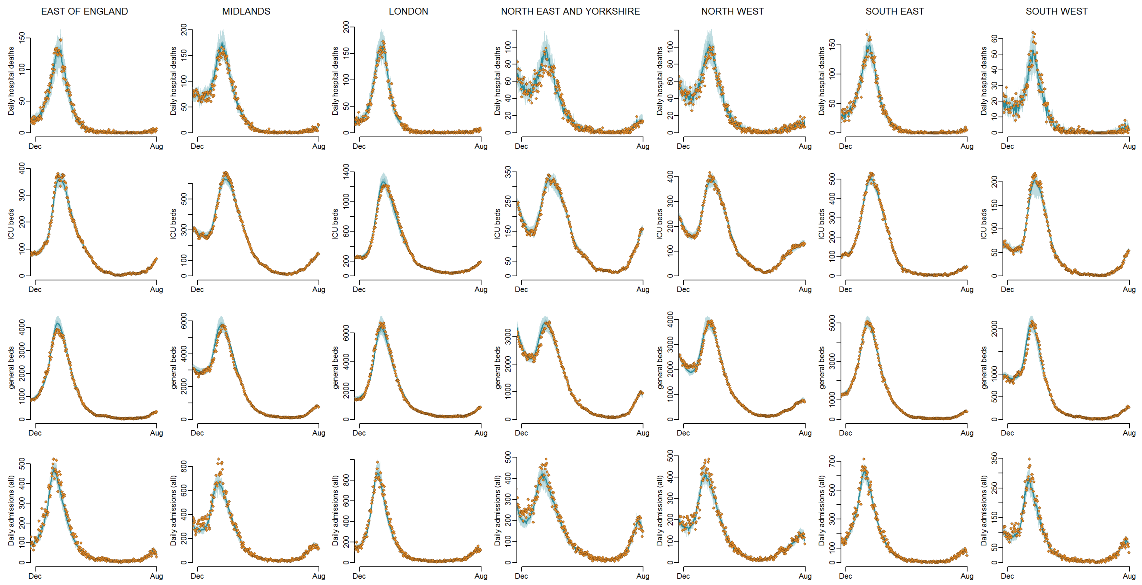

**Figure S15:** Model fits to England NHS regions: daily hospital deaths (top row), all hospital beds occupancy (second row), and all daily admissions (bottom row) by England NHS region (columns). The points show the data, the solid line the median model fit and the shaded area the 95% CrI. Green points indicate data streams that the model does not fit to.

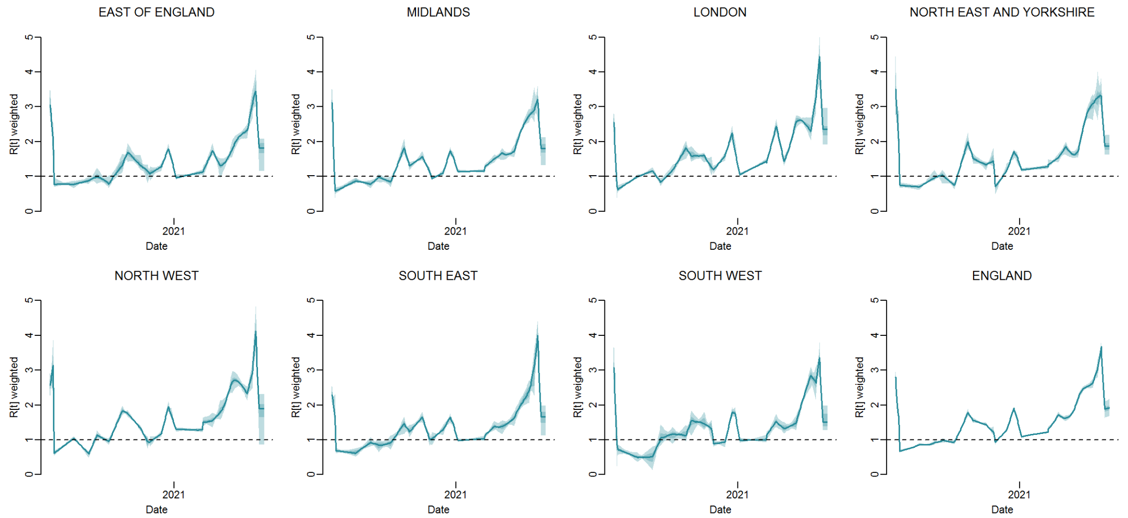

**Figure S16:** Inferred value of $R^j(t)$ by region over time. The solid line shows the median model fit and the shaded area shows the 95% CrI.

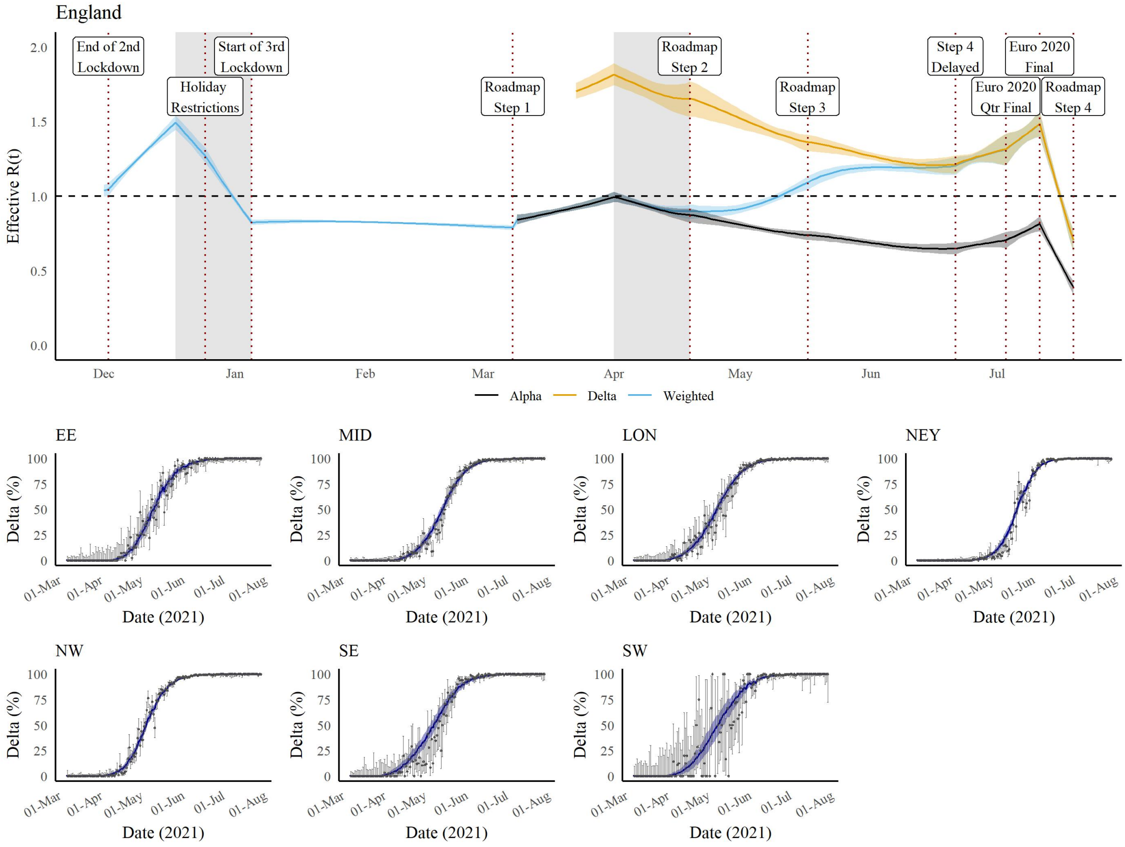

**Figure S17:** Trajectory of the COVID-19 epidemic in England and the emergence of the Delta variant under central assumptions. (A) Prevalence weighted (blue) and variant-specific (Alpha = black, Delta = orange) effective reproduction number over time from the end of the second national lockdown to 19 July 2021. The solid line shows the median and the shaded area the 95% CrI. The vertical dashed lines show key dates of the roadmap steps and the shaded area the school holidays. (B-H) Model fit to the proportion of Delta cases (variant and mutation data, VAM) over time from 8 March to 19 July. The points show the data, the bar the 95% CI, the blue line the model fit, and the shaded area the 95% CrI. LON = London; SE = South East; NW = North West; SW = South West; MID = Midlands; EE = East of England; NEY = North East and Yorkshire.

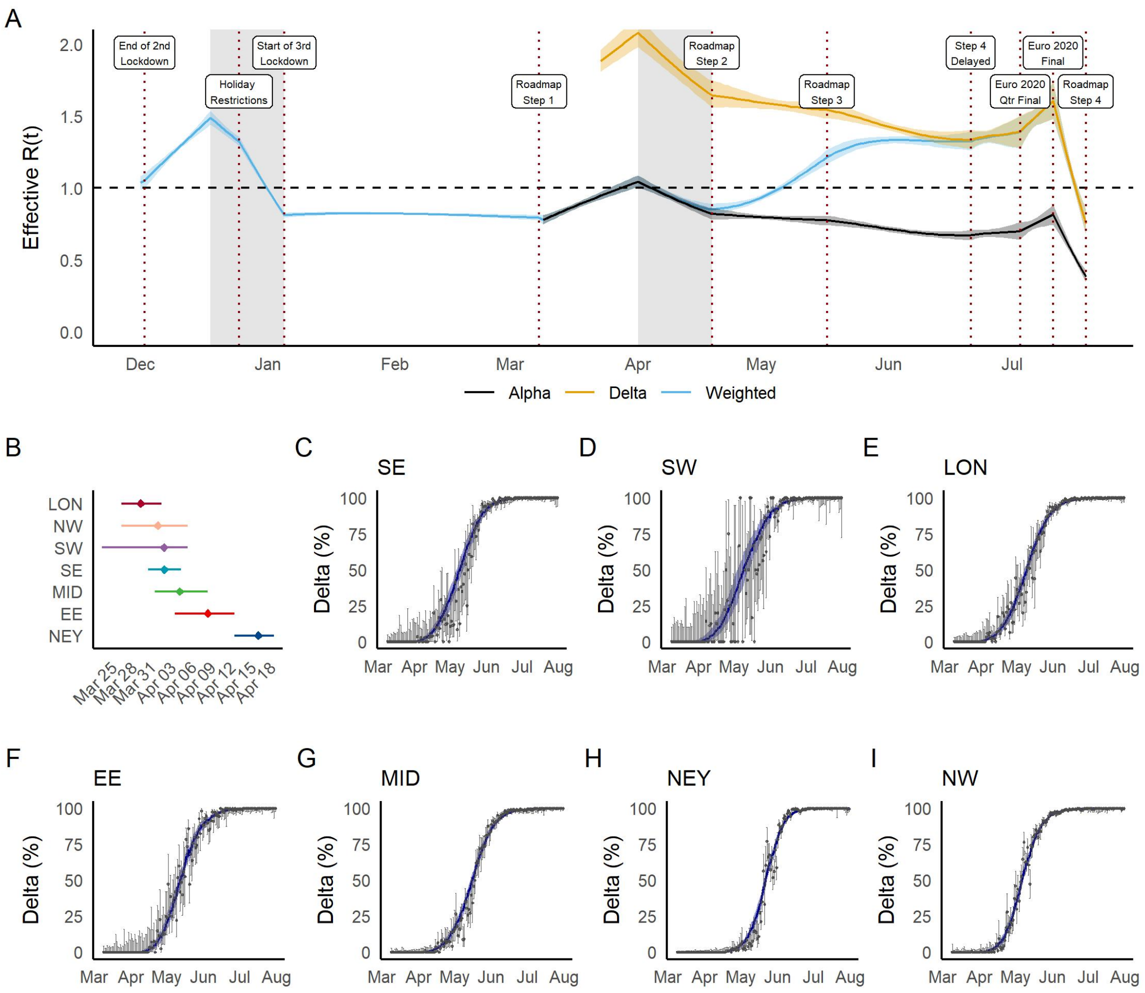

**Figure S18:** Trajectory of the COVID-19 epidemic in England and the emergence of the Delta variant under optimistic assumptions. (A) Prevalence weighted (blue) and variant-specific (Alpha = black, Delta = orange) effective reproduction number over time from the end of the second national lockdown to 19 July 2021. The solid line shows the median and the shaded area the 95% CrI. The vertical dashed lines show key dates of the roadmap steps and the shaded area the school holidays. (B) Estimated Delta seeding date by NHS region. (C-I) Model fit to the proportion of Delta cases (variant and mutation data, VAM) over time from 8 March to 19 July. The points show the data, the bar the 95% CI, the blue line the model fit, and the shaded area the 95% CrI. LON = London; SE = South East; NW = North West; SW = South West; MID = Midlands; EE = East of England; NEY = North East and Yorkshire.

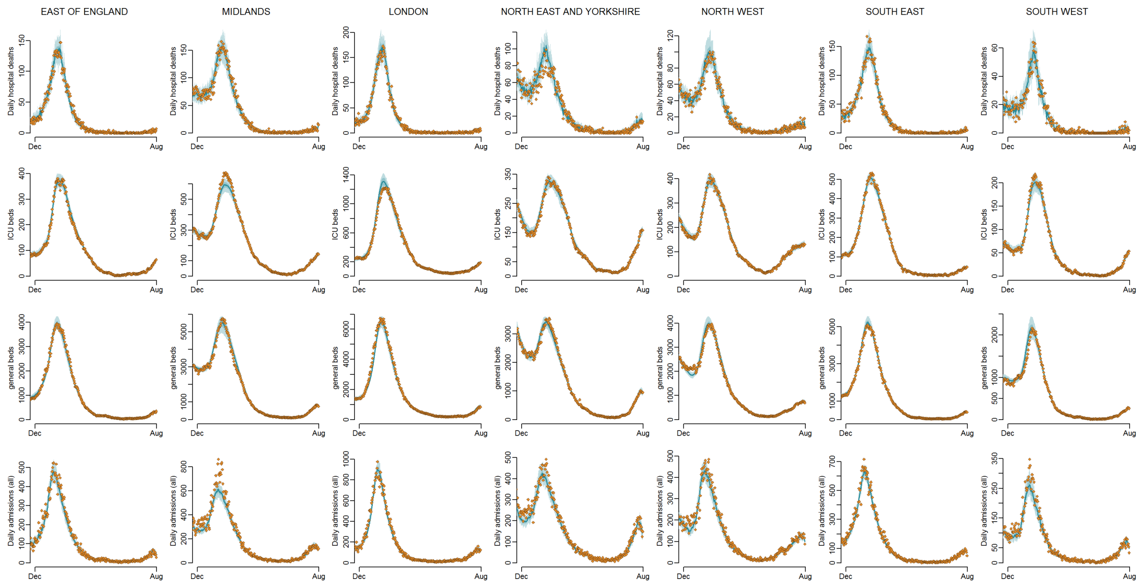

**Figure S19:** Model fits to England NHS regions: daily hospital deaths (top row), all hospital beds occupancy (second row), and all daily admissions (bottom row) by England NHS region (columns) for the optimistic assumptions. The points show the data, the solid line the median model fit and the shaded area the 95% CrI. Green points indicate data streams that the model does not fit to.

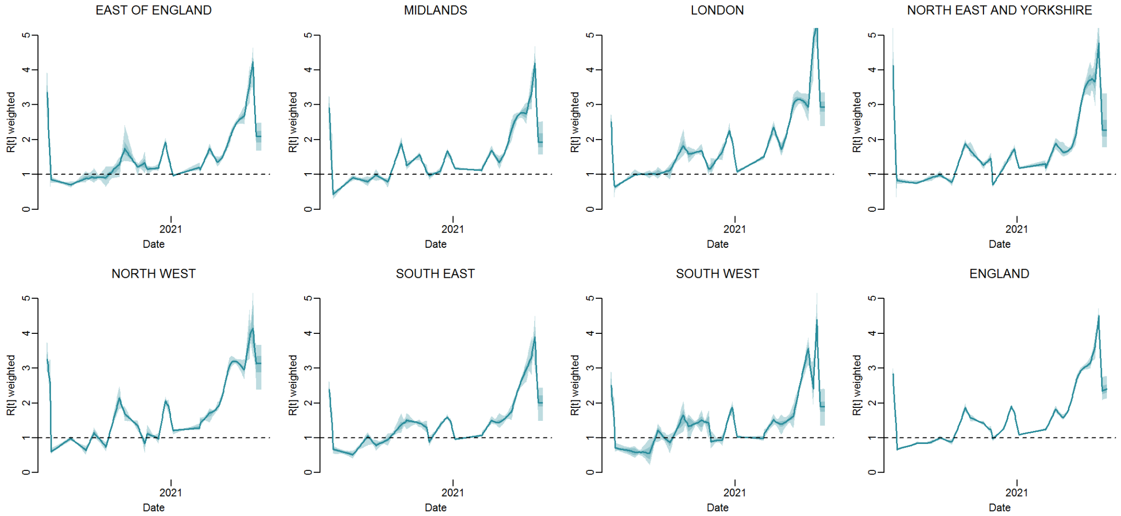

**Figure S20:** Inferred value of $R^j(t)$ by region over time for the optimistic assumptions. The solid line shows the median model fit and the shaded area shows the 95% CrI.

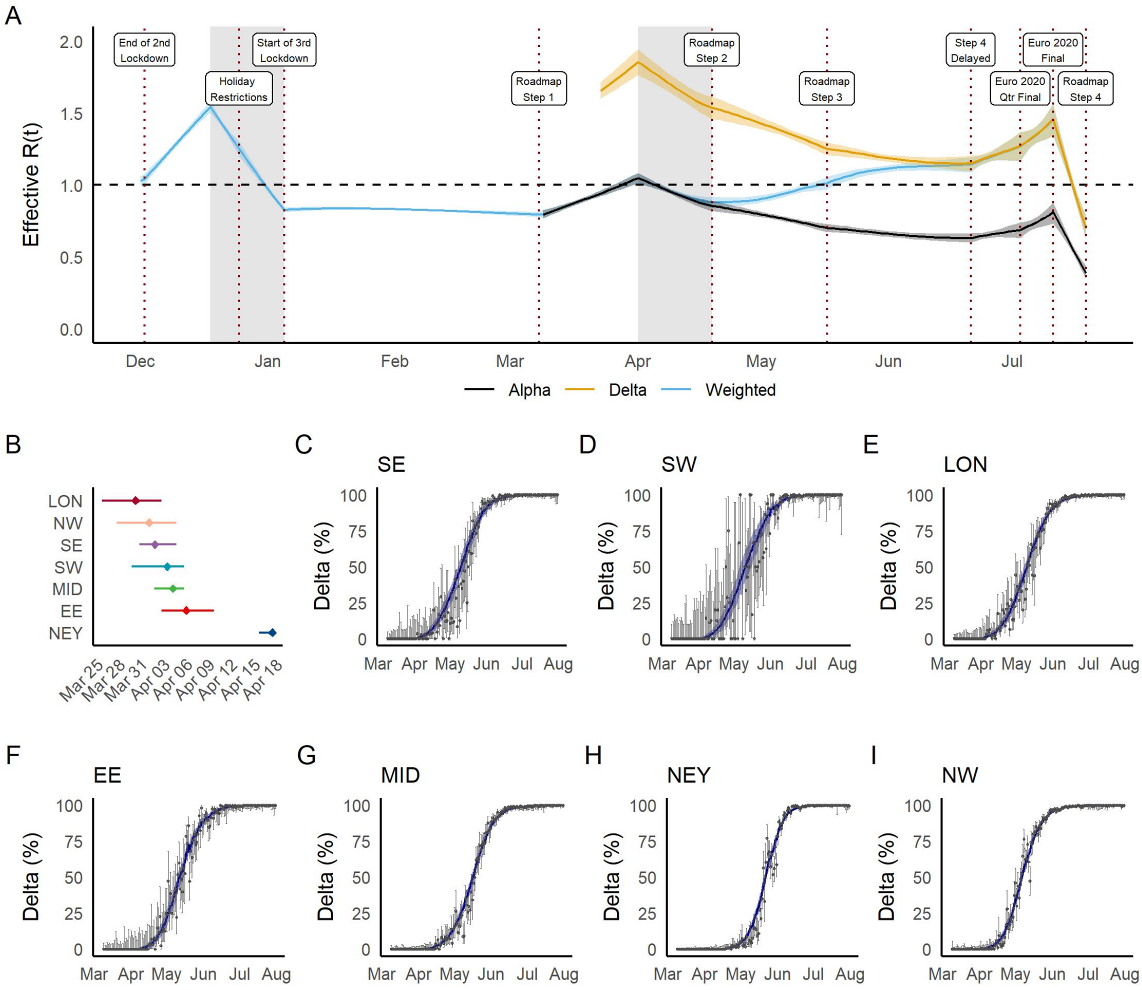

**Figure S21:** Trajectory of the COVID-19 epidemic in England and the emergence of the Delta variant under pessimistic assumptions. (A) Prevalence weighted (blue) and variant-specific (Alpha = black, Delta = orange) effective reproduction number over time from the end of the second national lockdown to 19 July 2021. The solid line shows the median and the shaded area the 95% CrI. The vertical dashed lines show key dates of the roadmap steps and the shaded area the school holidays. (B) Estimated Delta seeding date by NHS region. (C-I) Model fit to the proportion of Delta cases (variant and mutation data, VAM) over time from 8 March to 19 July. The points show the data, the bar the 95% CI, the blue line the model fit, and the shaded area the 95% CrI. LON = London; SE = South East; NW = North West; SW = South West; MID = Midlands; EE = East of England; NEY = North East and Yorkshire.

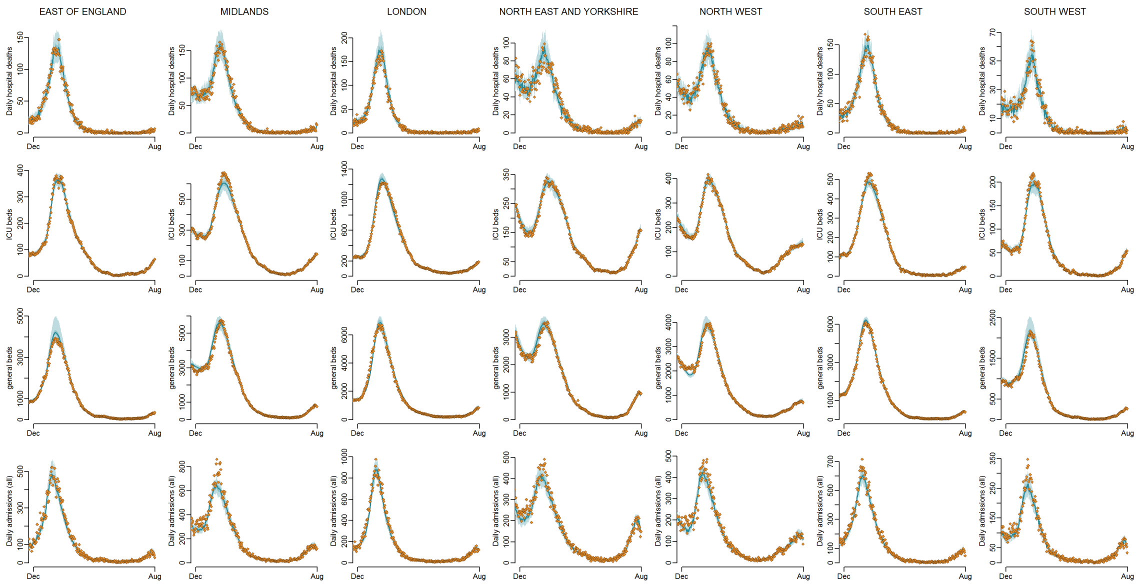

**Figure S22:** Model fits to England NHS regions: daily hospital deaths (top row), all hospital beds occupancy (second row), and all daily admissions (bottom row) by England NHS region (columns) for the pessimistic assumptions. The points show the data, the solid line the median model fit and the shaded area the 95% CrI. Green points indicate data streams that the model does not fit to.

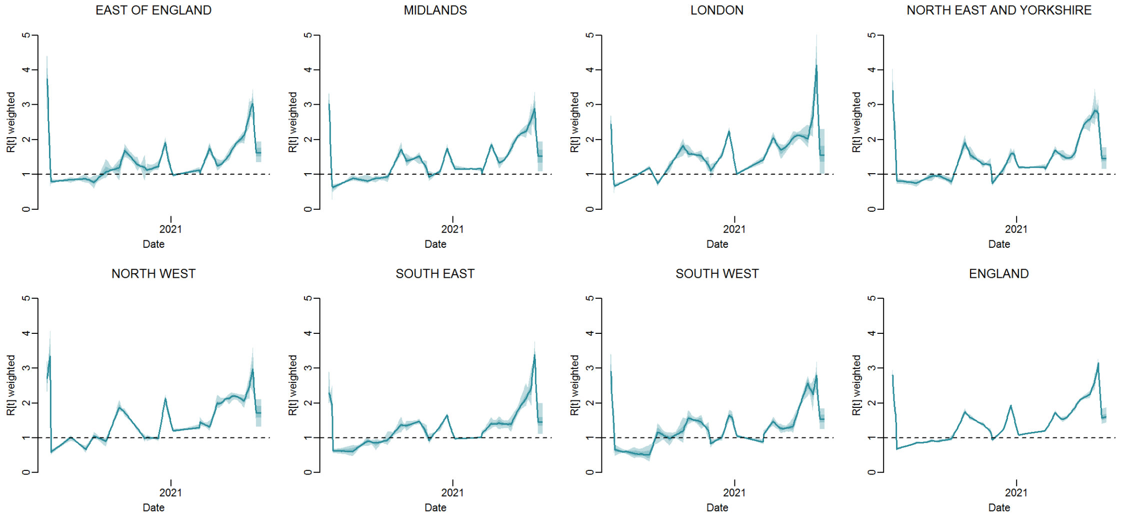

**Figure S23:** Inferred value of $R^j(t)$ by region over time for the pessimistic assumptions. The solid line shows the median model fit and the shaded area shows the 95% CrI.

#### 7.2 Simulation results

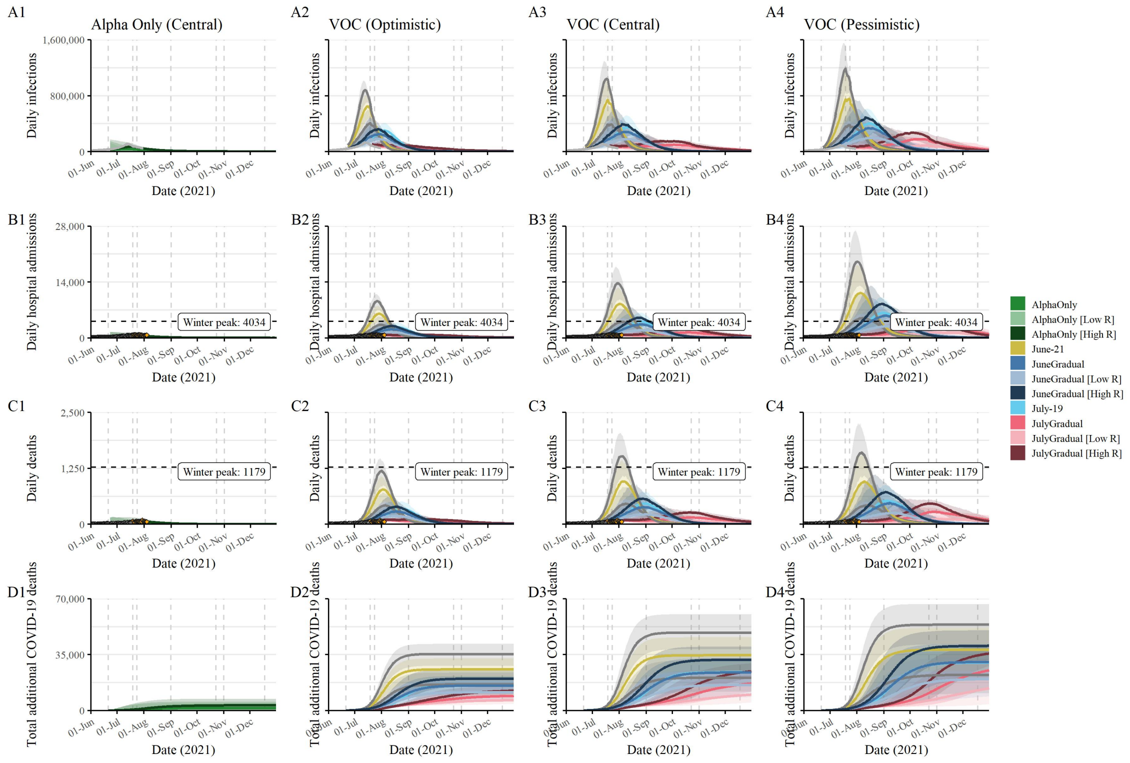

**Figure S24:** Forward projections for states under different assumptions and scenarios. (A1-D1) counterfactual without seeding of Delta. (A2-D2) scenarios with Delta under optimistic immunity assumptions. (A3-D3) scenarios with Delta under central immunity assumptions. (A4-D4) scenarios with Delta under pessimistic immunity assumptions. (A1-A4) projected daily infections. (B1-B4) projected daily hospital admissions. (C1-C4) projected daily deaths. (D1-D4) projected total COVID-19 deaths up to 11 months after full lift.

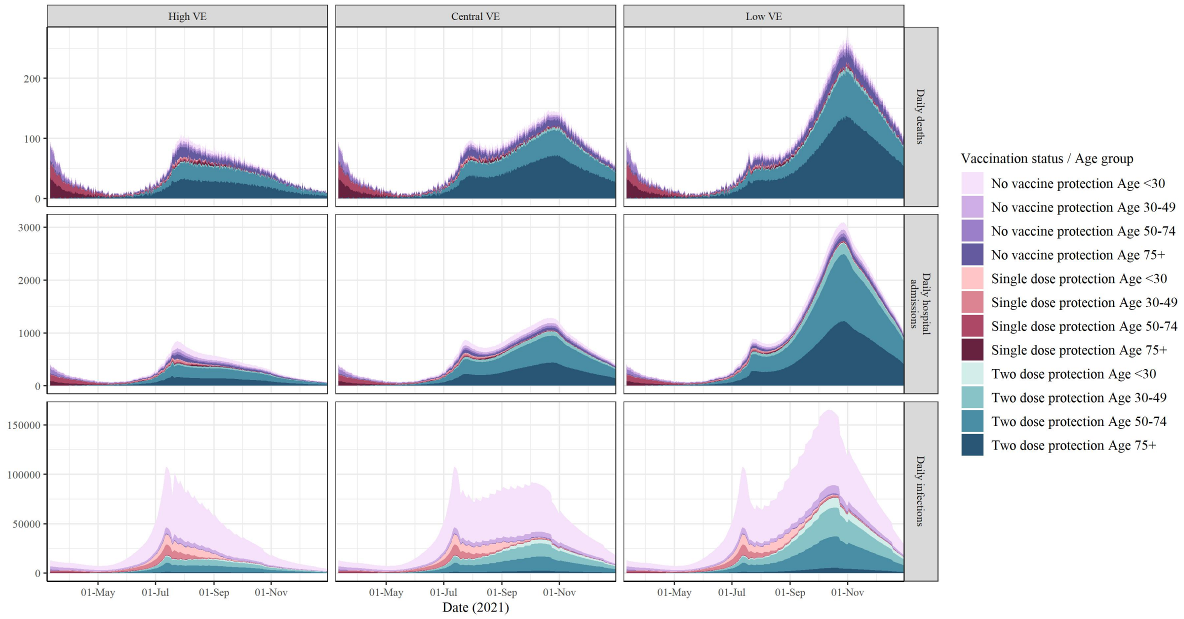

**Figure S25:** Simulations by age and vaccination status in the 'July-19' scenario. COVID-19 daily deaths (top), hospital admissions (middle), infections (bottom) by age group and vaccination status with optimistic (left), central (middle), and pessimistic (right) assumptions.

**Table S16:** Peak hospital admissions and deaths, and cumulative infections and deaths (with 95% confidence intervals) in forward projections across all analyses (column 1), scenarios (column 2), and states (column 3).

| Analysis | Scenario | State | Value |
|---|---|---|---|
| AlphaOnly | VOC Central VE | Daily deaths | 0 (0, 100) |
| AlphaOnly | VOC Central VE | Daily hospital admissions | 300 (0, 600) |
| AlphaOnly | VOC Central VE | Daily hospital deaths | 0 (0, 100) |
| AlphaOnly | VOC Central VE | Daily infections | 36800 (700, 77200) |
| AlphaOnly | VOC Central VE | Total additional COVID-19 deaths | 1700 (0, 5500) |
| AlphaOnly | VOC Central VE | Total additional COVID-19 infections | 2001700 (74400, 3632700) |
| AlphaOnly | VOC High VE | Daily deaths | 100 (0, 200) |
| AlphaOnly | VOC High VE | Daily hospital admissions | 600 (0, 1300) |
| AlphaOnly | VOC High VE | Daily hospital deaths | 0 (0, 100) |
| AlphaOnly | VOC High VE | Daily infections | 77900 (2300, 147700) |
| AlphaOnly | VOC High VE | Total additional COVID-19 deaths | 3600 (300, 7400) |
| AlphaOnly | VOC High VE | Total additional COVID-19 infections | 3210400 (500900, 4403400) |
| AlphaOnly | VOC Low VE | Daily deaths | 0 (0, 100) |
| AlphaOnly | VOC Low VE | Daily hospital admissions | 100 (0, 500) |
| AlphaOnly | VOC Low VE | Daily hospital deaths | 0 (0, 0) |
| AlphaOnly | VOC Low VE | Daily hospital deaths | 0 (0, 0) |
| AlphaOnly | VOC Low VE | Daily infections | 15700 (200, 59400) |
| AlphaOnly | VOC Low VE | Total additional COVID-19 deaths | 900 (0, 4400) |
| AlphaOnly | VOC Low VE | Total additional COVID-19 infections | 1188400 (11000, 3177200) |
| AlphaOnly [High R] | VOC Central VE | Daily deaths | 100 (0, 100) |
| AlphaOnly [High R] | VOC Central VE | Daily hospital admissions | 500 (0, 1000) |
| AlphaOnly [High R] | VOC Central VE | Daily hospital deaths | 0 (0, 100) |
| AlphaOnly [High R] | VOC Central VE | Daily infections | 71900 (1600, 119600) |
| AlphaOnly [High R] | VOC Central VE | Total additional COVID-19 deaths | 3600 (1700, 7300) |
| AlphaOnly [High R] | VOC Central VE | Total additional COVID-19 infections | 4210800 (2323200, 5608100) |
| AlphaOnly [High R] | VOC High VE | Daily deaths | 100 (0, 200) |
| AlphaOnly [High R] | VOC High VE | Daily hospital admissions | 1000 (0, 1600) |
| AlphaOnly [High R] | VOC High VE | Daily hospital deaths | 100 (0, 100) |
| AlphaOnly [High R] | VOC High VE | Daily infections | 116500 (5200, 178700) |

Continued on next page

| Table S16 – continued from previous page | | | |
|---|---|---|---|
| Analysis | Scenario | State | Value |
| AlphaOnly [High R] | VOC High VE | Total additional COVID-19 deaths | 5900 (2000, 9200) |
| AlphaOnly [High R] | VOC High VE | Total additional COVID-19 infections | 5253800 (2757000, 6371000) |
| AlphaOnly [High R] | VOC Low VE | Daily deaths | 0 (0, 100) |
| AlphaOnly [High R] | VOC Low VE | Daily hospital admissions | 200 (0, 600) |
| AlphaOnly [High R] | VOC Low VE | Daily hospital deaths | 0 (0, 100) |
| AlphaOnly [High R] | VOC Low VE | Daily infections | 32800 (300, 104900) |
| AlphaOnly [High R] | VOC Low VE | Total additional COVID-19 deaths | 2900 (600, 5800) |
| AlphaOnly [High R] | VOC Low VE | Total additional COVID-19 infections | 3922600 (1101700, 5077900) |
| AlphaOnly [Low R] | VOC Central VE | Daily deaths | 0 (0, 100) |
| AlphaOnly [Low R] | VOC Central VE | Daily hospital admissions | 100 (0, 800) |
| AlphaOnly [Low R] | VOC Central VE | Daily hospital deaths | 0 (0, 100) |
| AlphaOnly [Low R] | VOC Central VE | Daily infections | 21300 (400, 107700) |
| AlphaOnly [Low R] | VOC Central VE | Total additional COVID-19 deaths | 800 (0, 4500) |
| AlphaOnly [Low R] | VOC Central VE | Total additional COVID-19 infections | 852000 (15500, 2327800) |
| AlphaOnly [Low R] | VOC High VE | Daily deaths | 0 (0, 200) |
| AlphaOnly [Low R] | VOC High VE | Daily hospital admissions | 400 (0, 1600) |
| AlphaOnly [Low R] | VOC High VE | Daily hospital deaths | 0 (0, 100) |
| AlphaOnly [Low R] | VOC High VE | Daily infections | 57800 (1500, 153600) |
| AlphaOnly [Low R] | VOC High VE | Total additional COVID-19 deaths | 2100 (100, 6200) |
| AlphaOnly [Low R] | VOC High VE | Total additional COVID-19 infections | 1783400 (62300, 2882700) |
| AlphaOnly [Low R] | VOC Low VE | Daily deaths | 0 (0, 100) |
| AlphaOnly [Low R] | VOC Low VE | Daily hospital admissions | 100 (0, 700) |
| AlphaOnly [Low R] | VOC Low VE | Daily hospital deaths | 0 (0, 100) |
| AlphaOnly [Low R] | VOC Low VE | Daily infections | 8100 (100, 90500) |
| AlphaOnly [Low R] | VOC Low VE | Total additional COVID-19 deaths | 300 (0, 3500) |
| AlphaOnly [Low R] | VOC Low VE | Total additional COVID-19 infections | 327300 (2800, 1943700) |
| July-19 | VOC Central VE | Daily deaths | 500 (300, 700) |
| July-19 | VOC Central VE | Daily hospital admissions | 4100 (2400, 6300) |
| July-19 | VOC Central VE | Daily hospital deaths | 300 (200, 500) |
| July-19 | VOC Central VE | Daily infections | 348900 (221800, 514700) |
| July-19 | VOC Central VE | Total additional COVID-19 deaths | 25000 (18200, 33100) |

Continued on next page

**Table S16 – continued from previous page**

| Analysis | Scenario | State | Value |
|---|---|---|---|
| July-19 | VOC Central VE | Total additional COVID-19 infections | 14371700 (10802400, 18079900) |
| July-19 | VOC High VE | Daily deaths | 400 (200, 500) |
| July-19 | VOC High VE | Daily hospital admissions | 2700 (1900, 3800) |
| July-19 | VOC High VE | Daily hospital deaths | 200 (100, 300) |
| July-19 | VOC High VE | Daily infections | 302000 (221700, 412700) |
| July-19 | VOC High VE | Total additional COVID-19 deaths | 17000 (13000, 21600) |
| July-19 | VOC High VE | Total additional COVID-19 infections | 10109300 (8159900, 12385600) |
| July-19 | VOC Low VE | Daily deaths | 500 (300, 900) |
| July-19 | VOC Low VE | Daily hospital admissions | 6300 (3100, 10000) |
| July-19 | VOC Low VE | Daily hospital deaths | 300 (200, 600) |
| July-19 | VOC Low VE | Daily infections | 402100 (218800, 603100) |
| July-19 | VOC Low VE | Total additional COVID-19 deaths | 31100 (21100, 41300) |
| July-19 | VOC Low VE | Total additional COVID-19 infections | 18984600 (13947300, 23674200) |
| July-19 [High R] | VOC Central VE | Daily deaths | 800 (500, 1100) |
| July-19 [High R] | VOC Central VE | Daily hospital admissions | 7300 (5000, 9500) |
| July-19 [High R] | VOC Central VE | Daily hospital deaths | 500 (400, 700) |
| July-19 [High R] | VOC Central VE | Daily infections | 552700 (408300, 716000) |
| July-19 [High R] | VOC Central VE | Total additional COVID-19 deaths | 35800 (28300, 43800) |
| July-19 [High R] | VOC Central VE | Total additional COVID-19 infections | 18796000 (15725500, 21839900) |
| July-19 [High R] | VOC High VE | Daily deaths | 600 (400, 700) |
| July-19 [High R] | VOC High VE | Daily hospital admissions | 4300 (3300, 5500) |
| July-19 [High R] | VOC High VE | Daily hospital deaths | 400 (300, 500) |
| July-19 [High R] | VOC High VE | Daily infections | 437800 (349600, 550800) |
| July-19 [High R] | VOC High VE | Total additional COVID-19 deaths | 23200 (18900, 28000) |
| July-19 [High R] | VOC High VE | Total additional COVID-19 infections | 12849300 (11127800, 14854500) |
| July-19 [High R] | VOC Low VE | Daily deaths | 900 (500, 1300) |
| July-19 [High R] | VOC Low VE | Daily hospital admissions | 11300 (6900, 15300) |
| July-19 [High R] | VOC Low VE | Daily hospital deaths | 600 (400, 800) |
| July-19 [High R] | VOC Low VE | Daily infections | 659200 (427100, 864800) |
| July-19 [High R] | VOC Low VE | Total additional COVID-19 deaths | 43100 (32900, 52000) |
| July-19 [High R] | VOC Low VE | Total additional COVID-19 infections | 24334600 (20029300, 28025400) |

Continued on next page

**Table S16 – continued from previous page**

| Analysis | Scenario | State | Value |
|---|---|---|---|
| July-19 [Low R] | VOC Central VE | Daily deaths | 200 (100, 400) |
| July-19 [Low R] | VOC Central VE | Daily hospital admissions | 2000 (1000, 3500) |
| July-19 [Low R] | VOC Central VE | Daily hospital deaths | 100 (100, 300) |
| July-19 [Low R] | VOC Central VE | Daily infections | 187100 (102600, 319900) |
| July-19 [Low R] | VOC Central VE | Total additional COVID-19 deaths | 15700 (10100, 23100) |
| July-19 [Low R] | VOC Central VE | Total additional COVID-19 infections | 9650700 (6028000, 13780900) |
| July-19 [Low R] | VOC High VE | Daily deaths | 200 (100, 300) |
| July-19 [Low R] | VOC High VE | Daily hospital admissions | 1500 (1000, 2300) |
| July-19 [Low R] | VOC High VE | Daily hospital deaths | 100 (100, 200) |
| July-19 [Low R] | VOC High VE | Daily infections | 183600 (124400, 274100) |
| July-19 [Low R] | VOC High VE | Total additional COVID-19 deaths | 11300 (8000, 15600) |
| July-19 [Low R] | VOC High VE | Total additional COVID-19 infections | 7008700 (4997900, 9617500) |
| July-19 [Low R] | VOC Low VE | Daily deaths | 200 (100, 500) |
| July-19 [Low R] | VOC Low VE | Daily hospital admissions | 2600 (900, 5700) |
| July-19 [Low R] | VOC Low VE | Daily hospital deaths | 100 (0, 300) |
| July-19 [Low R] | VOC Low VE | Daily infections | 190600 (76700, 382200) |
| July-19 [Low R] | VOC Low VE | Total additional COVID-19 deaths | 19600 (10800, 29700) |
| July-19 [Low R] | VOC Low VE | Total additional COVID-19 infections | 12814400 (7459100, 18578800) |
| JulyGradual | VOC Central VE | Daily deaths | 200 (100, 200) |
| JulyGradual | VOC Central VE | Daily hospital admissions | 1400 (700, 1500) |
| JulyGradual | VOC Central VE | Daily hospital deaths | 100 (0, 100) |
| JulyGradual | VOC Central VE | Daily infections | 98900 (48400, 107600) |
| JulyGradual | VOC Central VE | Total additional COVID-19 deaths | 20400 (17100, 24600) |
| JulyGradual | VOC Central VE | Total additional COVID-19 infections | 11795000 (10167600, 13964200) |
| JulyGradual | VOC High VE | Daily deaths | 100 (100, 100) |
| JulyGradual | VOC High VE | Daily hospital admissions | 800 (800, 900) |
| JulyGradual | VOC High VE | Daily hospital deaths | 100 (0, 100) |
| JulyGradual | VOC High VE | Daily infections | 102400 (76100, 129700) |
| JulyGradual | VOC High VE | Total additional COVID-19 deaths | 10700 (8800, 13400) |
| JulyGradual | VOC High VE | Total additional COVID-19 infections | 6445600 (5242500, 8245400) |
| JulyGradual | VOC Low VE | Daily deaths | 300 (100, 400) |

Continued on next page

**Table S16 – continued from previous page**

| Analysis | Scenario | State | Value |
|---|---|---|---|
| JulyGradual | VOC Low VE | Daily hospital admissions | 3400 (1100, 3800) |
| JulyGradual | VOC Low VE | Daily hospital deaths | 200 (100, 200) |
| JulyGradual | VOC Low VE | Daily infections | 177900 (62500, 200800) |
| JulyGradual | VOC Low VE | Total additional COVID-19 deaths | 29500 (23400, 35100) |
| JulyGradual | VOC Low VE | Total additional COVID-19 infections | 17653400 (14832000, 20480200) |
| JulyGradual [High R] | VOC Central VE | Daily deaths | 300 (200, 300) |
| JulyGradual [High R] | VOC Central VE | Daily hospital admissions | 2300 (1700, 2500) |
| JulyGradual [High R] | VOC Central VE | Daily hospital deaths | 200 (100, 200) |
| JulyGradual [High R] | VOC Central VE | Daily infections | 151800 (74700, 178500) |
| JulyGradual [High R] | VOC Central VE | Total additional COVID-19 deaths | 27300 (23600, 31300) |
| JulyGradual [High R] | VOC Central VE | Total additional COVID-19 infections | 14674000 (13237000, 16435900) |
| JulyGradual [High R] | VOC High VE | Daily deaths | 100 (100, 200) |
| JulyGradual [High R] | VOC High VE | Daily hospital admissions | 800 (800, 1000) |
| JulyGradual [High R] | VOC High VE | Daily hospital deaths | 100 (0, 100) |
| JulyGradual [High R] | VOC High VE | Daily infections | 104500 (79800, 132700) |
| JulyGradual [High R] | VOC High VE | Total additional COVID-19 deaths | 14100 (12400, 16500) |
| JulyGradual [High R] | VOC High VE | Total additional COVID-19 infections | 8235300 (7453300, 9683500) |
| JulyGradual [High R] | VOC Low VE | Daily deaths | 500 (200, 600) |
| JulyGradual [High R] | VOC Low VE | Daily hospital admissions | 5600 (3400, 6100) |
| JulyGradual [High R] | VOC Low VE | Daily hospital deaths | 300 (200, 400) |
| JulyGradual [High R] | VOC Low VE | Daily infections | 272600 (148600, 292400) |
| JulyGradual [High R] | VOC Low VE | Total additional COVID-19 deaths | 38800 (32700, 43800) |
| JulyGradual [High R] | VOC Low VE | Total additional COVID-19 infections | 21629200 (19260900, 23900100) |
| JulyGradual [Low R] | VOC Central VE | Daily deaths | 100 (100, 100) |
| JulyGradual [Low R] | VOC Central VE | Daily hospital admissions | 900 (800, 900) |
| JulyGradual [Low R] | VOC Central VE | Daily hospital deaths | 100 (0, 100) |
| JulyGradual [Low R] | VOC Central VE | Daily infections | 80600 (59100, 104800) |
| JulyGradual [Low R] | VOC Central VE | Total additional COVID-19 deaths | 13500 (9500, 17800) |
| JulyGradual [Low R] | VOC Central VE | Total additional COVID-19 infections | 8430700 (5543700, 11014700) |
| JulyGradual [Low R] | VOC High VE | Daily deaths | 100 (100, 100) |
| JulyGradual [Low R] | VOC High VE | Daily hospital admissions | 800 (800, 900) |

Continued on next page

**Table S16 – continued from previous page**

| Analysis | Scenario | State | Value |
|---|---|---|---|
| JulyGradual [Low R] | VOC High VE | Daily hospital deaths | 100 (0, 100) |
| JulyGradual [Low R] | VOC High VE | Daily infections | 97800 (72500, 129000) |
| JulyGradual [Low R] | VOC High VE | Total additional COVID-19 deaths | 7700 (5900, 10900) |
| JulyGradual [Low R] | VOC High VE | Total additional COVID-19 infections | 4524500 (3292700, 6804300) |
| JulyGradual [Low R] | VOC Low VE | Daily deaths | 100 (0, 200) |
| JulyGradual [Low R] | VOC Low VE | Daily hospital admissions | 1400 (200, 2100) |
| JulyGradual [Low R] | VOC Low VE | Daily hospital deaths | 100 (0, 100) |
| JulyGradual [Low R] | VOC Low VE | Daily infections | 86200 (13500, 108900) |
| JulyGradual [Low R] | VOC Low VE | Total additional COVID-19 deaths | 20000 (12500, 26000) |
| JulyGradual [Low R] | VOC Low VE | Total additional COVID-19 infections | 12973900 (8691700, 16508800) |
| June-21 | VOC Central VE | Daily deaths | 1000 (600, 1400) |
| June-21 | VOC Central VE | Daily hospital admissions | 8500 (5000, 12400) |
| June-21 | VOC Central VE | Daily hospital deaths | 600 (400, 900) |
| June-21 | VOC Central VE | Daily infections | 740000 (455800, 1006200) |
| June-21 | VOC Central VE | Total additional COVID-19 deaths | 34600 (24300, 45800) |
| June-21 | VOC Central VE | Total additional COVID-19 infections | 20831600 (15990100, 25166000) |
| June-21 | VOC High VE | Daily deaths | 800 (500, 1000) |
| June-21 | VOC High VE | Daily hospital admissions | 6100 (4100, 8000) |
| June-21 | VOC High VE | Daily hospital deaths | 500 (300, 700) |
| June-21 | VOC High VE | Daily infections | 646400 (463300, 777000) |
| June-21 | VOC High VE | Total additional COVID-19 deaths | 25700 (18800, 32600) |
| June-21 | VOC High VE | Total additional COVID-19 infections | 16720000 (13693900, 19181500) |
| June-21 | VOC Low VE | Daily deaths | 1000 (500, 1500) |
| June-21 | VOC Low VE | Daily hospital admissions | 11200 (5900, 17500) |
| June-21 | VOC Low VE | Daily hospital deaths | 600 (300, 1000) |
| June-21 | VOC Low VE | Daily infections | 756100 (445300, 1059600) |
| June-21 | VOC Low VE | Total additional COVID-19 deaths | 38100 (24900, 51800) |
| June-21 | VOC Low VE | Total additional COVID-19 infections | 24252700 (17977700, 30236600) |
| June-21 [High R] | VOC Central VE | Daily deaths | 1500 (1100, 2000) |
| June-21 [High R] | VOC Central VE | Daily hospital admissions | 13700 (9300, 17300) |
| June-21 [High R] | VOC Central VE | Daily hospital deaths | 1000 (700, 1300) |

Continued on next page

**Table S16 – continued from previous page**

| Analysis | Scenario | State | Value |
|---|---|---|---|
| June-21 [High R] | VOC Central VE | Daily infections | 1043200 (758800, 1259900) |
| June-21 [High R] | VOC Central VE | Total additional COVID-19 deaths | 48800 (38400, 60300) |
| June-21 [High R] | VOC Central VE | Total additional COVID-19 infections | 25800700 (21919900, 29248800) |
| June-21 [High R] | VOC High VE | Daily deaths | 1200 (800, 1400) |
| June-21 [High R] | VOC High VE | Daily hospital admissions | 9200 (7000, 10900) |
| June-21 [High R] | VOC High VE | Daily hospital deaths | 800 (500, 1000) |
| June-21 [High R] | VOC High VE | Daily infections | 881300 (684700, 994000) |
| June-21 [High R] | VOC High VE | Total additional COVID-19 deaths | 35400 (28000, 41700) |
| June-21 [High R] | VOC High VE | Total additional COVID-19 infections | 20052700 (17550300, 21907000) |
| June-21 [High R] | VOC Low VE | Daily deaths | 1600 (1000, 2200) |
| June-21 [High R] | VOC Low VE | Daily hospital admissions | 19100 (12800, 25900) |
| June-21 [High R] | VOC Low VE | Daily hospital deaths | 1000 (700, 1400) |
| June-21 [High R] | VOC Low VE | Daily infections | 1192000 (852900, 1509400) |
| June-21 [High R] | VOC Low VE | Total additional COVID-19 deaths | 53700 (41300, 66700) |
| June-21 [High R] | VOC Low VE | Total additional COVID-19 infections | 30368700 (25794700, 34942100) |
| June-21 [Low R] | VOC Central VE | Daily deaths | 500 (200, 800) |
| June-21 [Low R] | VOC Central VE | Daily hospital admissions | 4000 (1900, 7400) |
| June-21 [Low R] | VOC Central VE | Daily hospital deaths | 300 (100, 500) |
| June-21 [Low R] | VOC Central VE | Daily infections | 392000 (200300, 640000) |
| June-21 [Low R] | VOC Central VE | Total additional COVID-19 deaths | 20500 (11800, 31400) |
| June-21 [Low R] | VOC Central VE | Total additional COVID-19 infections | 14387400 (9071000, 19931000) |
| June-21 [Low R] | VOC High VE | Daily deaths | 400 (200, 700) |
| June-21 [Low R] | VOC High VE | Daily hospital admissions | 3300 (1700, 5300) |
| June-21 [Low R] | VOC High VE | Daily hospital deaths | 300 (100, 400) |
| June-21 [Low R] | VOC High VE | Daily infections | 409400 (237000, 580000) |
| June-21 [Low R] | VOC High VE | Total additional COVID-19 deaths | 16300 (10100, 23700) |
| June-21 [Low R] | VOC High VE | Total additional COVID-19 infections | 12345000 (8638400, 15924200) |
| June-21 [Low R] | VOC Low VE | Daily deaths | 400 (200, 900) |
| June-21 [Low R] | VOC Low VE | Daily hospital admissions | 4900 (1700, 10600) |
| June-21 [Low R] | VOC Low VE | Daily hospital deaths | 300 (100, 600) |
| June-21 [Low R] | VOC Low VE | Daily infections | 383200 (152800, 765900) |

Continued on next page

**Table S16 – continued from previous page**

| Analysis | Scenario | State | Value |
|---|---|---|---|
| June-21 [Low R] | VOC Low VE | Total additional COVID-19 deaths | 22400 (11500, 36800) |
| June-21 [Low R] | VOC Low VE | Total additional COVID-19 infections | 16350000 (9564200, 24188500) |
| JuneGradual | VOC Central VE | Daily deaths | 400 (100, 600) |
| JuneGradual | VOC Central VE | Daily hospital admissions | 3400 (1300, 4400) |
| JuneGradual | VOC Central VE | Daily hospital deaths | 200 (100, 400) |
| JuneGradual | VOC Central VE | Daily infections | 285700 (117900, 348200) |
| JuneGradual | VOC Central VE | Total additional COVID-19 deaths | 23900 (16900, 32700) |
| JuneGradual | VOC Central VE | Total additional COVID-19 infections | 15807100 (11896200, 20274600) |
| JuneGradual | VOC High VE | Daily deaths | 300 (100, 500) |
| JuneGradual | VOC High VE | Daily hospital admissions | 2200 (1000, 3400) |
| JuneGradual | VOC High VE | Daily hospital deaths | 200 (100, 300) |
| JuneGradual | VOC High VE | Daily infections | 252400 (114100, 365900) |
| JuneGradual | VOC High VE | Total additional COVID-19 deaths | 15600 (10800, 21600) |
| JuneGradual | VOC High VE | Total additional COVID-19 infections | 11879200 (8915900, 14865800) |
| JuneGradual | VOC Low VE | Daily deaths | 500 (100, 600) |
| JuneGradual | VOC Low VE | Daily hospital admissions | 5600 (1900, 6800) |
| JuneGradual | VOC Low VE | Daily hospital deaths | 300 (100, 400) |
| JuneGradual | VOC Low VE | Daily infections | 337000 (125300, 409200) |
| JuneGradual | VOC Low VE | Total additional COVID-19 deaths | 30600 (21200, 41700) |
| JuneGradual | VOC Low VE | Total additional COVID-19 infections | 20315200 (15256900, 25998500) |
| JuneGradual [High R] | VOC Central VE | Daily deaths | 600 (300, 800) |
| JuneGradual [High R] | VOC Central VE | Daily hospital admissions | 5000 (2600, 5900) |
| JuneGradual [High R] | VOC Central VE | Daily hospital deaths | 400 (200, 500) |
| JuneGradual [High R] | VOC Central VE | Daily infections | 390900 (174000, 525900) |
| JuneGradual [High R] | VOC Central VE | Total additional COVID-19 deaths | 31800 (25200, 40000) |
| JuneGradual [High R] | VOC Central VE | Total additional COVID-19 infections | 19291000 (15992700, 22923000) |
| JuneGradual [High R] | VOC High VE | Daily deaths | 400 (200, 600) |
| JuneGradual [High R] | VOC High VE | Daily hospital admissions | 3000 (1600, 4200) |
| JuneGradual [High R] | VOC High VE | Daily hospital deaths | 300 (100, 400) |
| JuneGradual [High R] | VOC High VE | Daily infections | 326600 (178100, 417400) |
| JuneGradual [High R] | VOC High VE | Total additional COVID-19 deaths | 19900 (15200, 25700) |

Continued on next page

**Table S16 – continued from previous page**

| Analysis | Scenario | State | Value |
|---|---|---|---|
| JuneGradual [High R] | VOC High VE | Total additional COVID-19 infections | 13969000 (11495000, 16573400) |
| JuneGradual [High R] | VOC Low VE | Daily deaths | 700 (300, 900) |
| JuneGradual [High R] | VOC Low VE | Daily hospital admissions | 8500 (3700, 10000) |
| JuneGradual [High R] | VOC Low VE | Daily hospital deaths | 500 (200, 600) |
| JuneGradual [High R] | VOC Low VE | Daily infections | 490400 (201600, 604600) |
| JuneGradual [High R] | VOC Low VE | Total additional COVID-19 deaths | 40600 (31700, 50200) |
| JuneGradual [High R] | VOC Low VE | Total additional COVID-19 infections | 24831000 (20722200, 29193200) |
| JuneGradual [Low R] | VOC Central VE | Daily deaths | 200 (100, 400) |
| JuneGradual [Low R] | VOC Central VE | Daily hospital admissions | 1900 (500, 3300) |
| JuneGradual [Low R] | VOC Central VE | Daily hospital deaths | 100 (0, 300) |
| JuneGradual [Low R] | VOC Central VE | Daily infections | 179700 (53500, 291300) |
| JuneGradual [Low R] | VOC Central VE | Total additional COVID-19 deaths | 16100 (9100, 25600) |
| JuneGradual [Low R] | VOC Central VE | Total additional COVID-19 infections | 11773800 (7275400, 17399500) |
| JuneGradual [Low R] | VOC High VE | Daily deaths | 200 (100, 400) |
| JuneGradual [Low R] | VOC High VE | Daily hospital admissions | 1400 (500, 2800) |
| JuneGradual [Low R] | VOC High VE | Daily hospital deaths | 100 (0, 200) |
| JuneGradual [Low R] | VOC High VE | Daily infections | 180500 (71000, 307700) |
| JuneGradual [Low R] | VOC High VE | Total additional COVID-19 deaths | 11400 (6600, 17600) |
| JuneGradual [Low R] | VOC High VE | Total additional COVID-19 infections | 9392500 (6039500, 13087500) |
| JuneGradual [Low R] | VOC Low VE | Daily deaths | 300 (0, 500) |
| JuneGradual [Low R] | VOC Low VE | Daily hospital admissions | 2900 (600, 4500) |
| JuneGradual [Low R] | VOC Low VE | Daily hospital deaths | 200 (0, 300) |
| JuneGradual [Low R] | VOC Low VE | Daily infections | 198800 (44700, 317700) |
| JuneGradual [Low R] | VOC Low VE | Total additional COVID-19 deaths | 20000 (11300, 33000) |
| JuneGradual [Low R] | VOC Low VE | Total additional COVID-19 infections | 14693600 (9137100, 22096100) |

##### 7.3 Sensitivity analyses

The results of the sensitivity analyses are computed with Shapley values [53]. Shapley values are rooted in game theory and treat each variable as 'players' in a 'game' that contribute towards the final simulation. In our analyses we use Shapley values to firstly calculate the importance of each 'level' of a feature, where a level is a possible value the feature can take (e.g. optimistic/central/pessimistic for VE), and secondly to describe the overall importance of the feature. In the simplest explanation, a Shapley value is the expected increase or decrease in a projection caused by the variable (or level) of interest. For example, in Figure S26, the 'July-19 [High R]' scenario is expected to result in 20,706 more deaths than other scenarios, where 'other scenarios' is the expected number of deaths in all scenarios except 'July-19 [High R]'. To derive a measure of variable importance, we take the expectation of the absolute Shapley value across all levels in a variable, such as in Figure S27. Here, variables are ordered according to importance (top is most important and bottom is least) and the numbers correspond to the absolute expected difference. These plots provide a measure of variable importance by comparing the magnitude of the expected difference, e.g. vaccine effectiveness is more important than waning rate as it can lead to an almost 10-fold difference in number of expected deaths.

To demonstrate how these are computed, take the bottom-left cell as an example in which we compute the Shapley value for the 'July-19 [High R]' scenario on the total additional COVID-19 deaths, this is computed as follows, let

- $f_{St}$ be a simulation for a given state, $St$ ('Total deaths'), from our fitted model with all variables as described previously with different combinations of the variables listed below;
- $SC_i$ be the $i$th level of the scenario variable ('July-19 [High R]');
- $VE_j$ be the $j$th level of the vaccine effectiveness variable, e.g. 'optimistic';
- $CI_k$ be the $k$th level of the cross immunity variable, e.g. 'optimistic';
- $WR_l$ be the $l$th level of the waning rate variable, e.g. 'optimistic';
- $\phi_{v_i}$ be the Shapley value for the $i$th level of the $v$th variable (Shapley for 'July-19 [High R]' of the scenario variable);
- $n_v$ be the number of levels in the variable of interest, $v$ ($n_v = 6$);
- $n'_v$ be the total number of levels in all variables except $v$ ($n_{v'} = 27$).

Then for the scenario variable:

$$\phi_{v_i} = \frac{1}{n_v - 1} \sum_{i \neq i'} \frac{1}{n_{v'}} \sum_{j,k,l} f_{St}(SC'_i, VE_j, CI_k, WR_l) - f_{St}(SC_i, VE_j, CI_k, WR_l) \Big|_{v=SC} \tag{216}$$

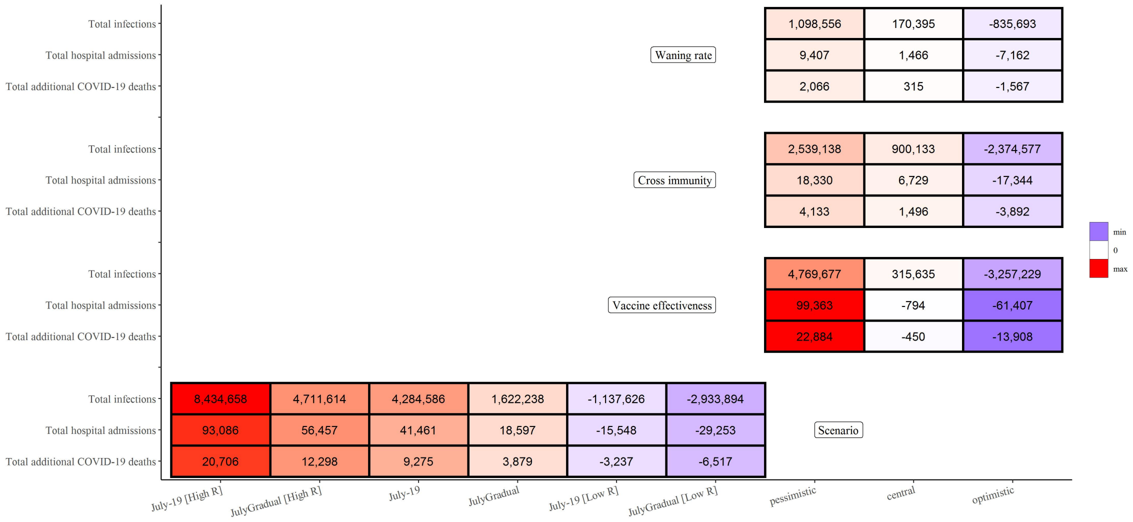

**Figure S26:** Shapley values computed from three different simulated states (total deaths, admissions, and hospitalisations) on 1 June 2022 from the model fit with ‘central’ assumptions. Shapley values are computed as the expected difference in the measured state within variables. For example, the bottom left cell indicates that the July-19 [High R] scenario is expected to lead to 20,706 more deaths than other scenarios.

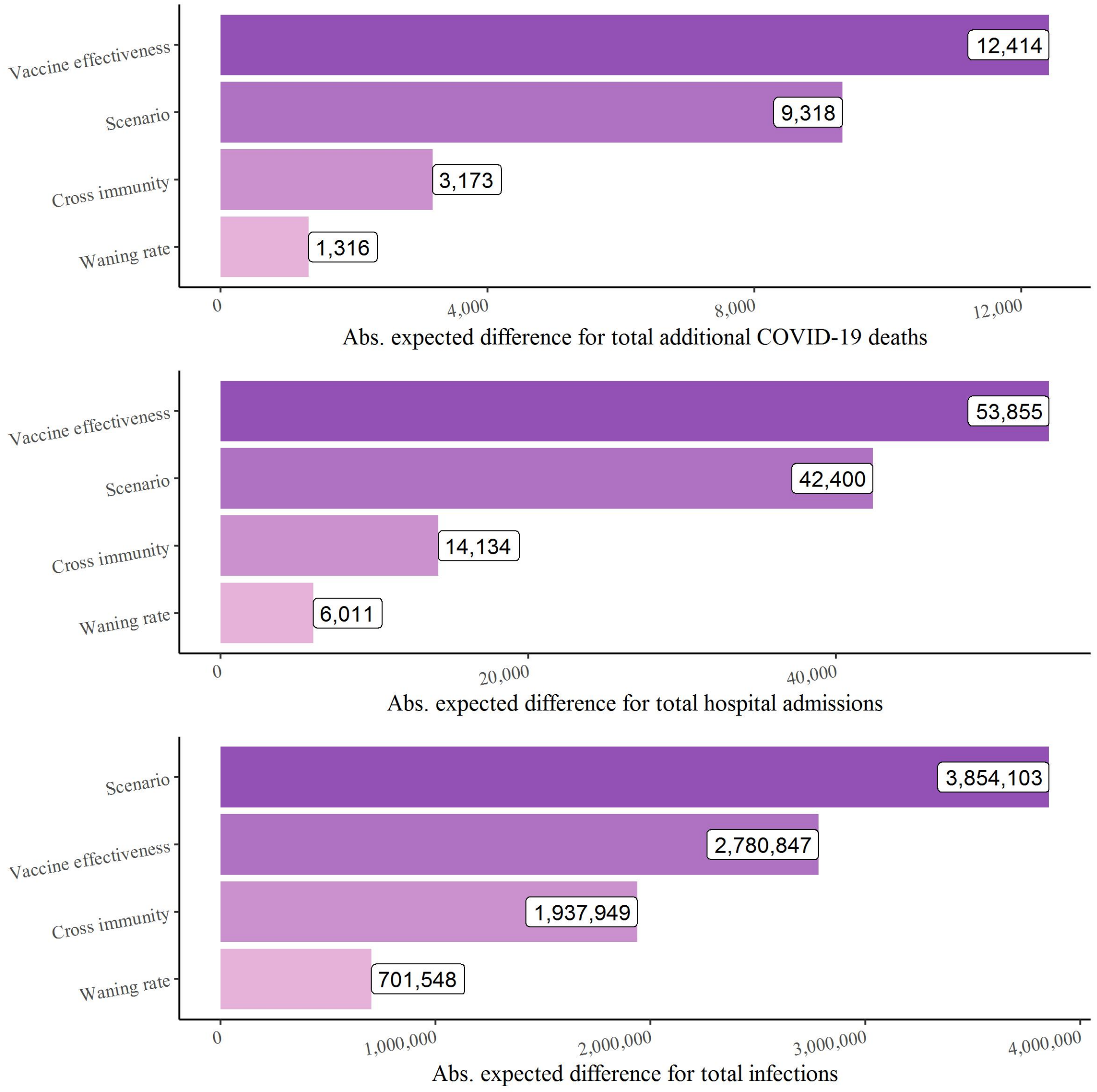

**Figure S27:** Absolute expected Shapley values for the model with central assumptions for total additional COVID-19 deaths (top), hospital admissions (middle), and COVID-19 infections (bottom) by 1 June 2022 (counted from 19 July 2021). Each plot is ordered from most important variable (top) to least (bottom), according to the expected absolute Shapley value with expectations taken over feature levels.

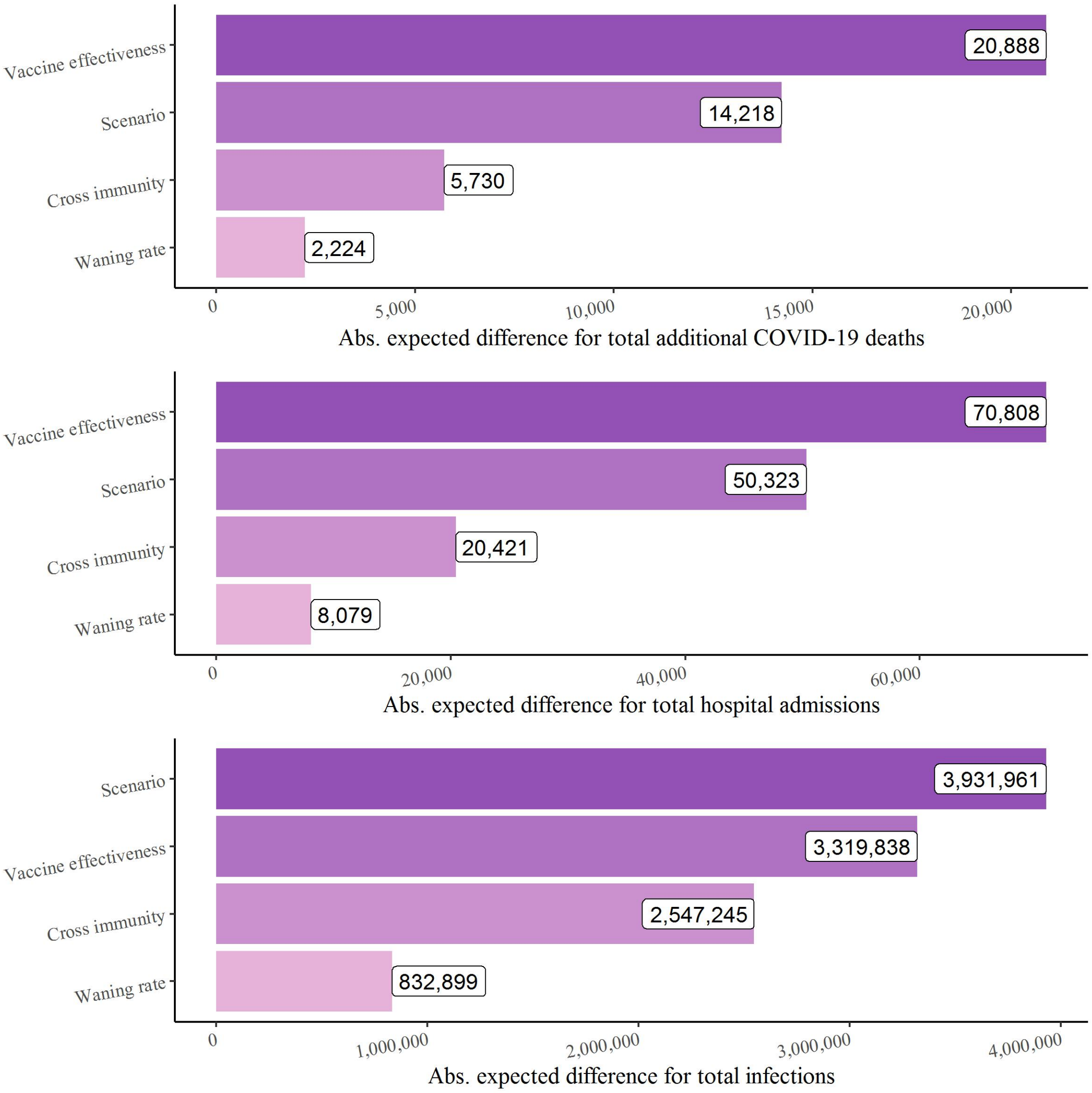

**Figure S28:** Absolute expected Shapley values for the model with optimistic assumptions for total additional COVID-19 deaths (top), hospital admissions (middle), and COVID-19 infections (bottom) by 1 June 2022 (counted from 19 July 2021). Each plot is ordered from most important variable (top) to least (bottom), according to the expected absolute Shapley value with expectations taken over feature levels.

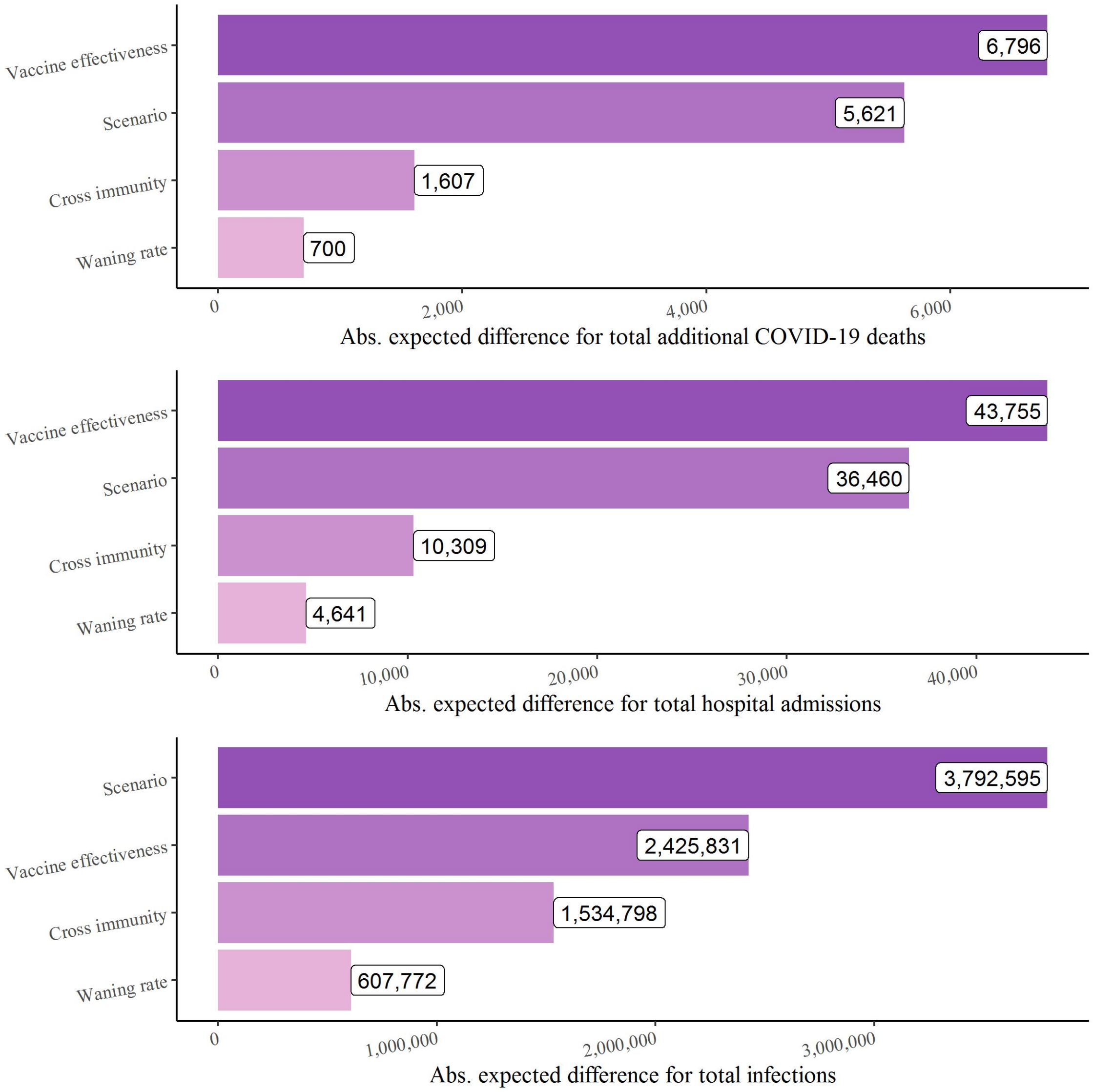

**Figure S29:** Absolute expected Shapley values for the model with pessimistic assumptions for total additional COVID-19 deaths (top), hospital admissions (middle), and COVID-19 infections (bottom) by 1 June 2022 (counted from 19 July 2021). Each plot is ordered from most important variable (top) to least (bottom), according to the expected absolute Shapley value with expectations taken over feature levels.

#### Symbol Glossary

| Symbol | Definition |
|---|---|
| | **Abbreviations** |
| CHW | Carehome workers |
| CHR | Carehome residents |
| VE | Vaccine effectiveness |
| AZ | AstraZeneca ChAdOx1 nCoV-19 (AZD1222) vaccine |
| PF | Pfizer-BioNTech COVID-19 Vaccine BNT162b2 |
| Mod | Moderna mRNA-1273 vaccine |
| | **Model Compartments** |
| $S^{ik}$ | Susceptible |
| $E^{i,j,k}$ | Exposed |
| $I_P^{i,j,k}$ | Infected pre-symptomatic |
| $I_A^{i,j,k}$ | Infected aymptomatic |
| $I_{C_1}^{i,j,k}$ | Symptomatic infected (infectious) |
| $I_{C_2}^{i,j,k}$ | Symptomatic infected (not infectious) |
| $G_D^{i,j,k}$ | Severe disease, not hospitalised |
| $D^{i,j,k}$ | Deceased (as a result of COVID-19) |
| $R^{i,j,k}$ | Recovered |
| $V_k$ | Vaccination strata |
| $ICU_{pre}^{i,j,k}$ | Awaiting admission to ICU |
| $ICU_{W_R}^{i,j,k}$ | Hospitalised in ICU, leading to recovery |
| $ICU_{W_D}^{i,j,k}$ | Hospitalised in ICU, leading to death following step-down from ICU |
| $ICU_D^{i,j,k}$ | Hospitalised in ICU, leading to death |
| $W_D^{i,j,k}$ | Step-down post-ICU period, leading to death |
| $W_R^{i,j,k}$ | Step-down recovery period |
| $H_D^{i,j,k}$ | Hospitalised on general ward leading to death |
| $H_R^{i,j,k}$ | Hospitalised on general ward leading to recovery |
| | **Model Parameters** |
| $p_H^{i,j,k}(t)$ | Probability of hospitalisation given symptomatic |
| $p_{G_D}^{i,j,k}$ | Probability of severe disease but not hospitalised |
| $p_{ICU}^{i}(t)$ | Probability of ICU admission given hospitalised |
| $p_{H_D}^{i,j,k}(t)$ | Probability of death given hospitalised and not in ICU |
| $p_{ICU_D}^{i,j,k}(t)$ | Probability of death given ICU |
| $p_{W_D}^{i,j,k}(t)$ | Probability of death after discharge |
| $\chi^{i,j,k}(t)$ | Susceptibility of an individual to variant $j$ |
| $\xi^{i,j,k}(t)$ | Infectivity of an individual infected with variant $j$ |
| $\lambda^{i,j,k}(t)$ | Variant-specific force of infection |
| $\Lambda^{i,k}(t)$ | Combined force of infection (both variants) |
| $\zeta^{i,k}(t)$ | Rate of progression from vaccine strata $k$ to $k+1$ |

Continued on next page

| Table S17 – continued from previous page | |
|---|---|
| Symbol | Definition |
| $\gamma_x$ | Rate of progression from compartment $x$ |
| $R^j(t)$ | R number for variant $j$ at time $t$ |
| $R_e^j(t)$ | Effective R number for variant $j$ at time $t$ |
| $t_0$ | Regional outbreak start date |
| $t_{Delta}$ | Delta seeding date |
| $\sigma$ | Delta transmission advantage |
| $m_{i,i'}(t)$ | Person-to-person transmission rate |
| $c_{i,j}$ | Person-to-person contact rate |
| $\beta(t)$ | Transmission rate |
| $\beta_i$ | Transmission rate at change-point $t_i$ |
| $\Theta_{i,j,k}(t)$ | Weighted number of infectious individuals |
| $\varepsilon$ | Relative reduction in contacts between CHR and general population |
| $\Delta_I^{i,k}$ | Mean duration of infectiousness weighted by infectivity |
| $\hat{\sigma}(t)$ | Ratio of effective reproduction number for Alpha and Delta ($R^{Alpha}(t)$ and $R^{Delta}(t)$). |
| | **Vaccine Effectiveness vs.** |
| $e_{inf}$ | Infection |
| $e_{sympt}$ | Symptoms |
| $e_{SD}$ | Severe disease |
| $e_{death}$ | Death |
| $e_{sympt\|inf}$ | Symptoms given infection |
| $e_{SD\|sympt}$ | Severe disease given symptoms |
| $e_{death\|SD}$ | Death given severe disease |
| $e_{ins}$ | Infectiousness |
| | **Fixed Parameters** |
| $p_C^{i,j,k}$ | Probability of being symptomatic given infected |
| $p^*(t)$ | Probability of COVID-19 diagnosis confirmed prior to hospital admission |
| $\gamma_U$ | Rate at which unconfirmed hospital patients are confirmed as infected |
| $\gamma_R$ | Rate at which natural immunity from infection wanes |
| $p_{sero_{pos}}$ | Probability of seroconversion following infection |
| $p_{sero_{spec}}$ | Specificity of serology test |
| $p_{sero_{sens}}$ | Sensitivity of serology test |
| $1/\gamma_{sero_{pre}}$ | Mean time to seroconversion from onset of infectiousness |
| $1/\gamma_{sero^1_{pos}}$ | Mean duration of seropositivity (Euroimmun assay) |
| $1/\gamma_{sero^2_{pos}}$ | Mean duration of seropositivity (Roche N) |
| $\eta$ | Proportion of cross-immunity to Delta following infection from Alpha |
| $\theta_{I_A}$ | Infectivity of an asymptomatic individual, relative to a symptomatic one |
